# Toward a geography of community health workers in Niger: a geospatial analysis

**DOI:** 10.1101/2021.02.02.21250043

**Authors:** Nicholas P Oliphant, Nicolas Ray, Khaled Bensaid, Adama Ouedraogo, Asma Yaroh Gali, Oumarou Habi, Ibhrahim Maazou, Rocco Panciera, Maria Muñiz, Samuel OM Manda, Zeynabou Sy, Debra Jackson, Tanya Doherty

**Author notes:** **CORRESPONDENCE** Nicholas P Oliphant, School of Public Health, University of the Western Cape, Private Bag X17, Belleville 7535, Republic of South Africa.

## Abstract

**Background:** Little is known about the geography of community health workers (CHWs), their contribution to geographical accessibility of primary health care (PHC) services, and strategies for optimizing efficiency of CHW deployment in the context of universal health coverage (UHC).

**Methods:** Using a complete georeferenced census of front-line health facilities and CHWs in Niger and other high resolution spatial datasets, we modelled travel times to front-line health facilities and CHWs between 2000-2013, accounting for training, essential commodities, and maximum population capacity. We estimated additional CHWs needed to maximize geographical accessibility of the population beyond the reach of existing front-line health facilities and CHWs. We assessed the efficiency of geographical targeting of the existing CHW network compared to modelled CHW networks designed to optimize geographical targeting of the estimated population, under-five deaths, and *plasmodium falciparum* malaria cases.

**Results:** The percent of the population within 60 minutes walking to the nearest CHW increased from 0·0% to 17·5% between 2000-2013, with 15·5% within 60 minutes walking to the nearest CHW trained on integrated community case management (iCCM) – making PHC services and iCCM, specifically, geographically accessible for an estimated 2·3 million and 2·0 million additional people, respectively. An estimated 10·4 million people (59·0%) remained beyond a 60-minute catchment of front-line health facilities and CHWs. Optimal deployment of 8064 additional CHWs could increase geographic coverage of the estimated total population from 41·5% to 73·6%. Geographical targeting of the existing CHW network was inefficient but optimized CHW networks could improve efficiency by 55·0%-81·9%, depending on targeting metric.

**Interpretations:** We provide the first high-resolution maps and estimates of geographical accessibility to CHWs at national scale, highlighting improvements between 2000-2013 in Niger, geographies where gaps remained, approaches for improving targeting, and the importance of putting CHWs on the map to inform planning in the context of UHC.

KEY QUESTIONS

What is already known?

- Previous studies have estimated geographical accessibility (as travel time) to CHWs for subnational areas only^1-4^ and have assessed efficiency of the distribution of hospitals in low and middle-income countries.^5^

What are the new findings?

- The percent of the population within 60 minutes walking to the nearest CHW increased from 0·0% to 17·5% between 2000-2013, with 15·5% within 60 minutes walking to the nearest CHW trained on integrated community case management (iCCM) – making PHC services and iCCM, specifically, geographically accessible for an estimated 2·3 million and 2·0 million additional people, respectively.
- An estimated 10·4 million people (59·0%) remained beyond a 60-minute catchment of front-line health facilities and CHWs in 2013, with important variation across subnational geographies, training of CHWs, and availability of essential commodities.
- Optimal deployment of 8064 additional CHWs could increase geographic coverage of the estimated total population from 41·5% to 73·6%, providing physical access to PHC services for an additional 5·7 million people not covered in 2013.
- Optimized CHW networks increased efficiency of geographical targeting compared to the existing CHW network by 55·0%-81·9%, depending on targeting metric.

What do the new findings imply?

- Geographical accessibility to primary health care services, including iCCM, improved in Niger between 2000-2013 with important contributions by CHWs.
- Gaps in geographical accessibility remained as of 2013 but scale-up of the CHW network, using the scale-up approach described in this study, could substantially increase geographical accessibility of PHC services.
- The efficiency of geographical targeting of the existing network of CHWs was suboptimal. The approach for optimizing efficiency of geographical targeting described in this study could be used to improve geographical targeting of CHW deployment in Niger and other countries.
- This work is a first step toward establishing a geography of CHWs in Niger and is a call to action to put CHWs on the map globally to inform health system planning and maximize geographical accessibility, efficiency, and impact of investments in the context of UHC.

## BACKGROUND

Community health workers (CHWs) can play an important role in improving equitable access to quality primary health care (PHC) and other services in the context of Universal Health Coverage (UHC) as front-line service providers and as a trusted bridge between health systems and communities.^6-8^ CHWs typically focus on maternal, newborn and child health and nutrition, providing a range of preventive, health promotion and curative services – including single disease or integrated community case management (iCCM).^9^ iCCM is the provision of integrated case management services for two or more childhood illnesses among children less than five years of age by CHWs, where geographical accessibility (i.e. physical access) to health facility-based case management services is limited.^10^ However, little is known about the geography of CHWs globally – how many there are, where they are deployed, their contribution to geographical accessibility to health services, efficiency of their deployment, and needs for further scale-up.^6,9^ In this article, we provide the first description of the geography of CHWs at national scale in Niger.

## DATA AND METHODS

In this section we describe the study settings, data, and methods used. Supplementary Appendix 1 provides a simplified analysis flow (pages 3-5) and additional details on the data and methods (pages 13-56).

### Study settings

Niger is a large country (1·267 million sq. kilometers) in the Sahel region of sub-Saharan Africa,^11^ and is ranked among the poorest countries in the world.^12^ Infrastructure, including roads, is underdeveloped^13^ and possession of motorized transportation is low.^14^ The North is predominantly bare/sparse desert, and the South is predominantly herbaceous vegetation and cropland.^15^ The climate is Sudanese, with a long dry season from October to May and a short rainy season from May to September.^11^

During the period of focus of this study, 2000-2013, Niger was divided into four political administrative levels: communes, departments, regions, and national.^11^ The health system of Niger included a public and private sector organized in a decentralized, pyramidal structure with three administrative levels overseen by the Ministry of Public Health (MOPH). Details on the health system are provided in Supplementary Appendix 1, pages 22-24. Our analysis focuses on the first level (periphery): the *Centre de Santé Intégré* (CSI) and *Case de Santé* (CS) networks of the public sector. As of December 2012, there were 856 CSI, offering a minimum package of services, focused on PHC, referral from and counter-referral to the CS, and supervision of the CS.^11^ CSI were typically staffed by nurses and in certain large communes by a generalist doctor and midwives and were intended to serve a maximum population of 5000-15000 inhabitants, depending on population density.^11,16^ According to national norms, CS were intended to be situated 5km beyond a supervising CSI and served a population of 2500 to 5000.^16^ CS provided a minimum package of services, focused on PHC, including prevention services, health promotion services, and services for reproductive, maternal, newborn and child health, including iCCM. CS were typically staffed by a cadre of paid, full-time CHWs called *agent de santé* communautaire (ASC) and/or, in some cases, a nurse.^17^ CS and ASC were scaled-up between 2000-2013 – a period of considerable progress on under-five mortality.^17,18^ As of December 2012, there were 2451 CS.^11^ Some CS were supported by one or more volunteer CHWs called *relais communautiare* (RC), providing health promotion and prevention interventions in the communities within the catchment area (typically a 5km radius) of the CS.^11,16^ The MOPH in Niger plans to scale-up RC – some targeted to communities beyond 5km of CS or CSI to provide a standard package of preventive, promotive and curative services, including iCCM.^19^

### Data

We obtained spatial datasets for the following inputs: administrative boundaries (levels 0-3),^20^ health service delivery networks (CSI, CS and ASC),^20^ digital elevation model,^22^ land cover,^15^ roads,^23^ rivers and other water bodies (treated as barriers to movement where no road crossed),^24^ travel scenarios, modelled estimates for population counts for 2000-2013^25^ and 2015,^26^ modelled estimates for the annual mean under-five mortality rate in 2013,^27^ and modelled estimates for the annual mean incidence of Plasmodium falciparum (*Pf*) malaria among all ages (0-99 years) in 2013.^28^ We prepared the input datasets in a coordinate reference system for Niger at 100m x 100m resolution for our analysis of accessibility coverage and 1km x 1km for our analysis of geographic coverage, targeting and scale-up. Further details are in Supplementary Appendix 1 (pages 21-33).

We prepared travel speed tables for two travel scenarios: 1) walking in dry conditions and 2) walking to the nearest road and then using motorized transportation (assumed to be immediately available) in dry conditions. We set travel speeds by travel scenario for each land cover class and road class. Travel speeds were adapted from previous studies and experience in Niger and broader sub-Saharan Africa.^29,30^

### Assessing geographical accessibility

We assessed geographical accessibility through two measures: accessibility coverage and geographic coverage.

We defined accessibility coverage as the estimated percentage of people within a given travel time to the nearest health service delivery location of a given health service delivery network, accounting for travel speeds of different modes of transportation over different land cover classes and slope, with the direction of travel toward the health service delivery location.^29^ We estimated accessibility coverage at 100m x 100m resolution for the CSI, CS, and CS-ASC (includes CS with or without ASC and the small number of ASC sites not within a CS) networks in 2013 – and for the ASC network by gender, year of deployment (2000-2013), training, and availability of essential commodities – using 30-minute and 60-minute cutoffs for administrative levels 0-3 and the two travel scenarios. We used 30-minute and 60-minute cutoffs as previous analyses have shown careseeking decays as a function of travel time after these cutoffs^31^ and they are clinically relevant (e.g., for prompt treatment of severe illness).^32^ The analysis was constrained to national borders but allowed for travel across subnational administrative boundaries. We used the “geographic accessibility” module within AccessMod 5 (v5·6 ·48)^29^ to calculate travel time layers and the “zonal statistics” module to calculate the zonal statistics for each travel time layer by administrative level.

We defined geographic coverage as the theoretical catchment area of a health service delivery location, within a maximum travel time, accounting for the mode of transportation and the maximum population coverage capacity of the type of health service delivery location.^29^ We used the “geographic coverage” module of AccessMod 5 (v5·6 ·48)^29^ to estimate geographic coverage for the CSI and CS-ASC networks in 2013 at 1km x 1km resolution for the two travel scenarios. The maximum travel time was set at 60 minutes. The maximum population capacity was set at 10000 for CSI and 2500 for CS-ASC based on norms of the MOPH of Niger.^18^ The maximum extent of a catchment was therefore delimited by the maximum travel time of 60 minutes except in cases where the estimated population in the catchment exceeded the maximum population capacity of the health service delivery location – in which case the extent of the catchment was smaller than the maximum travel time and was defined by the area containing the estimated population, up to the maximum population capacity.

### Assessing geographic coverage of a hypothetical scale-up network of RC

To estimate the number of RC needed to maximize geographical accessibility of the population beyond the geographic coverage of the existing CSI and CS-ASC networks, we simulated a hypothetical network of RC in grid cells with at least 250 people in 2013 located beyond the geographic coverage of the existing CSI and CS-ASC networks at 1km x 1km resolution, using a ratio of 1 RC per 1000 population (with a minimum threshold of 250 people to allocate 1 RC). We conducted a geographic coverage analysis at 1km x 1km resolution to estimate the percent of the estimated residual population that could be covered by the hypothetical RC network, within a maximum travel time of 60 min walking to the nearest RC and maximum population capacity of 1000 for each RC.

### Assessing efficiency of geographical targeting

We assessed the efficiency of geographical targeting of the CS-ASC network, using the concept of technical efficiency. We defined technical efficiency as the maximization of a health outcome (geographic coverage) for a given set of inputs (the number of CS-ASC). ^33^ We used the estimated population, under-five deaths, and *Pf* malaria cases beyond the geographic coverage (60 minutes walking) of the CSI network in 2013 – hereafter called the estimated residual population, under-five deaths, and *Pf* malaria cases, respectively – as the “populations” to target in our geographical targeting analysis. We assessed the efficiency of geographical targeting of the existing CS-ASC network with three metrics a) geographic coverage of the estimated residual population b) geographic coverage of the estimated residual under-five deaths and c) geographic coverage of the estimated residual *Pf* malaria cases among all ages beyond the catchment of the CSI network in 2013 at 1km x 1km resolution compared to three hypothetical CS-ASC networks designed to optimize metrics a-c. For comparison with the existing CS-ASC network (n=2550) we restricted the hypothetical networks to the 2550 sites that maximized geographic coverage of the targeted population. We assessed the potential effect of uncertainty of the estimates for under-five deaths and *Pf* malaria cases among all ages on interpretation of our targeting results (see Supplementary Appendix 1, pages 54 and 57, and Supplementary Appendix 7).

### Patient and public involvement statement

We did not involve patients or the public in this study.

## RESULTS

### Accessibility coverage

Accessibility coverage of the ASC network increased from 0·0% to 17·5% between 2000-2013, with large variation at subnational levels, given a 60-minute cutoff and walking scenario (Table 1, Figure 1, Supplementary Appendix 2). Video 1 and video 2 show the evolution of accessibility coverage of the ASC network between 2000-2013 by mode of transportation.

**Table 1.**
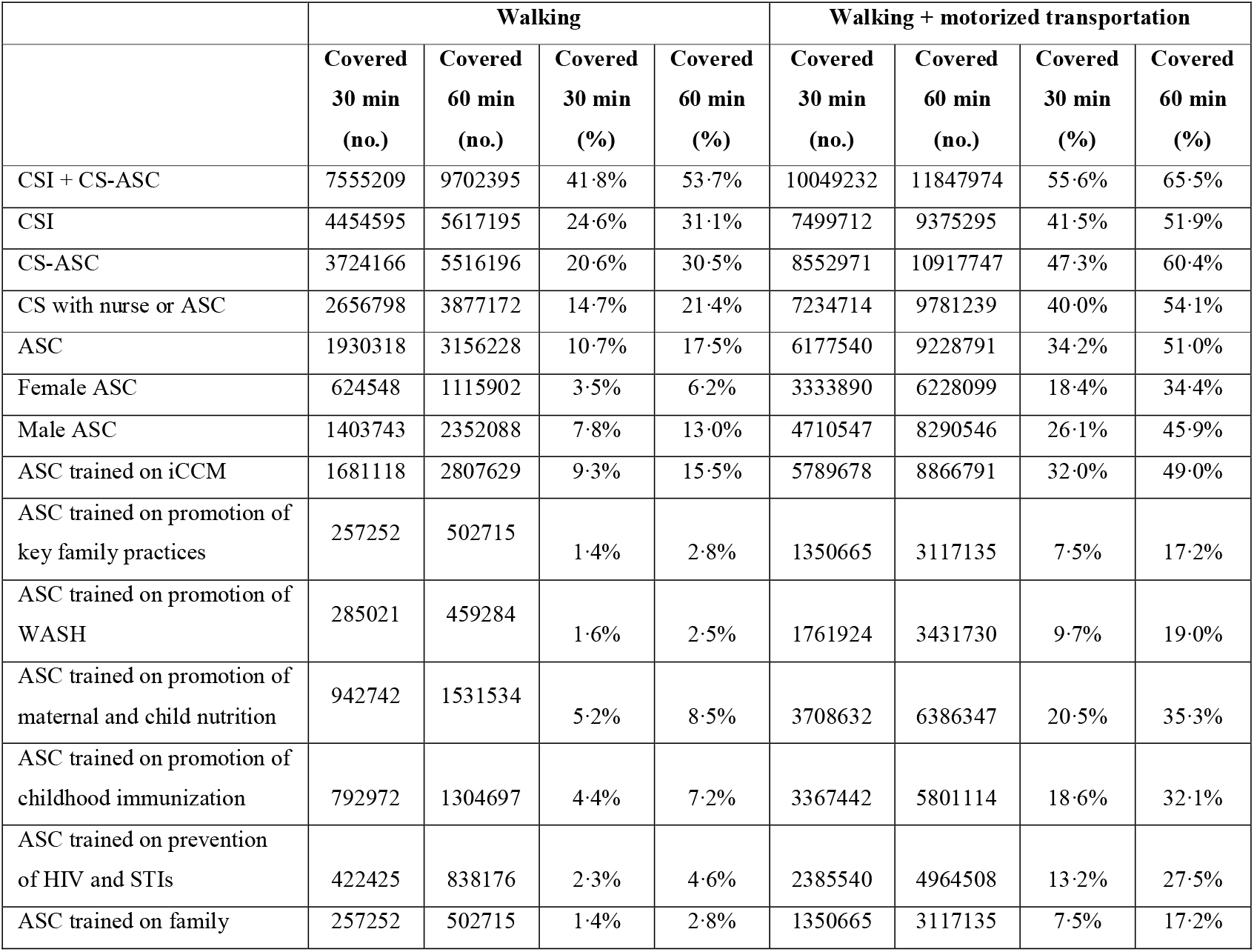
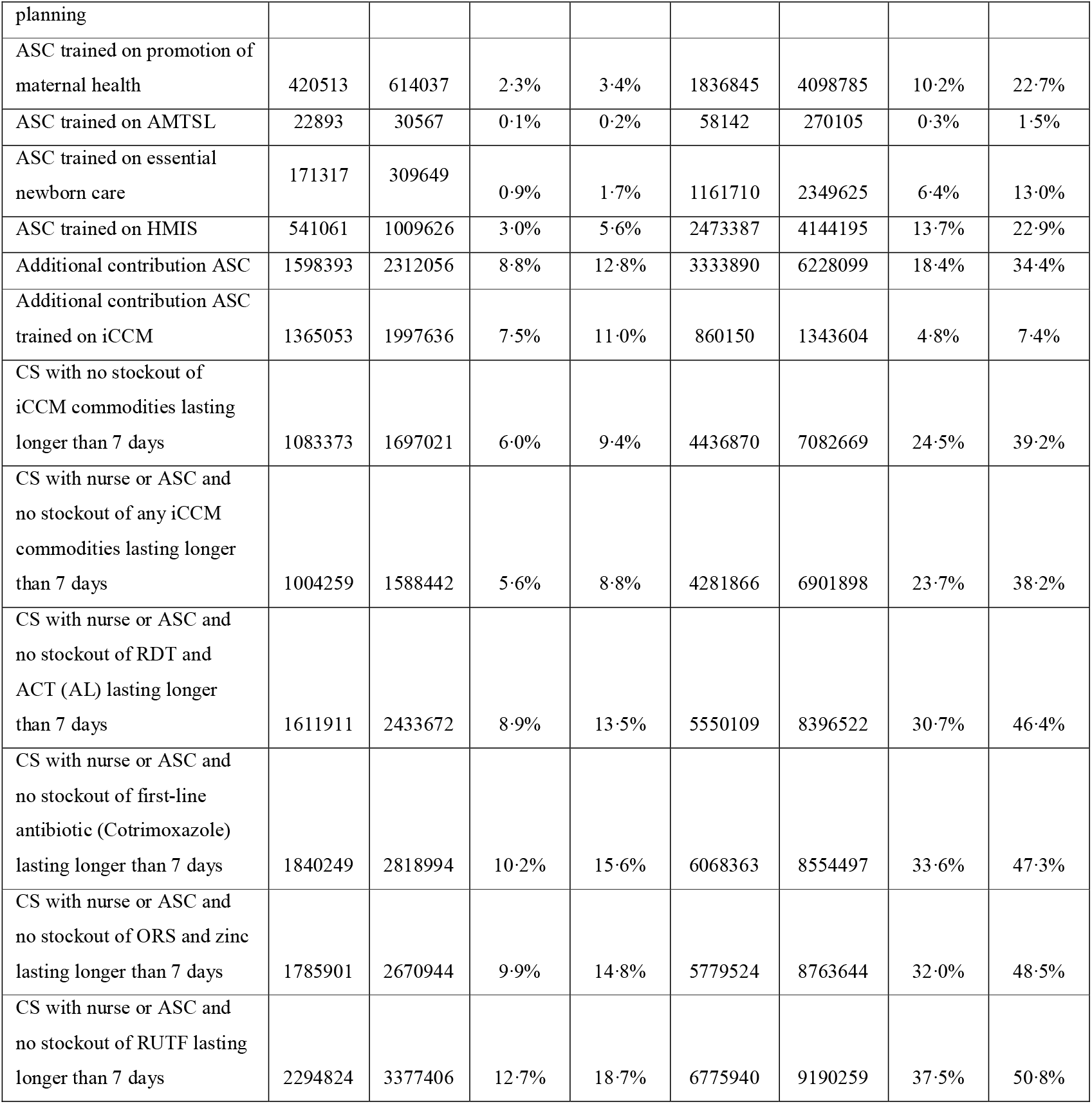
Accessibility coverage of the front-line health facility and ASC networks

**Figure 1.**
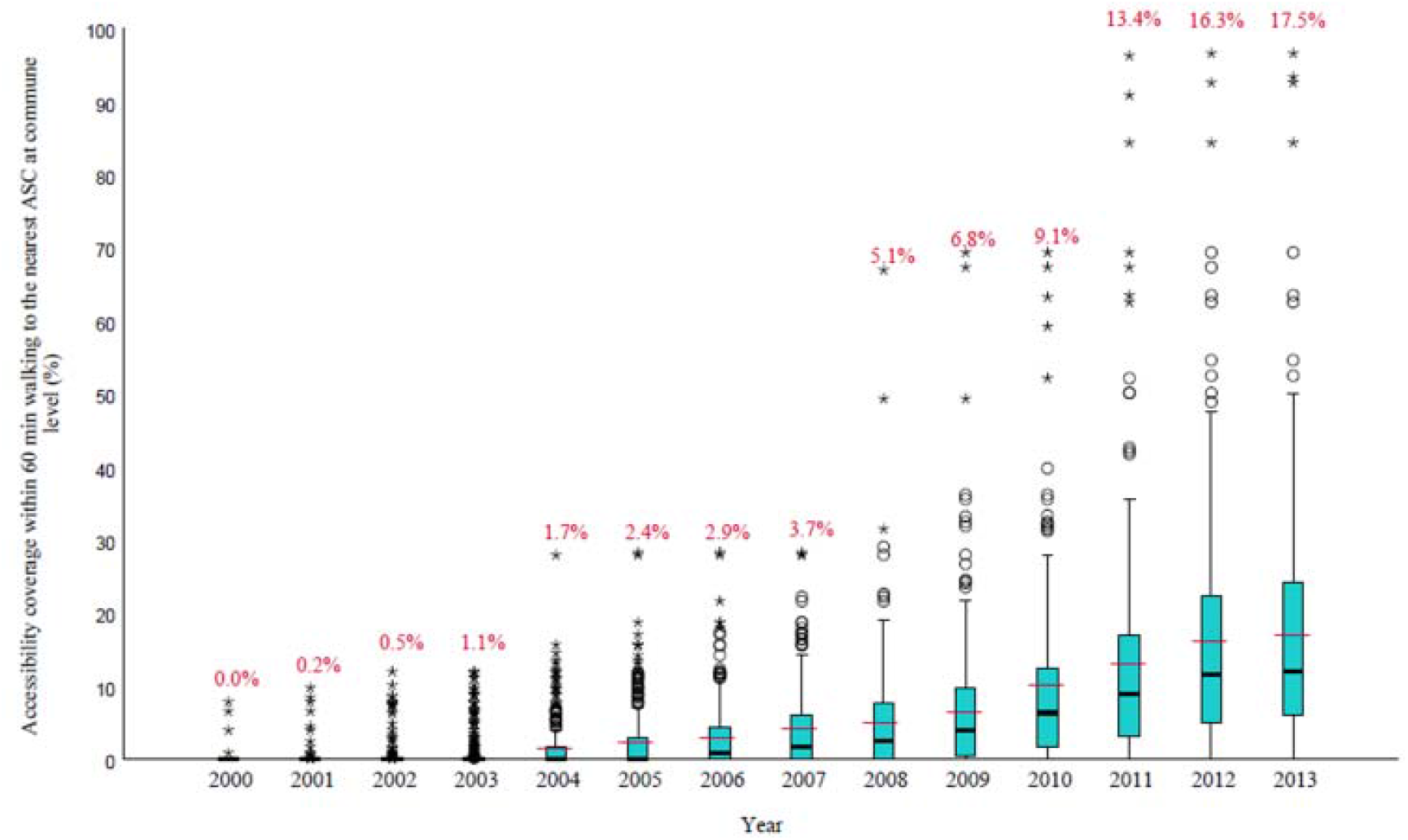
Median and interquartile range of the percent of the population within 60 minutes walking of an ASC at administrative level 3 (commune) between 2000-2013 at 100m x 100m resolution. Black lines indicate the median at commune level. Blue boxes represent the interquartile range at commune level. Circles and stars indicate communes outside of the interquartile range. Red lines and percentages indicate the national mean.

Accessibility coverage of the ASC network varied by gender of the ASC and training on specific interventions (Table 1, Supplementary Figure 2A-L on page 9 of Supplementary Appendix 1). Accessibility coverage of the ASC network trained on iCCM was 15·5% in 2013, given a 60-minute cutoff and walking scenario (Table 1, Figure 2D). The estimated additional contribution of the ASC network and ASC network trained on iCCM to accessibility coverage beyond the accessibility coverage of the existing CSI and CS (without ASC) networks combined, given a 60-minute cutoff and walking scenario, was 12·8% and 11·0%, covering an estimated 2·3 million and 2·0 million additional people, respectively (Table 1).

**Figure 2.**
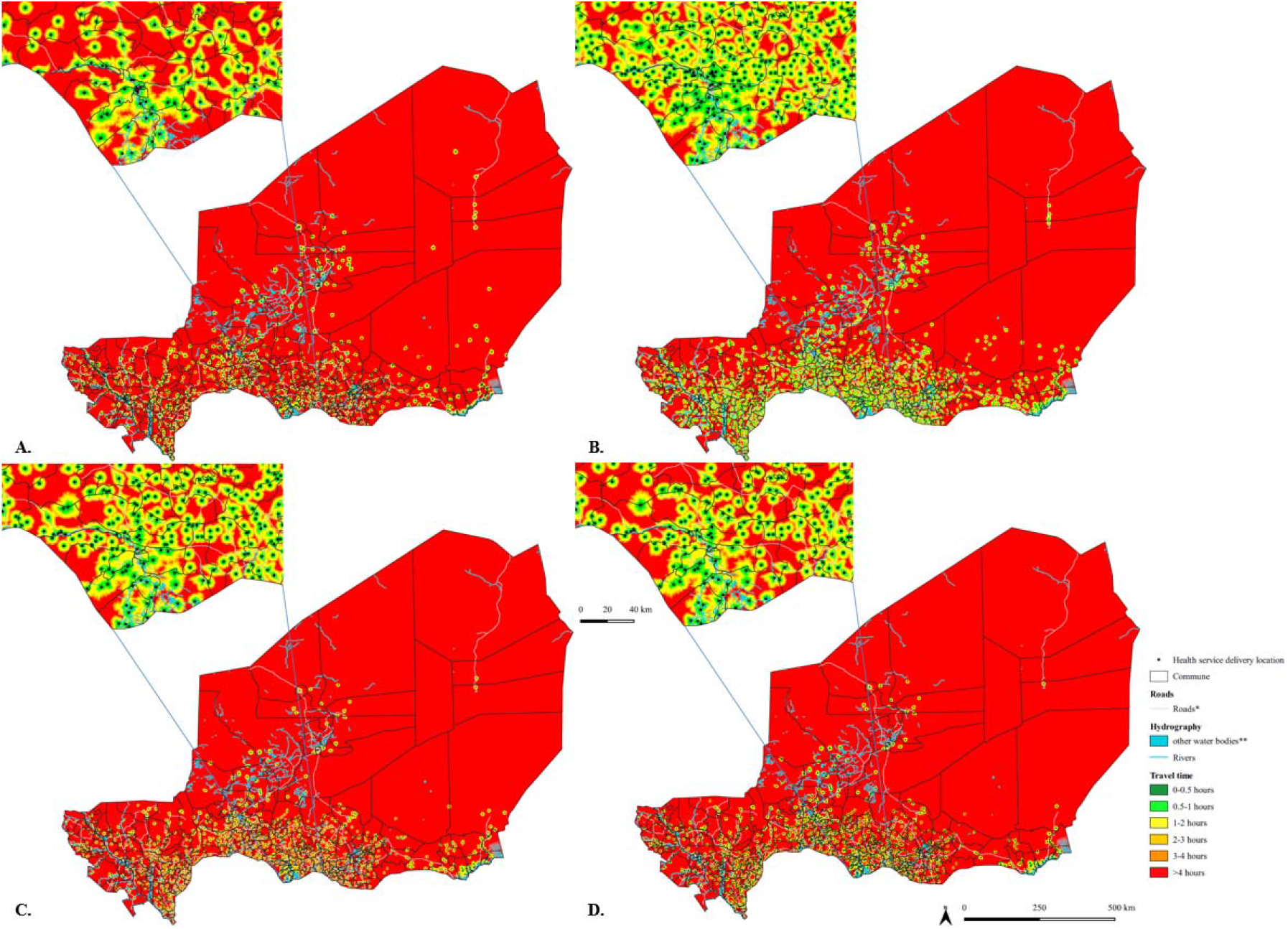
Geographic accessibility (travel time in minutes, walking in dry conditions) at 100m x 100m resolution. A) Centre de santé intégrée in 2013, n=839; B) Case de santé / Agent de santé communautaire in 2013, n=2550; C) Agent de santé communautaire in 2013, n=1457; D) Agent de santé communautaire in 2013 trained on iCCM, n=1214. Inset near Madarounfa commune in Maradi region. *For visualization purposes road classes limited to motorway, trunk, primary, secondary, and tertiary; **other water bodies from landcover layer included permanent water bodies, temporary water bodies and herbaceous wetlands.

Accessibility coverage in 2013, given a 60-minute cutoff and walking scenario, was 31·1% for the CSI network, 30·5% for the CS-ASC network, 21·4% for the CS network with a nurse or ASC, and 53·7% for the combined CSI + CS-ASC network (Table 1 and Figure 2A-D). An estimated 8·3 million people (58·2%) remained beyond 60 minutes walking to the nearest front-line health facility or ASC, without considering the maximum population capacity of these networks. Accessibility coverage of the CS network was lower when we considered availability of trained human resources (nurse or ASC) and essential commodities (Table 1 and Supplementary Figure 3A-G, page 7 of Supplementary Appendix 1). Accessibility coverage of all health service delivery networks was higher when considering the walking plus motorized transportation travel scenario (Table 1 and Supplementary Figure 4A-F, page 10 of Supplementary Appendix 1). We provide detailed results in Supplementary Appendix 2, tab “Detailed_Results”.

### Geographic coverage

Geographic coverage of the estimated total population in 2013 by the CSI network was 22·1%, assuming a walking scenario with a 60-minute catchment and maximum population capacity of 10000 per CSI (Figure 3 and Supplementary Appendix 3, tab “Summary”). Over one-third (347) of the CSI realized less than 30% of their maximum population capacity, indicating inefficiency (Supplementary Appendix 3, tab “Summary”).

**Figure 3.**
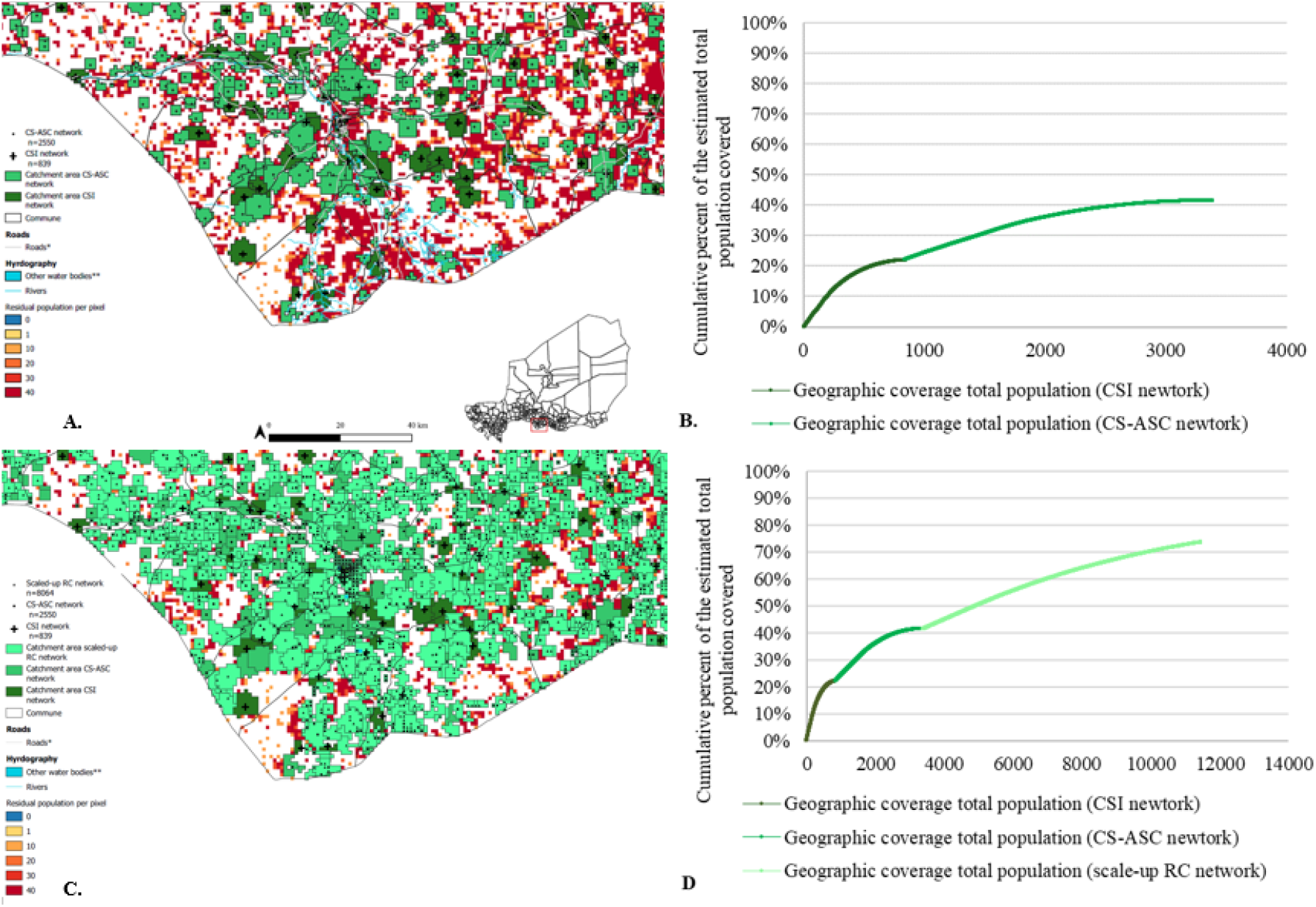
Geographic coverage of the CSI, CS-ASC, and hypothetical scale-up ASC networks in 2013, 60-minute catchment, walking scenario at 1km x 1km resolution. A) Geographic coverage at 1km x 1km resolution of the CSI (dark green) and CS-ASC networks (medium green) in 2013, 60-minute catchment (walking scenario) with inset near Madarounfa commune in Maradi region; B) Cumulative percent of the estimated total population covered within a 60-minute catchment, walking scenario (y-axis) by the number of CSI (x-axis, dark green line) and CS-ASC (x-axis, medium green line) at 1km x 1km resolution. C) Geographic coverage at 1km x 1km resolution of the CSI network (dark green), CS-ASC (medium green) and hypothetical scale-up RC network (light green) deployed to optimize geographic coverage of the residual population beyond the geographic coverage of the existing CSI and CS-ASC networks (60-minute catchment, walking scenario) in 2013, with maximum population capacity of 1000 people per RC, n=8064 RC, and inset near Madarounfa commune in Maradi region; D) Cumulative percent of the estimated total population covered within a 60-minute catchment, walking scenario (y-axis) by the number of CSI (x-axis, dark green), CS-ASC (x-axis, medium green), and hypothetical scale-up RC network (x-axis, light green) at 1km x 1km resolution. The hypothetical scale-up RC network targeted 1km x 1km grid cells with at least 250 people situated beyond the geographic coverage of the existing CSI and CS-ASC networks (60-minute catchment, walking scenario) in 2013. Maximum population capacity was set to 1000 people per RC. **Geographic coverage of the CSI and CS-ASC networks in 2013, 60-minute catchment, walking scenario at 1km x 1km resolution.** A) Geographic coverage at 1km x 1km resolution of the CSI and CS-ASC networks in 2013, 60-minute catchment (walking scenario) with inset near Madarounfa commune in Maradi region; B) Cumulative percent of the estimated total population covered within a 60-minute catchment, walking scenario (y-axis) by the number of CSI (x-axis, dark green line) and CS-ASC (x-axis, medium green line) at 1km x 1km resolution.

Geographic coverage of the total estimated population in 2013 by the CS-ASC network was 19·4%, assuming a walking scenario with a 60-minute catchment and maximum population capacity of 2500 per CS-ASC (Figure 3, Supplementary Figure 3). Over one-third (830) of the CS-ASC realized less than 30% of their maximum population capacity, indicating inefficiency (Figure 3 and Supplementary Appendix 3, tab “Summary”). Geographic coverage of the estimated residual population beyond the geographic coverage of the CSI network in 2013 by the CS-ASC network was 24·9%, providing an estimated 3·4 million additional people with physical access to PHC services, with important variation by region (Supplementary Appendix 3, tab “Summary” and Supplementary Figure 6).

### Geographic coverage of a hypothetical scale-up network of RC

A hypothetical network of 8064 RC with a maximum population capacity of 1000 people per RC, targeting cells with at least 250 people located beyond the geographic coverage of the existing CSI and CS-ASC networks, could cover 54·9% of this estimated residual population – providing physical access to PHC services for an estimated 5·7 million additional people in 2013 (Figure 5 and Supplementary Appendix 6, tab “Summary”). Geographic coverage of the estimated total population would increase from 41·5% covered by the existing CSI and CS-ASC networks to 73·6% by the combined CSI, CS-ASC, and hypothetical RC networks in 2013 (Supplementary Appendix 4, tab “Summary”).

### Efficiency of geographical targeting

Geographic coverage of the estimated residual population beyond the geographic coverage of the existing CSI network was 40.2% by the hypothetical CS-ASC network compared to 24·9% by the existing CS-ASC network, covering an estimated 2·1 million additional people – a 61.9% gain in efficiency (Figure 4 and Supplementary Appendix 5, tab “Comparison_Population”). Geographic coverage of the estimated residual under-five deaths beyond the geographic coverage of the existing CSI network was 58·0% by the hypothetical CS-ASC network compared to 37·5% by the existing CS-ASC network, covering an estimated 73000 under-five deaths not otherwise covered – a 55·0% gain in efficiency (Figure 4 and Supplementary Appendix 5, tab “Comparison_U5deaths”). Geographic coverage of the estimated residual *Pf* malaria cases (all ages) beyond the geographic coverage of the existing CSI network was 60·3% by the hypothetical CS-ASC network compared to 33·1% by the existing CS-ASC network, covering an estimated 1·6 million *Pf* malaria cases not otherwise covered – a 81·9% gain in efficiency (Figure 4 and Supplementary Appendix 5, tab “Comparison_Malaria”). Our uncertainty analysis for the efficiency of geographical targeting indicates bins/groups of CS-ASC catchments with relatively higher efficiency geographical targeting could be distinguished from bins/groups of CS-ASC catchments with relatively lower efficiency of geographical targeting (Supplementary Appendix 6).

**Figure 4.**
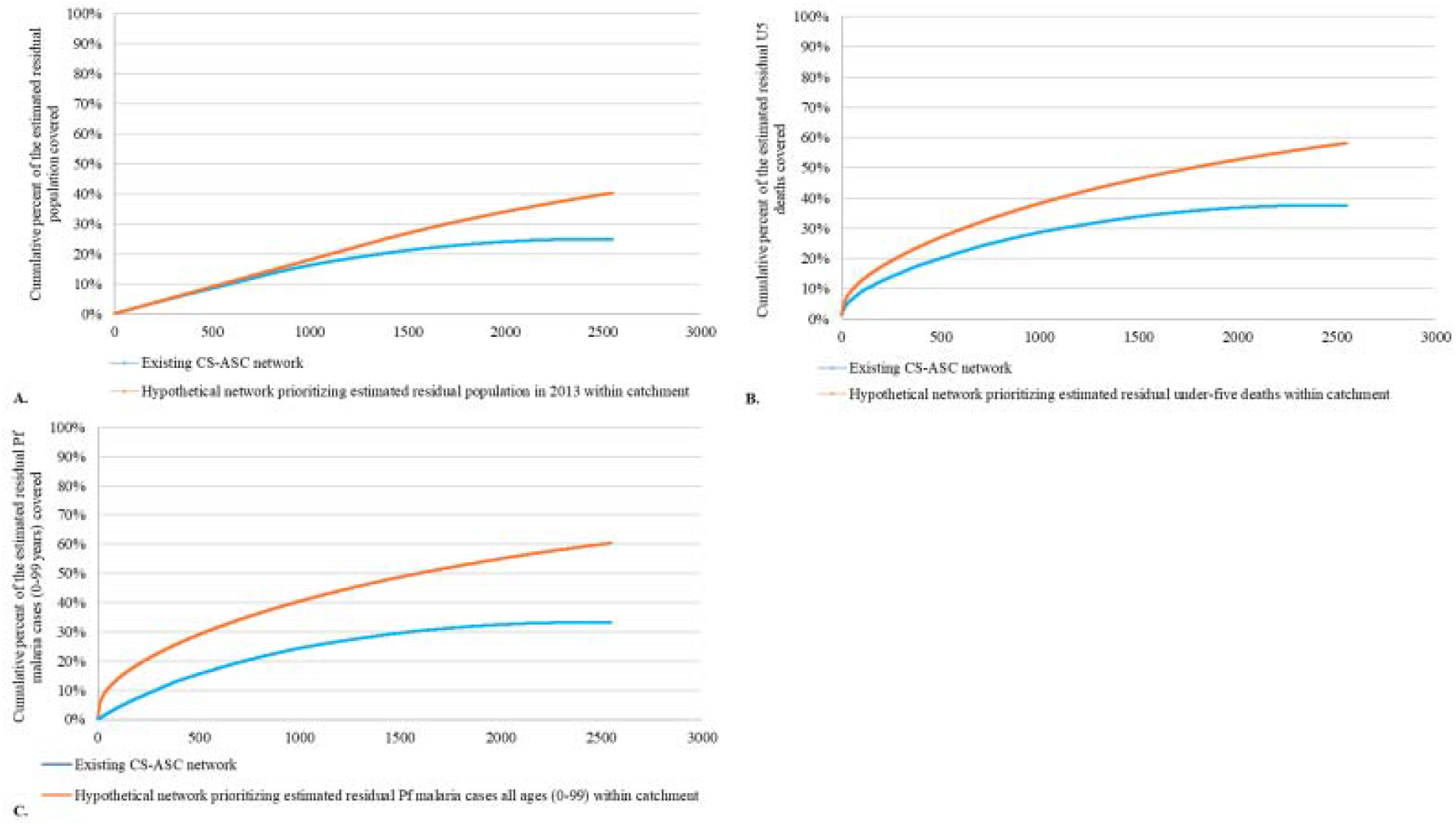
Targeting of the existing CS-ASC network compared to hypothetical optimized networks at 1km x 1km resolution. A) Comparison of the percent of the estimated residual population beyond the geographic coverage of the existing CSI network (60-minute catchment, walking scenario) that was covered by the existing CS-ASC network compared to a hypothetical CS-ASC network deployed to optimize geographic coverage of the estimated residual population; B) Comparison of the percent of the estimated residual under-five deaths beyond the geographic coverage of the existing CSI network (60-minute catchment, walking scenario) that was covered by the existing CS-ASC network compared to a hypothetical CS-ASC network deployed to optimize geographic coverage of the estimated residual under-five deaths; C) Comparison of the percent of the estimated residual Pf malaria cases among all ages (0-99 years) beyond the geographic coverage of the existing CSI network (60-minute catchment, walking scenario) that was covered by the existing CS-ASC network compared to a hypothetical CS-ASC network deployed to optimize geographic coverage of the estimated residual Pf malaria cases among all ages (0-99 years). All analyses at 1km x 1km resolution.

## DISCUSSION

### Summary

Our results indicate that geographical accessibility of PHC services improved in Niger between 2000-2013, with an important contribution of ASCs – making PHC services physically accessible for an estimated 2·3 million additional people, including an estimated 2·0 million additional people with physical access to iCCM. Despite this improvement, geographical accessibility of PHC services remained inadequate as of 2013, with important variation across subnational geographies, training of CHWs, and availability of essential commodities. Rational scale-up of the CHW network, using approaches to optimize deployment as described here, could increase geographic coverage of PHC services from 41·5% to 73·6%, covering an estimated 5·7 million additional people without adequate physical access to PHC services in 2013. The efficiency of geographical targeting of health facilities and CHWs was suboptimal relative to the distribution of the estimated population, under-five deaths and malaria burden – but could be improved through the targeting approach used in this study.

### Strengths and limitations

To our knowledge, this study provides the first estimates and high-resolution maps of geographical accessibility to CHWs at national scale for any country. We used a bespoke modelling approach with a complete national georeferenced census of front-line health facilities and CHWs in Niger, other high resolution spatial datasets, and realistic assumptions.

Our estimates of accessibility coverage are lower than those from Weiss and colleagues^34^ which estimated 61·8% and 80·7% of the population in Niger was within 60 minutes of the nearest health facility in 2020 based on a walking scenario and motorized transportation scenario, respectively. Paradoxically the health facility dataset used in Weiss and colleagues^34^ included 568 fewer health service delivery locations (n=2821 in Weiss and colleagues^34^ for 2020, n=3389 in our model for 2013). The higher estimates of accessibility coverage in Weiss and colleagues^34^ may be explained by three factors. First, travel speeds for motorized transportation used by Weiss and colleagues^34^ are higher than those used in our model. Second, the walking model by Weiss and colleagues^34^ assumes a travel speed of 5km per hour across all landcover classes, whereas our model varies walking travel speed from 5km per hour to 0 km per hour (impassable), according to land cover class and is based on realistic estimates of travel speeds from previous studies in Niger and the region.^29,30^ Third, Weiss and colleagues^34^ allowed for swimming across water barriers, whereas we did not but we allowed travel over road bridges. Another study, Blanford and colleagues,^30^ found that 39·1% of the population were within 60 minutes walking of the nearest health facility in 2009. However, their model used an incomplete network of hospitals and CSI (n=504) dating from 1995. We consider our results, while imperfect, to be more useful for national planning in Niger than results reported in Weiss and colleagues^34^ and Blanford and colleagues^31^ as our results were based on a bespoke model for Niger, with a more complete dataset on health service delivery locations and more realistic, contextualized travel speeds.

Our scale-up analysis provides a useful roadmap for the MOPH to consider as it plans to scale-up the RC network to fill gaps in physical access to PHC services, for example, by identifying optimal RC locations and enabling the prioritization of bins/groups of RC (the first 500 RC, next 500 RC and so on) that would maximize geographical accessibility, allowing a phased approach to scale-up. Lastly, our analysis highlights important inefficiencies in geographical targeting of the existing CS-ASC network relative to estimates of the spatial distribution of the population and underlying burden of under-five mortality and *Pf* malaria, suggests alternatives to guide fine-tuning of the network, and provides an approach that could be used in future planning efforts to maximize efficiency of geographical targeting.

We acknowledge that, in addition to physical accessibility, it is important to consider social and economic barriers to care seeking (e.g. social norms, intrahousehold power dynamics, costs of transportation, opportunity costs of travel time, costs of services and commodities) which may influence access to and use of health services.^35^ It is also important to consider the quality of health services and the potential for bypassing.^36,37^ Lastly, predominate modes of transportation may vary by socioeconomic status and geography^38^ and they may change in response to contextual factors (e.g. the lockdowns due to COVID-19 in 2020).

There are important limitations to this study. First, we did not include secondary or tertiary facilities or outreach/mobile sites. We focused on the question of physical access to CHWs and first level health facilities, rather than secondary or tertiary health facilities and permanent, fixed service locations rather than periodic, mobile services. Moreover, our service delivery location dataset is more complete than datasets publicly available at the time of our analysis in 2020.^39^ Second, our analysis is limited by the completeness and quality of the publicly available data on road and river networks. We acknowledge that more complete and/or accurate government or proprietary road and river network data may be available. For the river network, we acknowledge that some rivers, streams, and other waterways may not be perennial barriers to movement. We attempted to mitigate this limitation by allowing major road classes (motorway, trunk, primary, secondary, and tertiary) to cross rivers/streams and by incorporating data on the hydrographic network from the high-resolution Copernicus land cover layer^15^ in our merged land cover layer. We also conducted a sensitivity analysis using only waterways classified as “rivers” in the rivers input layer as barriers to movement and found this made no important difference to the results (Supplementary Appendix 2, tab Sensitivity_analysis”). Third, our accessibility coverage, geographic coverage, and targeting analyses do not account for uncertainty of the estimates of population. Previous analyses of accessibility coverage and geographic coverage have not uncounted for uncertainty of this kind, but we acknowledge this is an important limitation and area for improving future modelling. Fourth, our analysis does not account for national parks or other “no-go” zones (e.g., military bases) due to lack of access to the geography of these objects for 2013. Fifth, our travel speeds were based on similar analyses for Niger and other countries in sub-Saharan Africa^29,30^ but could be improved through a country-led consensus-building process with national experts to derive subnational travel speeds and validate other data inputs. Our travel speeds do not account for differences in travel speeds by season^30^ which may be relevant in the Sahel, possible group differences (e.g., pregnant women, people with illness, and caregivers carrying sick children may walk slower), river transportation, and our walking plus motorized transportation scenario assumes immediate access to a vehicle once a road is reached and does not account for road traffic or factors impacting road traffic (e.g. traffic lights). Sixth, our analysis does not account for the possibility of accessing health service delivery locations across national boundaries, an important consideration for cross-border and migrant populations. Seventh, the modelled population counts for 2000-2012 use the HRSL population settlement footprint from 2015,^26^ which may not accurately reflect the population settlement footprint for the early 2000s. Lastly, the accuracy of the modelled estimates of under-five mortality rates^27^ and *Pf* malaria incidence^28^ used in our targeting analysis is unknown. Despite this limitation, results from our uncertainty analysis indicated that our targeting approach could be used to confidently identify bins/groups of health service delivery catchment areas that are relatively more efficient at geographical targeting than other bins/groups – and that this information could be used in planning to enhance scale-up and geographical targeting of CHWs.

We understand that rational decisions on scale-up and targeting of CHWs, like with health facilities, cannot be addressed purely through modelling, as there are many factors involved in the political economy of health system planning and decision-making.^40,41^ Nonetheless, we think our results provide novel insights for policy makers, practitioners and researchers and future efforts can build on and refine this work through collaborative, country-led processes.

## CONCLUSION

Geographical accessibility of PHC services improved in Niger between 2000-2013, with important contributions from ASCs, providing an estimated 2·3 million additional people with physical access to PHC services – including 2·0 million additional people with physical access to iCCM. However, as of 2013, gaps in geographical accessibility remained and efficiency of geographical targeting of ASCs was suboptimal. Rational scale-up and geographical targeting of the RC network, using approaches to optimize deployment as described here, could make an important contribution toward increasing geographical accessibility to PHC services in Niger. These approaches could be adapted to meet planning needs in other contexts. As Niger works toward UHC, developing a geography of the health system – as part of a broader spatial data infrastructure^42^ – will be critical. Developing a geography of CHWs within this broader context will be equally important for these purposes and for maximizing efficiency, impact, and equity of investments in quality PHC within the context of UHC. This study is one step toward a geography of CHWs in Niger and a call to action for establishing a geography of CHWs globally.

## Data Availability

Data are available on a public, open access repository under the Creative Commons Attribution 4.0 Unported (CC BY 4.0) license, which permits others to copy, redistribute, remix, transform and build upon this work for any purpose, provided the original work is properly cited, a link to the license is given, and indication of whether changes were made. See: https://creativecommons.org/licenses/by/4.0/. Supplementary Appendices 2-6, Videos 1-2, and all model outputs are available in Supplementary Appendix 1b at https://doi.org/10.5281/zenodo.4482969. All model inputs (except existing service delivery locations) are available in Supplementary Appendix 1c at https://doi.org/10.6084/m9.figshare.13536779.v5. Health service delivery location data are only available through data sharing agreements with UNICEF and the Ministry of Public Health of Niger.

https://doi.org/10.6084/m9.figshare.13536779.v5

## ACKNOWLEDGEMENTS

The views expressed in this article are the authors’ views do not necessarily represent the views, positions, or policies of the institutions with which the authors are affiliated. This work would not have been possible without the efforts of the many people that contributed to the first georeferenced census of CSI, CS and ASC led by the INS, the Ministry of Public Health of Niger, and UNICEF in 2013.

## FOOTNOTES

### Contributors

NPO was responsible for the study conceptualization, methodology, data curation, and writing the draft manuscript. OH, IM, KB, AYG, NPO, and NR collected data or provided feedback on data. NPO, NR and ZY conducted the formal analysis and were responsible for data visualization. NPO, NR and TD verified the underlying data. TD, DJ, and NR provided supervision and overall guidance. All authors contributed to reviewing and editing the manuscript.

### Patient consent for publication

Not required.

### Provenance and peer review

Not commissioned; externally peer reviewed.

