## Supplementary figures and images for "Toward a geography of community health workers in Niger: a geospatial analysis"

### Video 1

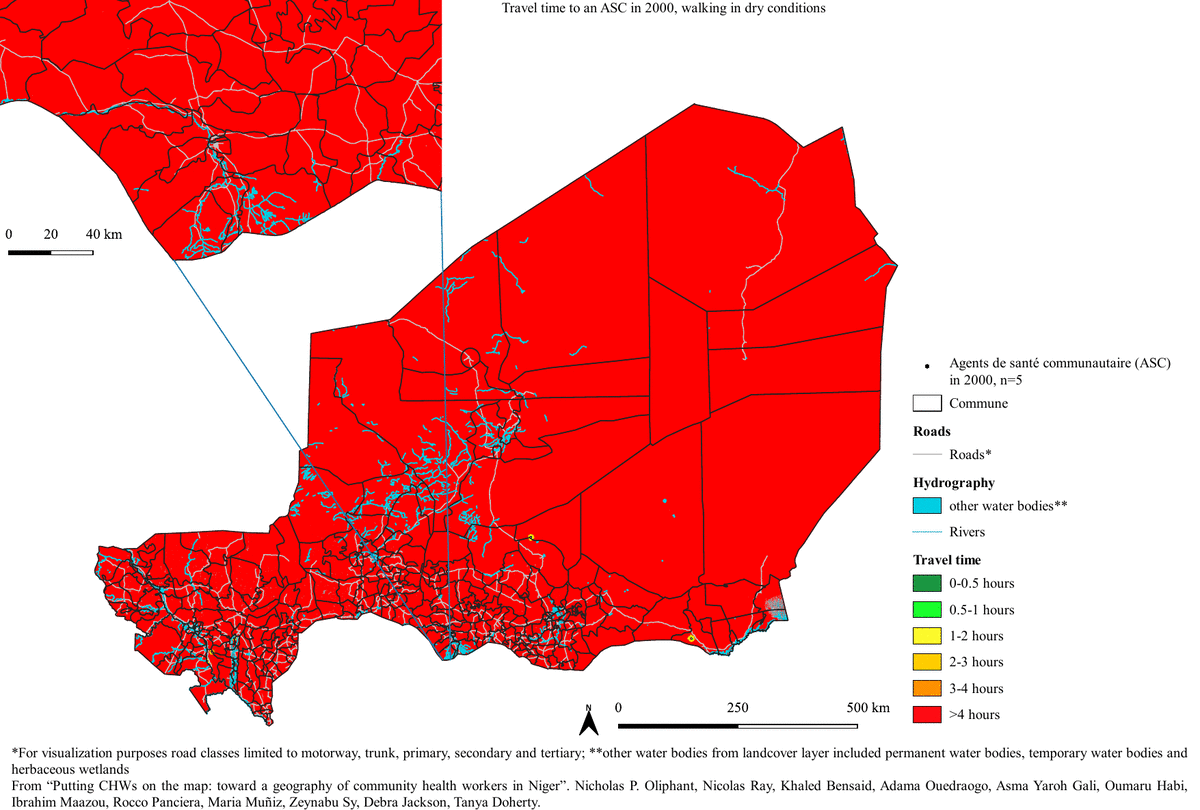

### Video 2

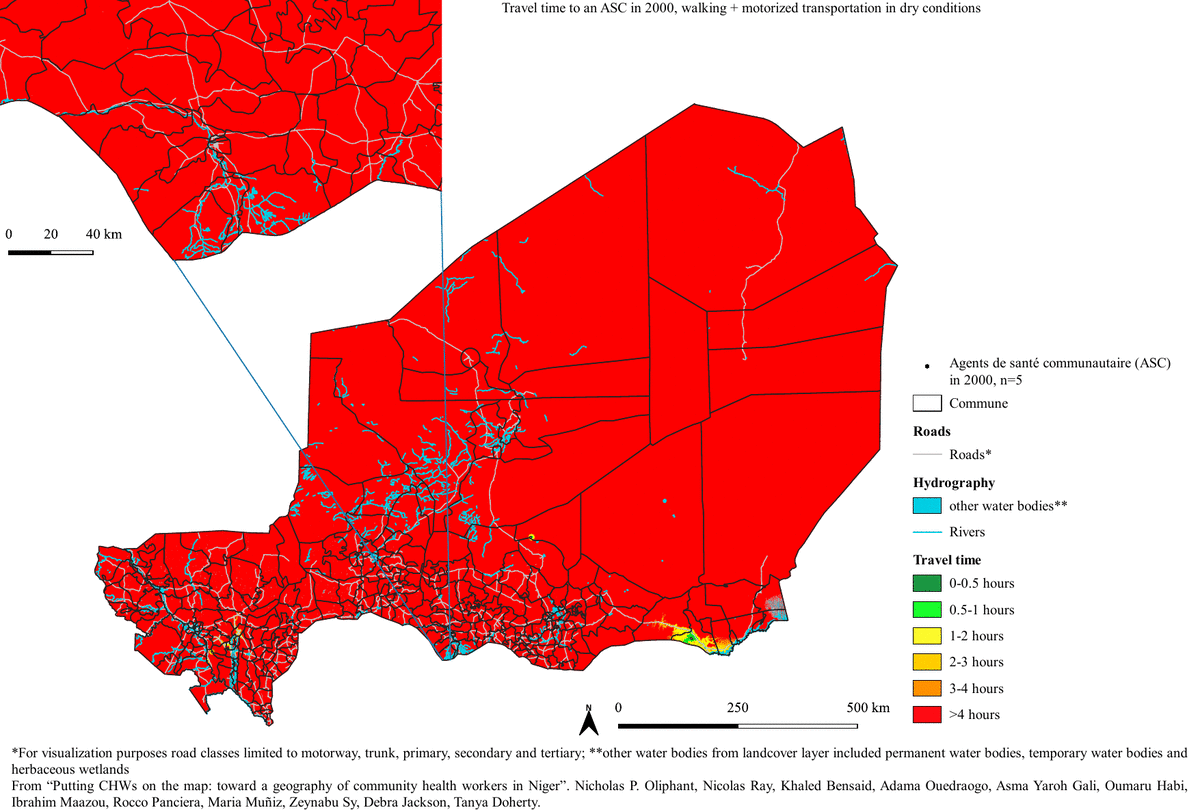
