## Supplementary Appendix 1 for "Toward a geography of community health workers in Niger: a geospatial analysis"

**Supplementary Appendix 1a**

This file provides supplementary figures, tables, and methods for “Toward a geography of community health workers in Niger: a geospatial analysis” by Nicholas P Oliphant, Nicolas Ray, Khaled Bensaid, Adama Ouedraogo, Asma Yaroh Gali, Oumaru Habi, Ibrahim Maazou, Rocco Panciera, Maria Muñiz, Samuel OM Manda, Zeynabou Sy, Debra Jackson, and Tanya Doherty.

**Table of Contents**

Supplementary Figure 1. Simplified analysis flow diagram	5

Supplementary Figure 2 (A). Geographic accessibility (travel time in minutes, walking in dry conditions) to the nearest female ASC in 2013	8

Supplementary Figure 2 (B). Geographic accessibility (travel time in minutes, walking in dry conditions) to the nearest male ASC in 2013	8

Supplementary Figure 2 (C). Geographic accessibility (travel time in minutes, walking in dry conditions) to the nearest ASC in 2013 trained on promotion of key family practices	9

Supplementary Figure 2 (D). Geographic accessibility (travel time in minutes, walking in dry conditions) to the nearest ASC in 2013 trained on promotion of WASH	9

Supplementary Figure 2 (E). Geographic accessibility (travel time in minutes, walking in dry conditions) to the nearest ASC in 2013 trained on promotion of maternal and child nutrition	10

Supplementary Figure 2 (F). Geographic accessibility (travel time in minutes, walking in dry conditions) to the nearest ASC in 2013 trained on promotion of childhood immunization	11

Supplementary Figure 2 (G). Geographic accessibility (travel time in minutes, walking in dry conditions) to the nearest ASC in 2013 trained on prevention of HIV and STI	11

Supplementary Figure 2 (H). Geographic accessibility (travel time in minutes, walking in dry conditions) to the nearest ASC in 2013 trained on family planning	12

Supplementary Figure 2 (I). Geographic accessibility (travel time in minutes, walking in dry conditions) to the nearest ASC in 2013 trained on promotion of maternal health	12

Supplementary Figure 2 (J). Geographic accessibility (travel time in minutes, walking in dry conditions) to the nearest ASC in 2013 trained on AMTL	13

Supplementary Figure 2 (K). Geographic accessibility (travel time in minutes, walking in dry conditions) to the nearest ASC in 2013 trained on essential newborn care	14

Supplementary Figure 2 (L). Geographic accessibility (travel time in minutes, walking in dry conditions) to the nearest ASC in 2013 trained on the HMIS	14

Supplementary Figure 3 (A). Geographic accessibility (travel time in minutes, walking in dry conditions) to the nearest CS in 2013 with a nurse or ASC	15

Supplementary Figure 3 (B). Geographic accessibility (travel time in minutes, walking in dry conditions) to the nearest CS in 2013 with no severe stockout of any iCCM commodities	15

Supplementary Figure 3 (C). Geographic accessibility (travel time in minutes, walking in dry conditions) to the nearest CS in 2013 with a nurse or ASC and no severe stockout of any iCCM commodities	16

Supplementary Figure 3 (D). Geographic accessibility (travel time in minutes, walking in dry conditions) to the nearest CS in 2013 with a nurse or ASC and no severe stockout of RDT or AL	17

Supplementary Figure 3 (E). Geographic accessibility (travel time in minutes, walking in dry conditions) to the nearest CS in 2013 with a nurse or ASC and no severe stockout of ORS or zinc	17

Supplementary Figure 3 (F). Geographic accessibility (travel time in minutes, walking in dry conditions) to the nearest CS in 2013 with a nurse or ASC and no severe stockout of cotrimoxazole	18

Supplementary Figure 3 (G). Geographic accessibility (travel time in minutes, walking in dry conditions) to the nearest CS in 2013 with a nurse or ASC and no severe stockout of RUTF	19

Supplementary Figure 4. (A) Geographic accessibility (travel time in minutes, walking + motorized transportation in dry conditions) to the nearest CSI in 2013	19

Supplementary Figure 4. (B) Geographic accessibility (travel time in minutes, walking + motorized transportation in dry conditions) to the nearest CS/ASC in 2013	20

Supplementary Figure 4. (C) Geographic accessibility (travel time in minutes, walking + motorized transportation in dry conditions) to the nearest ASC in 2013	20

Supplementary Figure 4. (D) Geographic accessibility (travel time in minutes, walking + motorized transportation in dry conditions) to the nearest ASC in 2013 trained on iCCM	21

Supplementary Figure 4. (E) Geographic accessibility (travel time in minutes, walking + motorized transportation in dry conditions) to the nearest female ASC in 2013	22

Supplementary Figure 4. (F) Geographic accessibility (travel time in minutes, walking + motorized transportation in dry conditions) to the nearest male ASC in 2013	22

Supplementary Figure 5 (A). Contribution of ASC to additional geographic accessibility beyond the existing CSI and CS (without ASC) networks in 2013 (walking scenario)	23

Supplementary Figure 5 (B). Contribution of ASC to additional geographic accessibility beyond the existing CSI and CS (without ASC) networks in 2013 (walking + motorized transportation scenario)	23

Supplementary Figure 5 (C). Contribution of ASC trained on iCCM to additional geographic accessibility beyond the existing CSI and CS (without ASC) networks in 2013 (walking scenario)	24

Supplementary Figure 5 (D). Contribution of ASC trained on iCCM to additional geographic accessibility beyond the existing CSI and CS (without ASC) networks in 2013 (walking + motorized transportation scenario)	25

Supplementary Figure 6. Median and interquartile range of geographic coverage at commune level (administrative level 3) of the residual population beyond the geographic coverage of the CSI network that were covered by the existing CS-ASC network, by region (administrative level 1**)**	25

Supplementary Figure 7. Median and interquartile range of geographic coverage at commune level (administrative level 3) of the residual population beyond the geographic coverage of the CSI network that were covered by the hypothetical CS-ASC network deployed to optimize geographic coverage of the residual population, by region (administrative level 1)	26

Supplementary Figure 8. Targeting with saturation	27

Supplementary Figure 9. Digital elevation model at 100m	28

Supplementary Figure 10. Estimated population count in 2013 (persons per grid cell) at 1km x 1km resolution	29

Supplementary Figure 11. Mean U5 deaths in 2013 at 1km x 1km	30

Supplementary Figure 12. Estimated *Pf* malaria cases among all ages (0-99 years) per grid cell at 1km x 1km resolution	31

Supplementary Figure 13. Road network	32

Supplementary Figure 14. Rivers	32

Supplementary Figure 15. Other water bodies	33

Supplementary Figure 16. Land cover	33

Supplementary Figure 17. Merged land cover at 100m x 100m resolution	34

Supplementary Figure 18. Health service delivery locations	35

Data	35

Administrative boundaries	35

Health system pyramid and health service delivery networks mapped	35

Supplementary Figure 19. Health system pyramid and health service delivery networks mapped	35

DEM	38

Land cover	38

Roads	39

Rivers and other water bodies	39

Merged land cover	39

Travel scenario tables	40

Population	40

Estimated under-five mortality	43

Estimated *Plasmodium falciparum* malaria cases	44

Analysis	44

Geographic accessibility	44

Geographic coverage	49

Targeting	53

References	65

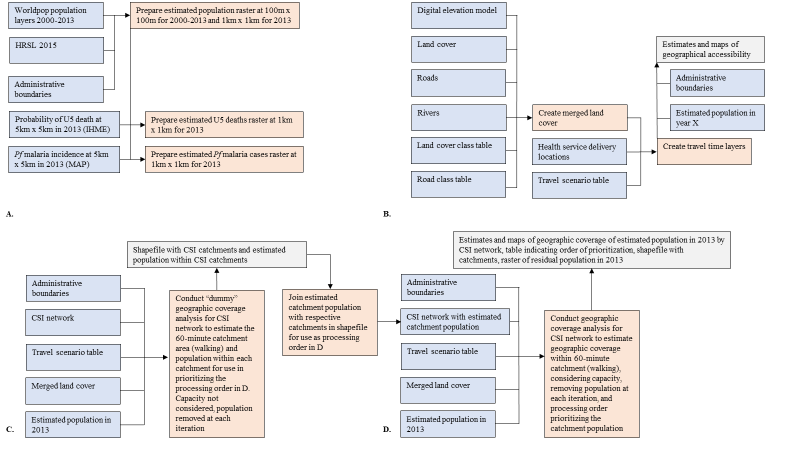

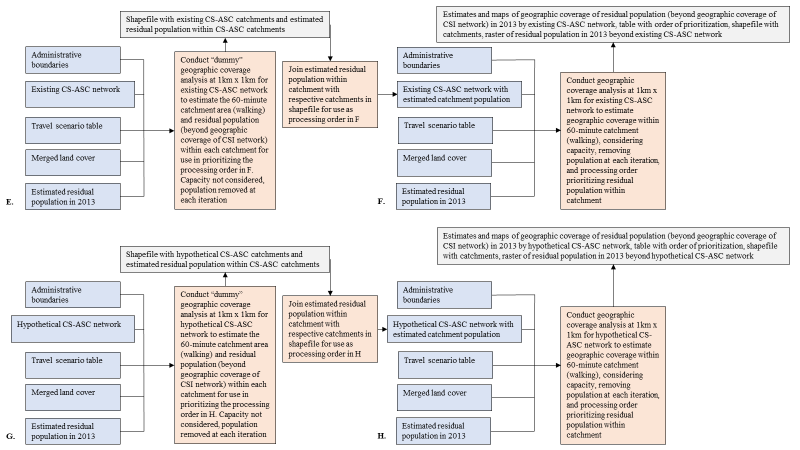

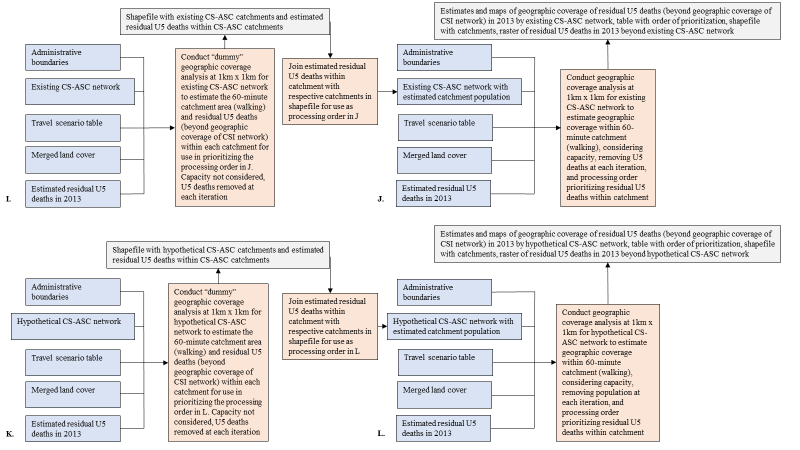

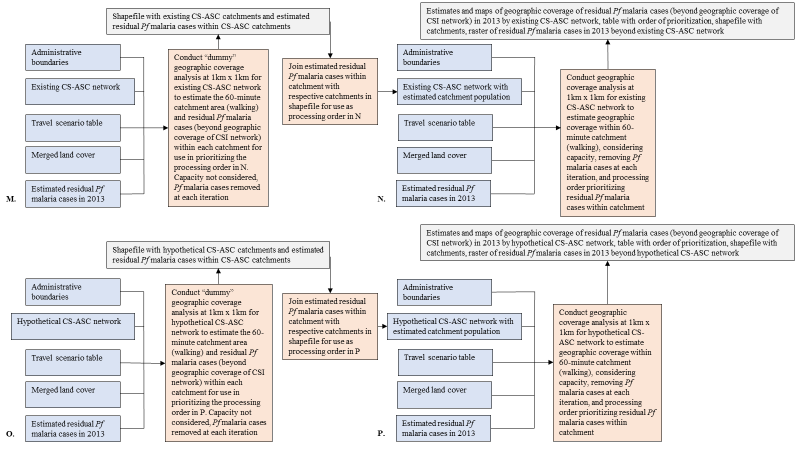

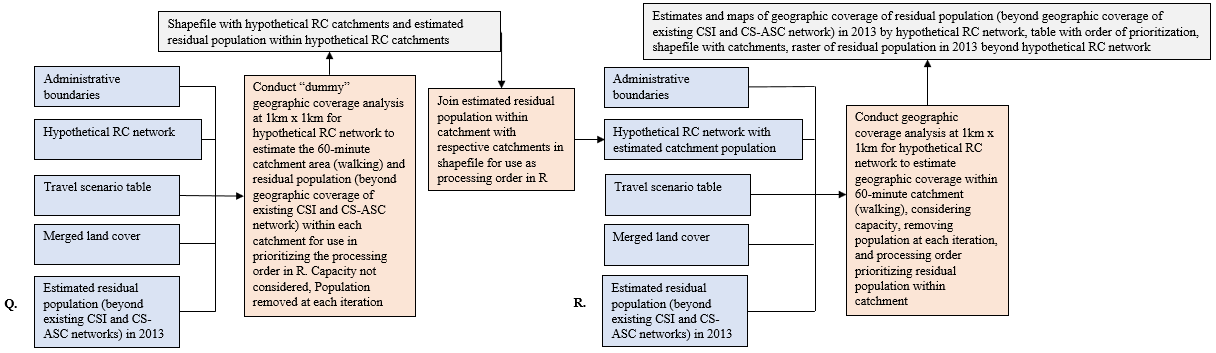

### **Supplementary Figure 1. Simplified analysis flow diagram**

Analysis flow for preparation of estimated population layers 2000-2013, estimated U5 deaths layer, and estimated Pf malaria cases layer. (B) Analysis flow for estimates and maps of geographical accessibility. (C) Analysis flow for “dummy” geographic coverage of the population in 2013 by the CSI network at 1km x 1km resolution in preparation for (D). (D) Analysis flow for estimates and maps of geographic coverage of the population in 2013 by the CSI network at 1km x 1km resolution. (E) Analysis flow for “dummy” geographic coverage of the estimated residual population (beyond geographic coverage of the CSI network) in 2013 by the existing CS-ASC network at 1km x 1km resolution in preparation for (F). (F) Analysis flow for estimates and maps of geographic coverage of the estimated residual population (beyond geographic coverage of the CSI network) in 2013 by existing CS-ASC network at 1km x 1km resolution. (G) Analysis flow for “dummy” geographic coverage of the estimated residual population (beyond geographic coverage of the CSI network) in 2013 by the hypothetical CS-ASC network at 1km x 1km resolution in preparation for (H). (H) Analysis flow for estimates and maps of geographic coverage of the estimated residual population (beyond geographic coverage of the CSI network) in 2013 by a hypothetical CS-ASC network deployed to optimize geographic coverage of the estimated residual population at 1km x 1km resolution. (I) Analysis flow for “dummy” geographic coverage at 1km x 1km resolution of the estimated residual under-five deaths (beyond geographic coverage of the CSI network) in 2013 by the existing CS-ASC network in preparation for (J). (J) Analysis flow for estimates and maps of geographic coverage of the estimated residual under-five deaths (beyond geographic coverage of the CSI network) in 2013 by existing CS-ASC network at 1km x 1km resolution. (K) Analysis flow for “dummy” geographic coverage of the estimated residual under-five deaths (beyond geographic coverage of the CSI network) in 2013 by the hypothetical CS-ASC network at 1km x 1km resolution in preparation for (L). (L) Analysis flow for estimates and maps of geographic coverage of the estimated residual under-five deaths (beyond geographic coverage of the CSI network) in 2013 by a hypothetical CS-ASC network deployed to optimize geographic coverage of the residual under-five deaths at 1km x 1km resolution. (M) Analysis flow for “dummy” geographic coverage of the estimated residual Pf malaria cases (beyond geographic coverage of the CSI network) in 2013 by the existing CS-ASC network at 1km x 1km resolution in preparation for (N). (N) Analysis flow for estimates and maps of geographic coverage of the estimated residual Pf malaria cases (beyond geographic coverage of the CSI network) in 2013 by the existing CS-ASC network at 1km x 1km resolution. (O) Analysis flow for “dummy” geographic coverage of the estimated residual Pf malaria cases (beyond geographic coverage of the CSI network) in 2013 by the hypothetical CS-ASC network at 1km x 1km resolution in preparation for (P). (P) Analysis flow for estimates and maps of geographic coverage of the estimated residual Pf malaria cases (beyond geographic coverage of the CSI network) in 2013 by a hypothetical CS-ASC network deployed to optimize geographic coverage of the residual Pf malaria cases at 1km x 1km resolution. (Q) Analysis flow for “dummy” geographic coverage of the estimated residual population malaria (beyond geographic coverage of the existing CSI and CS-ASC networks) in 2013 by a hypothetical scaled-up network of Relais Communautaire (RC) at 1km x 1km resolution in preparation for (R). (R) Analysis flow for estimates and maps of geographic coverage of the estimated residual population (beyond geographic coverage of the existing CSI and CS-ASC networks) in 2013 by a hypothetical scaled-up network of RC at 1km x 1km resolution. Blue boxes represent data inputs. Orange boxes represent analysis steps. Grey boxes represent outputs. HRSL = High Resolution Settlement Layer. IHME = Institute for Health Metrics and Evaluation. MAP = Malaria Atlas Project. Pf malaria = Plasmodium falciparum. U5 = under-five.

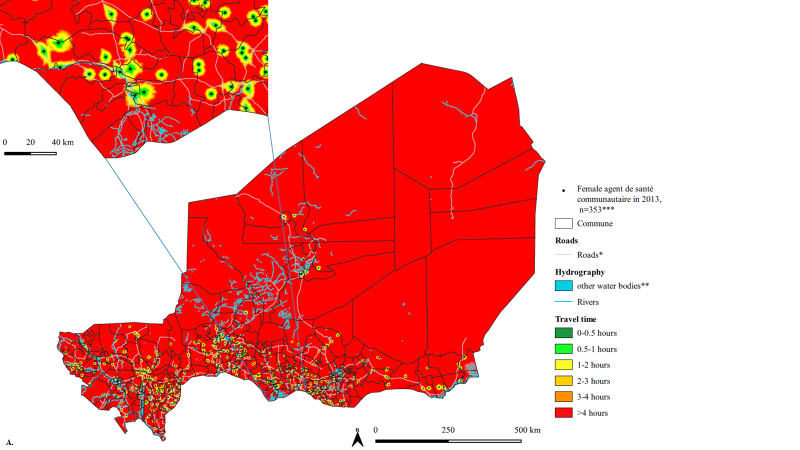

**Supplementary Figure 2 (A). Geographic accessibility (travel time in minutes, walking in dry conditions) to the nearest female ASC in 2013 at 100m x 100m resolution.**
Female ASC in 2013, n=353. *For visualization purposes road classes limited to motorway, trunk, primary, secondary, and tertiary. **Other water bodies from landcover layer included permanent water bodies, temporary water bodies and herbaceous wetlands. ***Gender for 2 ASC was not recorded, and these ASC were excluded from the gender analysis.

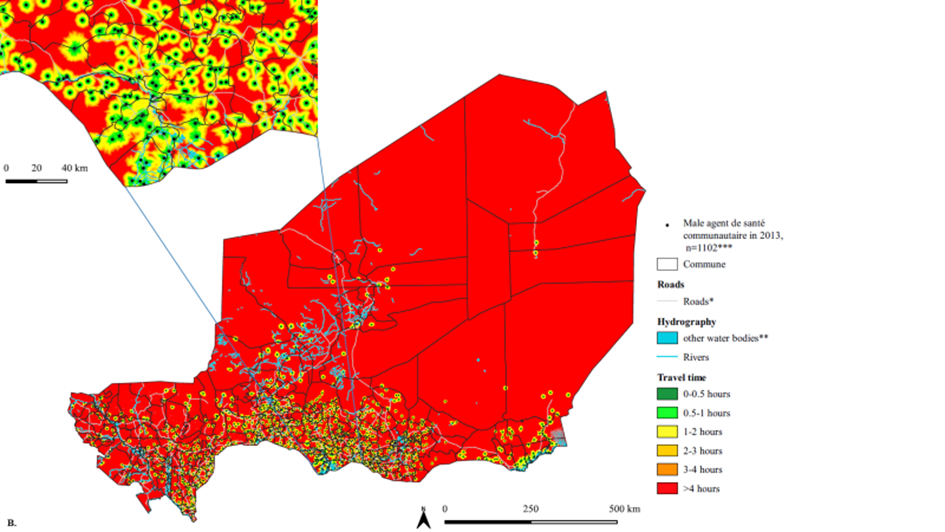

**Supplementary Figure 2 (B). Geographic accessibility (travel time in minutes, walking in dry conditions) to the nearest male ASC in 2013 at 100m x 100m resolution.**
Male ASC in 2013, n=1102. *For visualization purposes road classes limited to motorway, trunk, primary, secondary, and tertiary. **Other water bodies from landcover layer included permanent water bodies, temporary water bodies and herbaceous wetlands. ***Gender for 2 ASC was not recorded, and these ASC were excluded from the gender analysis.

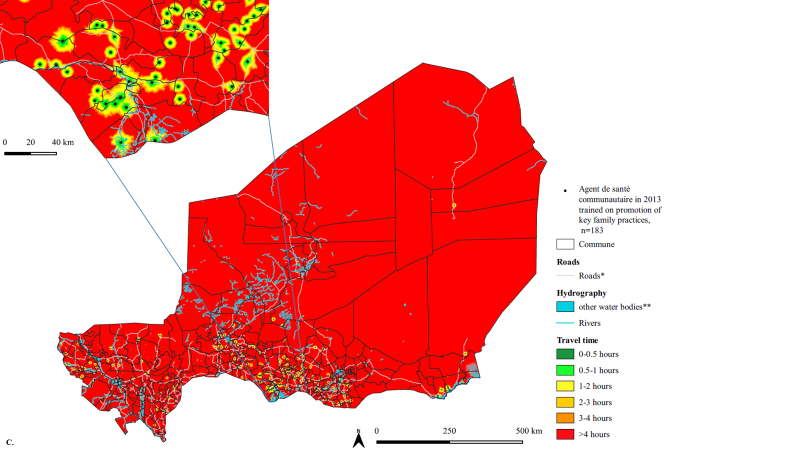

**Supplementary Figure 2 (C). Geographic accessibility (travel time in minutes, walking in dry conditions) to the nearest ASC in 2013 trained on promotion of key family practices at 100m x 100m resolution.**
ASC in 2013 trained on promotion of key family practices, n=183; *For visualization purposes road classes limited to motorway, trunk, primary, secondary, and tertiary; **other water bodies from landcover layer included permanent water bodies, temporary water bodies and herbaceous wetlands

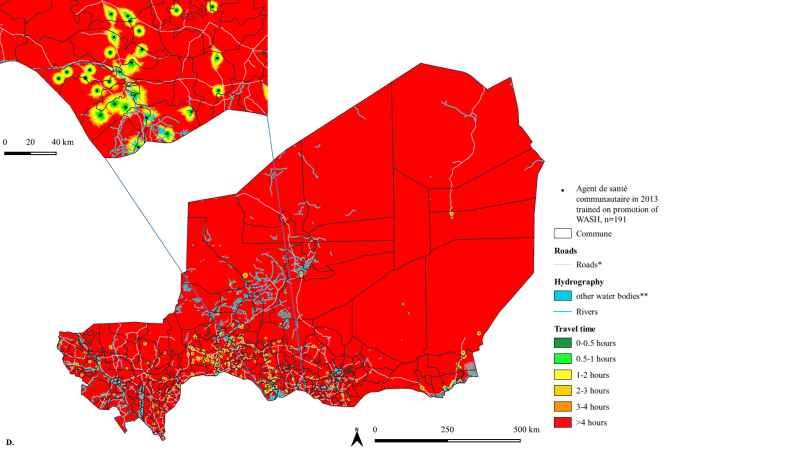

**Supplementary Figure 2 (D). Geographic accessibility (travel time in minutes, walking in dry conditions) to the nearest ASC in 2013 trained on promotion of WASH at 100m x 100m resolution.**
ASC in 2013 trained on promotion of WASH, n=1102. ASC=Agent de santé communautaire. WASH=water, sanitation, and hygiene. *For visualization purposes road classes limited to motorway, trunk, primary, secondary, and tertiary. **other water bodies from landcover layer included permanent water bodies, temporary water bodies and herbaceous wetlands

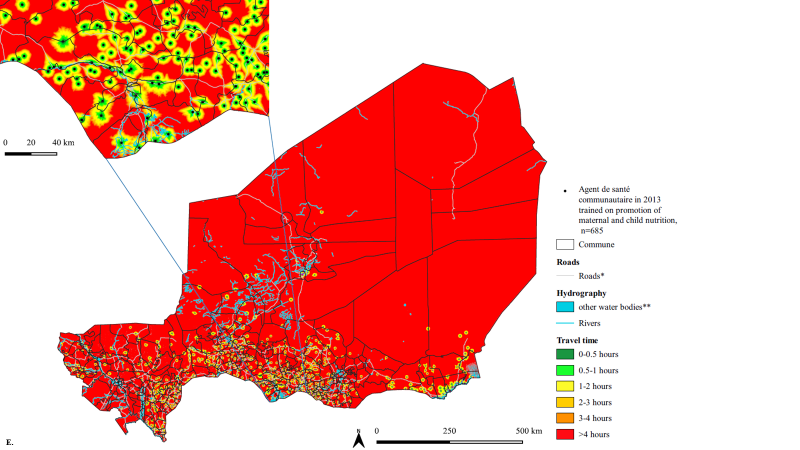

**Supplementary Figure 2 (E). Geographic accessibility (travel time in minutes, walking in dry conditions) to the nearest ASC in 2013 trained on promotion of maternal and child nutrition at 100m x 100m resolution.**
ASC in 2013 trained on promotion of maternal and child nutrition, n=685. ASC=Agent de santé communautaire. *For visualization purposes road classes limited to motorway, trunk, primary, secondary, and tertiary. **Other water bodies from landcover layer included permanent water bodies, temporary water bodies and herbaceous wetlands.

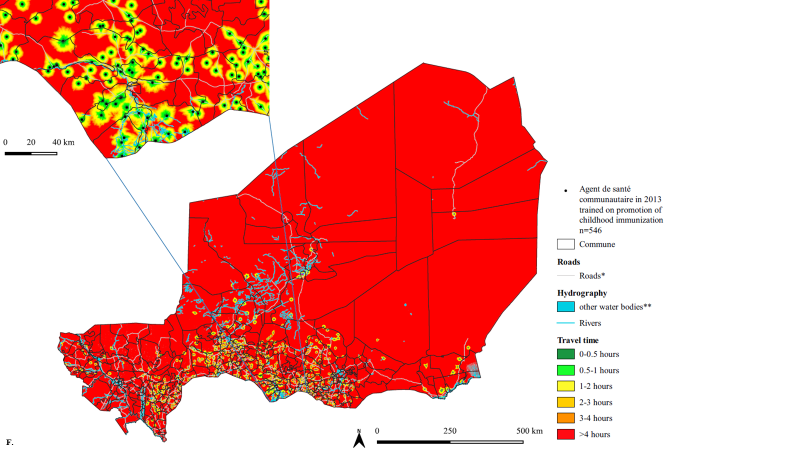

**Supplementary Figure 2 (F). Geographic accessibility (travel time in minutes, walking in dry conditions) to the nearest ASC in 2013 trained on promotion of childhood immunization at 100m x 100m resolution.**
ASC in 2013 trained on promotion of childhood immunization, n=546. ASC=Agent de santé communautaire. *For visualization purposes road classes limited to motorway, trunk, primary, secondary, and tertiary. **Other water bodies from landcover layer included permanent water bodies, temporary water bodies and herbaceous wetlands.

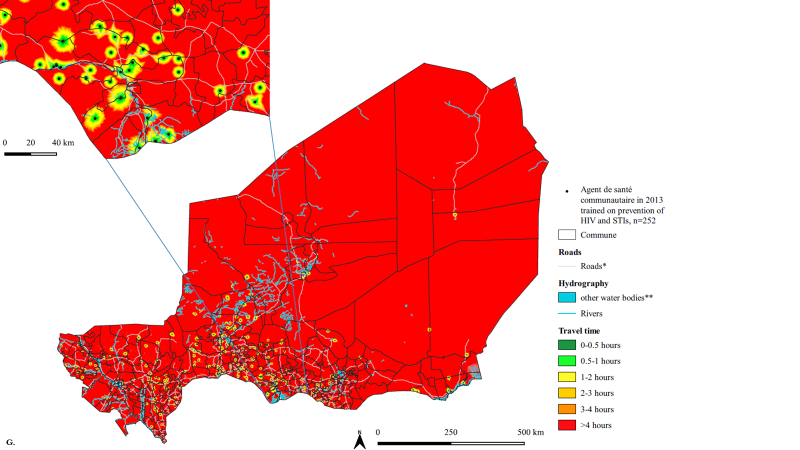

**Supplementary Figure 2 (G). Geographic accessibility (travel time in minutes, walking in dry conditions) to the nearest ASC in 2013 trained on prevention of HIV and STI at 100m x 100m resolution.**
ASC in 2013 trained on prevention of HIV and STI, n=252. ASC=Agent de santé communautaire. HIV=Human immunodeficiency virus. STI=sexually transmitted infection. *For visualization purposes road classes limited to motorway, trunk, primary, secondary, and tertiary. **Other water bodies from landcover layer included permanent water bodies, temporary water bodies and herbaceous wetlands.

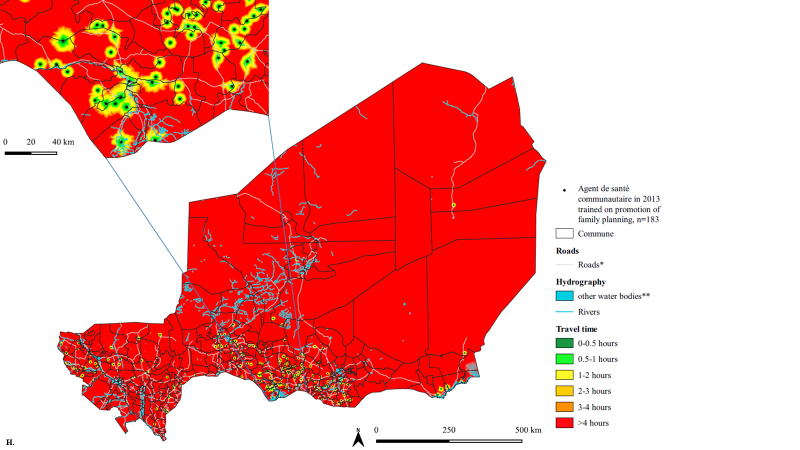

**Supplementary Figure 2 (H). Geographic accessibility (travel time in minutes, walking in dry conditions) to the nearest ASC in 2013 trained on family planning at 100m x 100m resolution.**
ASC in 2013 trained on family planning, n=183. ASC=Agent de santé communautaire. *For visualization purposes road classes limited to motorway, trunk, primary, secondary, and tertiary. **Other water bodies from landcover layer included permanent water bodies, temporary water bodies and herbaceous wetlands.

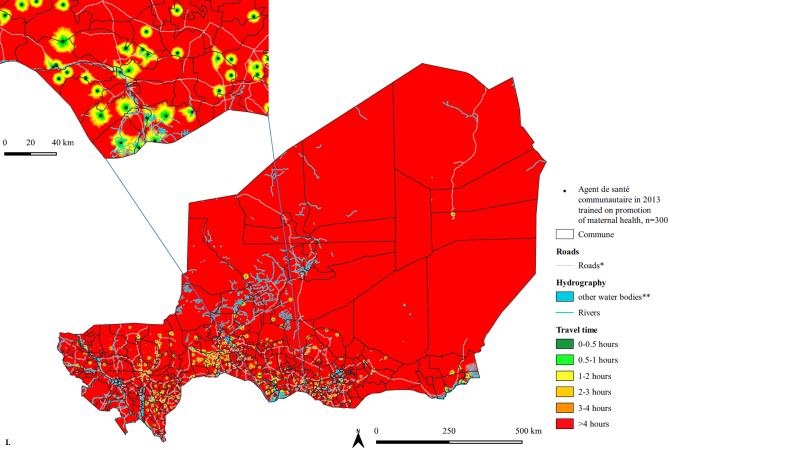

**Supplementary Figure 2 (I). Geographic accessibility (travel time in minutes, walking in dry conditions) to the nearest ASC in 2013 trained on promotion of maternal health at 100m x 100m resolution.**
ASC in 2013 trained on promotion of maternal health, n=300. ASC=Agent de santé communautaire. *For visualization purposes road classes limited to motorway, trunk, primary, secondary, and tertiary. **Other water bodies from landcover layer included permanent water bodies, temporary water bodies and herbaceous wetlands.

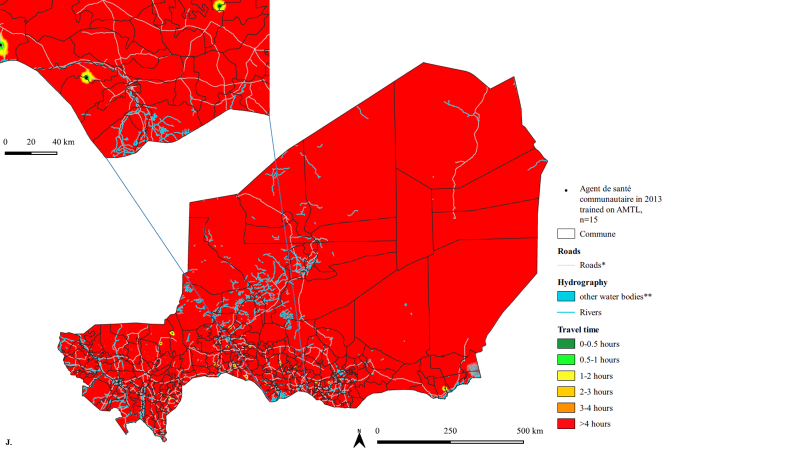

**Supplementary Figure 2 (J). Geographic accessibility (travel time in minutes, walking in dry conditions) to the nearest ASC in 2013 trained on AMTL at 100m x 100m resolution.**
ASC in 2013 trained on AMTL, n=15. ASC=Agent de santé communautaire. AMTL=Active management of the third stage of labor. *For visualization purposes road classes limited to motorway, trunk, primary, secondary, and tertiary. **Other water bodies from landcover layer included permanent water bodies, temporary water bodies and herbaceous wetlands.

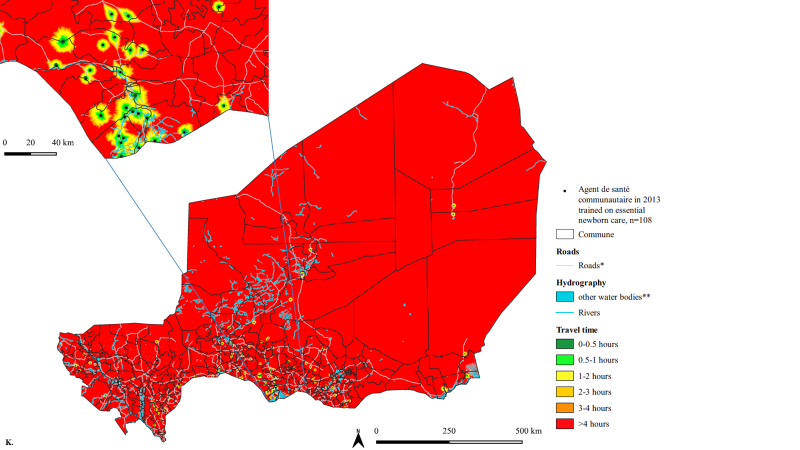

**Supplementary Figure 2 (K). Geographic accessibility (travel time in minutes, walking in dry conditions) to the nearest ASC in 2013 trained on essential newborn care at 100m x 100m resolution.**
ASC in 2013 trained on essential newborn care, n=108. ASC=Agent de santé communautaire. *For visualization purposes road classes limited to motorway, trunk, primary, secondary, and tertiary. **Other water bodies from landcover layer included permanent water bodies, temporary water bodies and herbaceous wetlands.

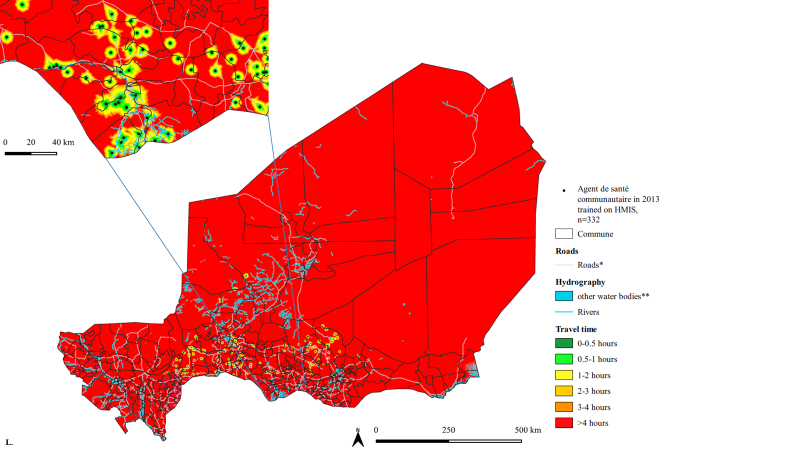

**Supplementary Figure 2 (L). Geographic accessibility (travel time in minutes, walking in dry conditions) to the nearest ASC in 2013 trained on the HMIS at 100m x 100m resolution.**
ASC in 2013 trained on the HMIS, n=332. ASC=Agent de santé communautaire. HMIS=Health management information system. *For visualization purposes road classes limited to motorway, trunk, primary, secondary, and tertiary. **Other water bodies from landcover layer included permanent water bodies, temporary water bodies and herbaceous wetlands.

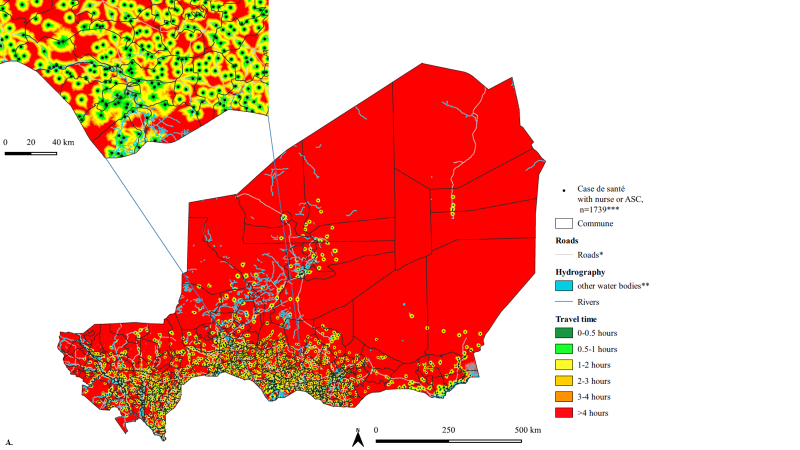

**Supplementary Figure 3 (A). Geographic accessibility (travel time in minutes, walking in dry conditions) to the nearest CS in 2013 with a nurse or ASC at 100m x 100m resolution.**
CS in 2013 with a nurse or ASC, n=1739 (***does not include 13 CS that met this criteria but did not have geocoordinates). CS=Case de santé. ASC=Agent de santé communautaire. *For visualization purposes road classes limited to motorway, trunk, primary, secondary, and tertiary. **Other water bodies from landcover layer included permanent water bodies, temporary water bodies and herbaceous wetlands.

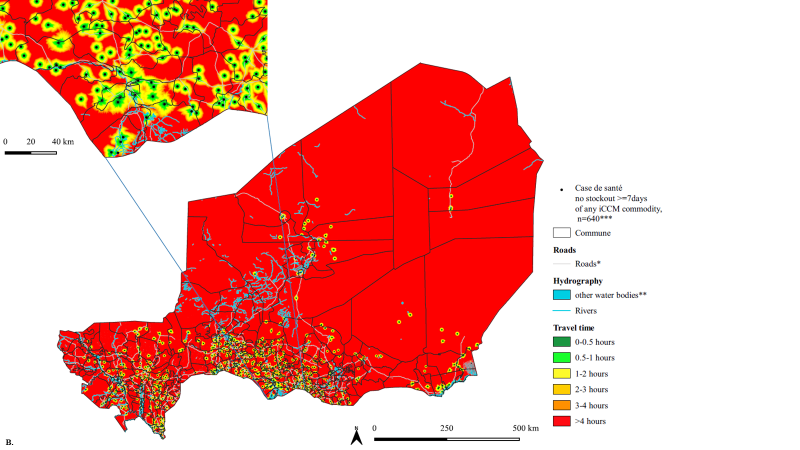

**Supplementary Figure 3 (B). Geographic accessibility (travel time in minutes, walking in dry conditions) to the nearest CS in 2013 with no severe stockout of any iCCM commodities at 100m x 100m resolution.**
CS with no stockout of any iCCM commodity lasting longer than seven days, n=640 (***does not include 8 CS that met this criteria but did not have geocoordinates). iCCM commodities = RDT and AL for malaria, low osmolarity ORS and zinc sulfate for diarrhea, cotrimoxazole (pill or syrup) for pneumonia. A stockout of any of these commodities lasting longer than 7 days resulted in the CS being considered as a CS with a severe stockout of any iCCM commodity. CS=Case de santé. ASC=Agent de santé communautaire. iCCM=integrated community case management. RDT=rapid diagnostic test for malaria. AL=artemether-lumefantrine. *For visualization purposes road classes limited to motorway, trunk, primary, secondary, and tertiary. **Other water bodies from landcover layer included permanent water bodies, temporary water bodies and herbaceous wetlands.

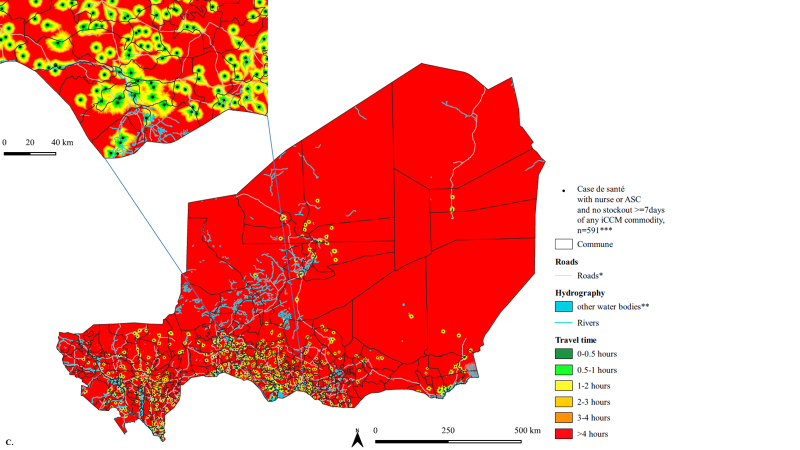

**Supplementary Figure 3 (C). Geographic accessibility (travel time in minutes, walking in dry conditions) to the nearest CS in 2013 with a nurse or ASC and no severe stockout of any iCCM commodities at 100m x 100m resolution.**
CS in 2013 with a nurse or ASC and no stockout of any iCCM commodity lasting longer than seven days, n=591 (does not include 7 CS that met this criteria but did not have geocoordinates). iCCM commodities = RDT and AL for malaria, low osmolarity ORS and zinc sulfate for diarrhea, cotrimoxazole (pill or syrup) for pneumonia. A stockout of any of these commodities lasting longer than 7 days resulted in the CS being considered as a CS with a severe stockout of any iCCM commodity. CS=Case de santé. ASC=Agent de santé communautaire. iCCM=integrated community case management. RDT=rapid diagnostic test for malaria. AL=artemether-lumefantrine. *For visualization purposes road classes limited to motorway, trunk, primary, secondary, and tertiary. **Other water bodies from landcover layer included permanent water bodies, temporary water bodies and herbaceous wetlands.

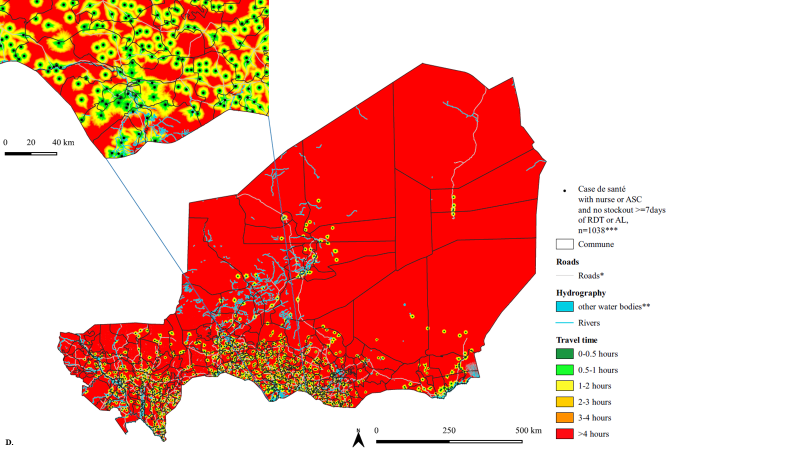

**Supplementary Figure 3 (D). Geographic accessibility (travel time in minutes, walking in dry conditions) to the nearest CS in 2013 with a nurse or ASC and no severe stockout of RDT or AL at 100m x 100m resolution.**
CS in 2013 with a nurse or ASC and no stockout of RDT or AL lasting longer than seven days, n=1038 (does not include 11 CS that met this criteria but did not have geocoordinates). A stockout of >= 7 days was considered a severe stockout. CS=Case de santé. ASC=Agent de santé communautaire. RDT=rapid diagnostic test for malaria. AL=artemether-lumefantrine. *For visualization purposes road classes limited to motorway, trunk, primary, secondary, and tertiary. **Other water bodies from landcover layer included permanent water bodies, temporary water bodies and herbaceous wetlands.

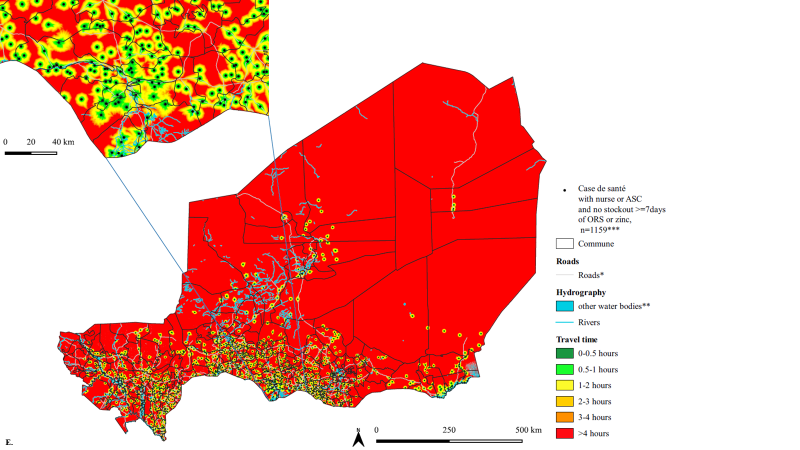

**Supplementary Figure 3 (E). Geographic accessibility (travel time in minutes, walking in dry conditions) to the nearest CS in 2013 with a nurse or ASC and no severe stockout of ORS or zinc at 100m x 100m resolution.**
CS in 2013 with a nurse or ASC and no stockout of ORS or zinc lasting longer than seven days, n=1159 (does not include 9 CS that met this criteria but did not have geocoordinates). A stockout of >= 7 days was considered a severe stockout. CS=Case de santé. ASC=Agent de santé communautaire. ORS = low osmolarity oral rehydration solution. *For visualization purposes road classes limited to motorway, trunk, primary, secondary, and tertiary. **Other water bodies from landcover layer included permanent water bodies, temporary water bodies and herbaceous wetlands.

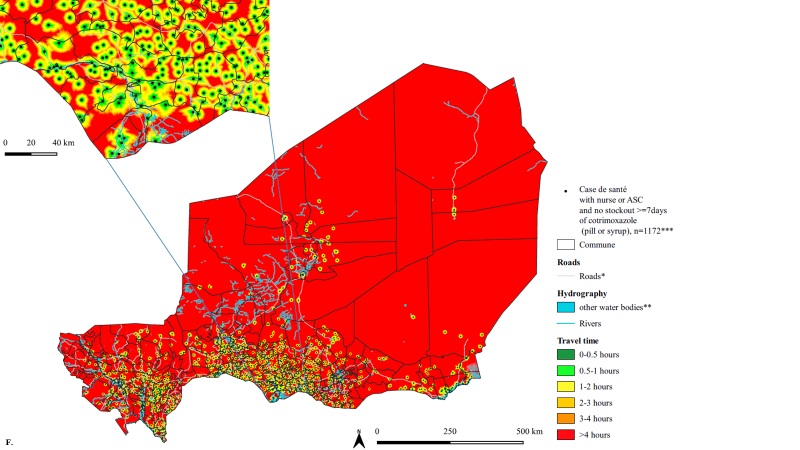

**Supplementary Figure 3 (F). Geographic accessibility (travel time in minutes, walking in dry conditions) to the nearest CS in 2013 with a nurse or ASC and no severe stockout of cotrimoxazole at 100m x 100m resolution.**
CS in 2013 with a nurse or ASC and no stockout of cotrimoxazole (pill or syrup) lasting longer than seven days, n=1172 (does not include 7 CS that met this criteria but did not have geocoordinates). A stockout of >= 7 days was considered a severe stockout. CS=Case de santé. ASC=Agent de santé communautaire. *For visualization purposes road classes limited to motorway, trunk, primary, secondary, and tertiary. **Other water bodies from landcover layer included permanent water bodies, temporary water bodies and herbaceous wetlands.

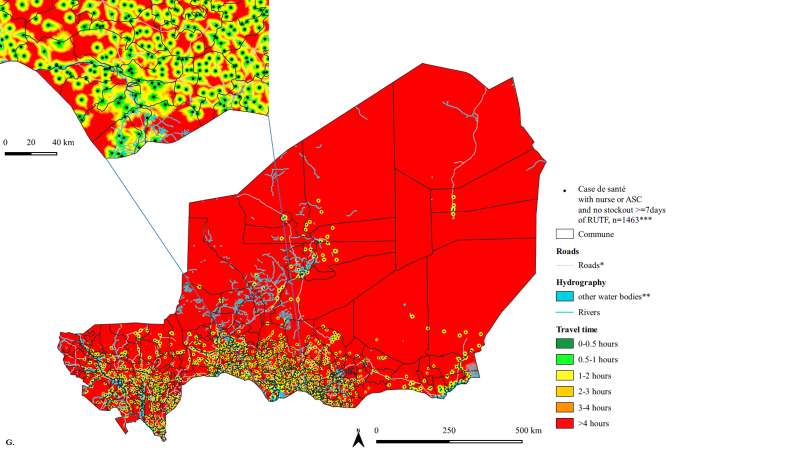

**Supplementary Figure 3 (G). Geographic accessibility (travel time in minutes, walking in dry conditions) to the nearest CS in 2013 with a nurse or ASC and no severe stockout of RUTF at 100m x 100m resolution.**
CS in 2013 with a nurse or ASC and no stockout of RUTF lasting longer than seven days, n=1463 (does not include 9 CS that met this criteria but did not have geocoordinates). A stockout of >= 7 days was considered a severe stockout. CS=Case de santé. ASC=Agent de santé communautaire. RUTF=ready-to-eat therapeutic food. *For visualization purposes road classes limited to motorway, trunk, primary, secondary, and tertiary. **Other water bodies from landcover layer included permanent water bodies, temporary water bodies and herbaceous wetlands.

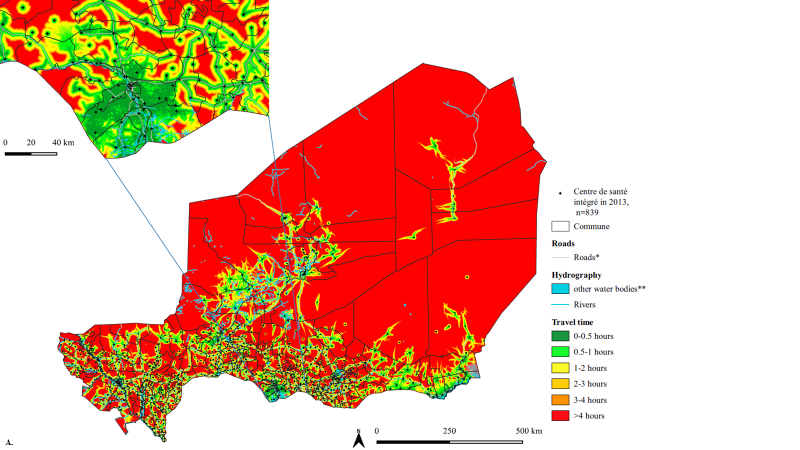

**Supplementary Figure 4. (A) Geographic accessibility (travel time in minutes, walking + motorized transportation in dry conditions) to the nearest CSI in 2013 at 100m x 100m resolution.**
CSI in 2013, n=839. CSI=Centre de santé intégrée. *For visualization purposes road classes limited to motorway, trunk, primary, secondary, and tertiary. **Other water bodies from landcover layer included permanent water bodies, temporary water bodies and herbaceous wetlands.

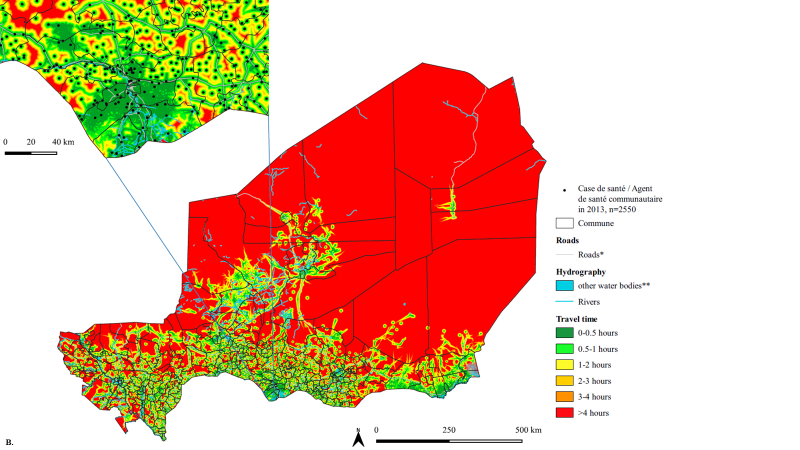

**Supplementary Figure 4. (B) Geographic accessibility (travel time in minutes, walking + motorized transportation in dry conditions) to the nearest CS/ASC in 2013 at 100m x 100m resolution.**
CS/ASC in 2013, n=2550. CS/ASC=Case de santé / Agent de santé communautaire. ASC=Agent de santé communautaire. *For visualization purposes road classes limited to motorway, trunk, primary, secondary, and tertiary. **Other water bodies from landcover layer included permanent water bodies, temporary water bodies and herbaceous wetlands.

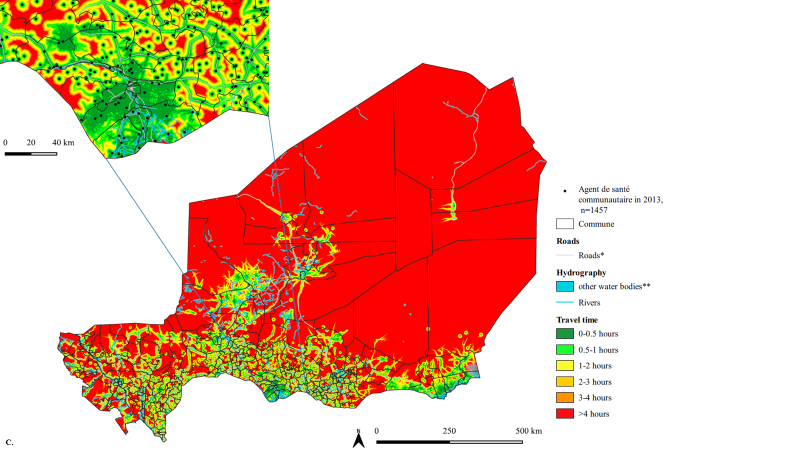

**Supplementary Figure 4. (C) Geographic accessibility (travel time in minutes, walking + motorized transportation in dry conditions) to the nearest ASC in 2013 at 100m x 100m resolution.**
ASC in 2013, n=1457. ASC=Agent de santé communautaire. *For visualization purposes road classes limited to motorway, trunk, primary, secondary, and tertiary. **Other water bodies from landcover layer included permanent water bodies, temporary water bodies and herbaceous wetlands.

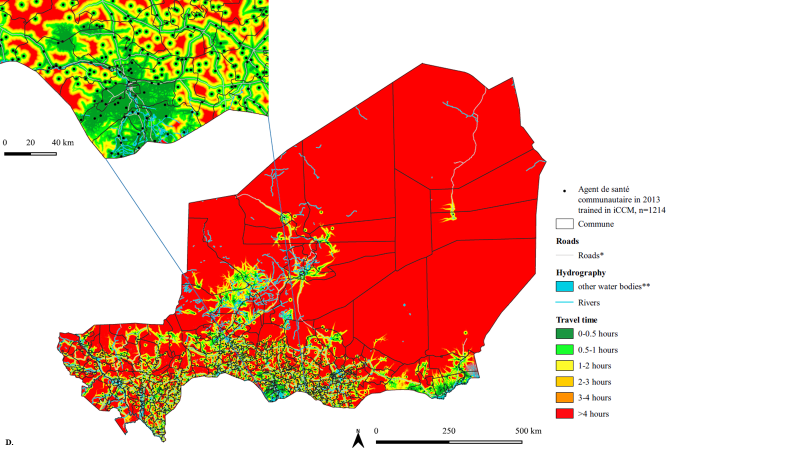

**Supplementary Figure 4. (D) Geographic accessibility (travel time in minutes, walking + motorized transportation in dry conditions) to the nearest ASC in 2013 trained on iCCM at 100m x 100m resolution.**
ASC in 2013 trained on iCCM, n=1214. ASC=Agent de santé communautaire. iCCM=integrated community case management. *For visualization purposes road classes limited to motorway, trunk, primary, secondary, and tertiary. **Other water bodies from landcover layer included permanent water bodies, temporary water bodies and herbaceous wetlands.

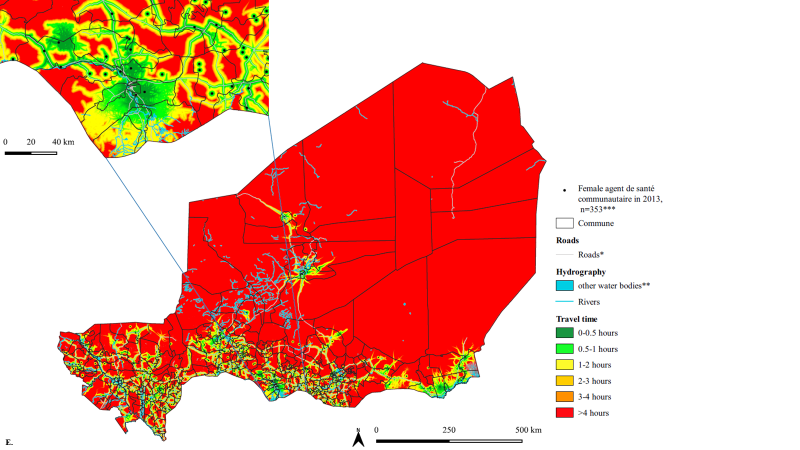

**Supplementary Figure 4. (E) Geographic accessibility (travel time in minutes, walking + motorized transportation in dry conditions) to the nearest female ASC in 2013 at 100m x 100m resolution.**
Female ASC in 2013, n=353. ASC=Agent de santé communautaire. *For visualization purposes road classes limited to motorway, trunk, primary, secondary, and tertiary. **Other water bodies from landcover layer included permanent water bodies, temporary water bodies and herbaceous wetlands. ***Gender for 2 ASC was not recorded, and these ASC were excluded from the gender analysis.

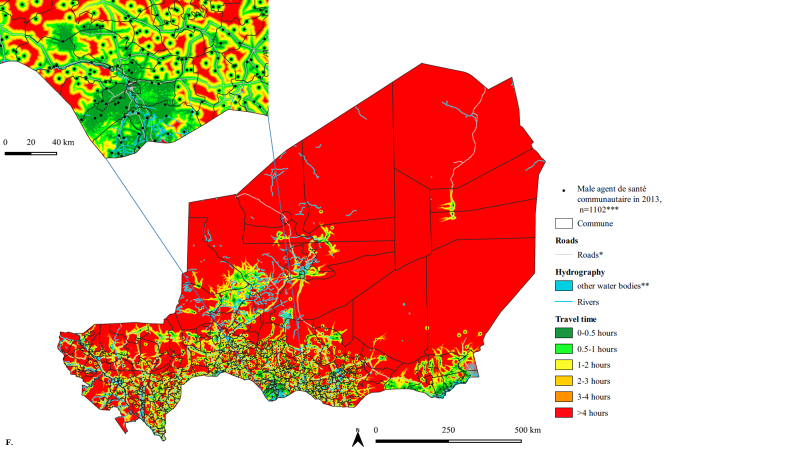

**Supplementary Figure 4. (F) Geographic accessibility (travel time in minutes, walking + motorized transportation in dry conditions) to the nearest male ASC in 2013 at 100m x 100m resolution.**
Male ASC in 2013 trained on iCCM, n=1102. ASC=Agent de santé communautaire. *For visualization purposes road classes limited to motorway, trunk, primary, secondary, and tertiary. **Other water bodies from landcover layer included permanent water bodies, temporary water bodies and herbaceous wetlands. ***Gender for 2 ASC was not recorded, and these ASC were excluded from the gender analysis.

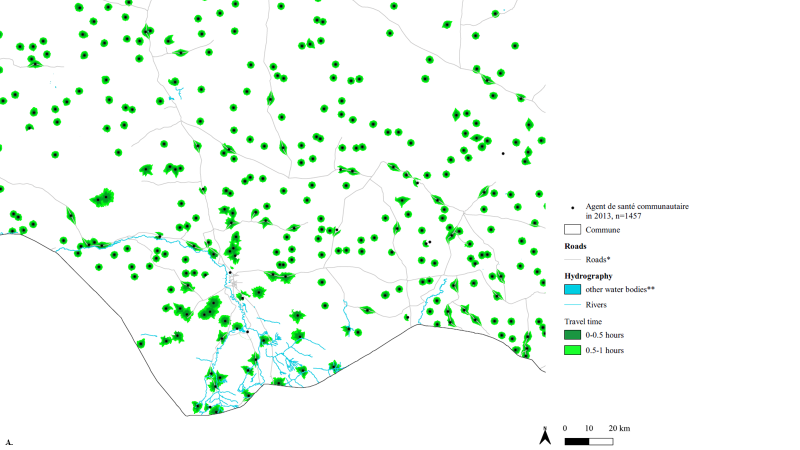

**Supplementary Figure 5 (A). Contribution of ASC to additional geographic accessibility beyond the existing CSI and CS (without ASC) networks in 2013 (walking scenario) at 100m x 100m resolution.**
ASC in 2013, n=1457, walking scenario. ASC=Agent de santé communautaire. *For visualization purposes road classes limited to motorway, trunk, primary, secondary, and tertiary. **Other water bodies from landcover layer included permanent water bodies, temporary water bodies and herbaceous wetlands.

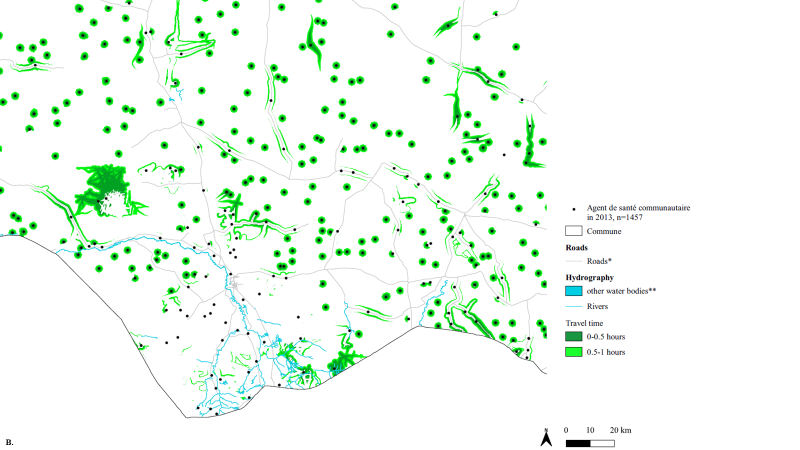

**Supplementary Figure 5 (B). Contribution of ASC to additional geographic accessibility beyond the existing CSI and CS (without ASC) networks in 2013 (walking + motorized transportation scenario) at 100m x 100m resolution.**
ASC in 2013, n=1457, walking + motorized transportation scenario. ASC=Agent de santé communautaire. *For visualization purposes road classes limited to motorway, trunk, primary, secondary, and tertiary. **Other water bodies from landcover layer included permanent water bodies, temporary water bodies and herbaceous wetlands.

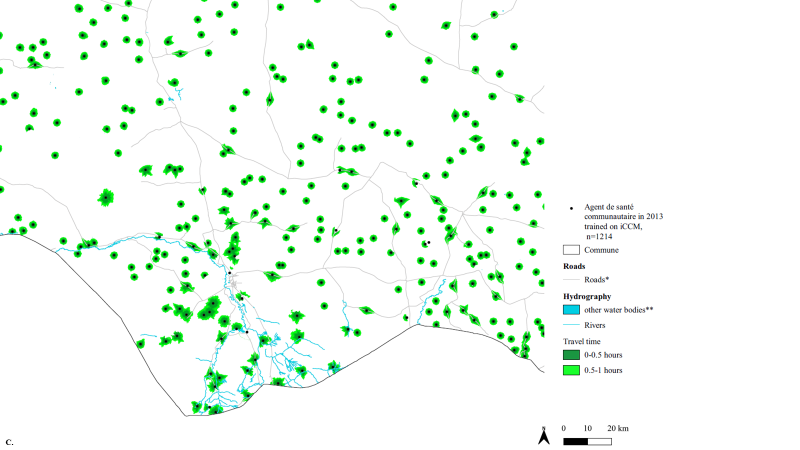

**Supplementary Figure 5 (C). Contribution of ASC trained on iCCM to additional geographic accessibility beyond the existing CSI and CS (without ASC) networks in 2013 (walking scenario) at 100m x 100m resolution.**
ASC in 2013 trained on iCCM, n=1214, walking scenario. ASC=Agent de santé communautaire. *For visualization purposes road classes limited to motorway, trunk, primary, secondary, and tertiary. **Other water bodies from landcover layer included permanent water bodies, temporary water bodies and herbaceous wetlands.

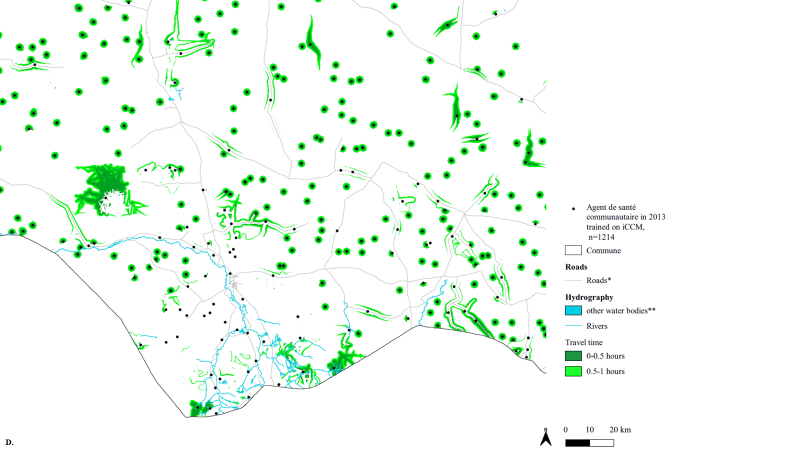

**Supplementary Figure 5 (D). Contribution of ASC trained on iCCM to additional geographic accessibility beyond the existing CSI and CS (without ASC) networks in 2013 (walking + motorized transportation scenario) at 100m x 100m resolution.**
ASC in 2013 trained on iCCM, n=1214, walking + motorized transportation scenario. ASC=Agent de santé communautaire. *For visualization purposes road classes limited to motorway, trunk, primary, secondary, and tertiary. **Other water bodies from landcover layer included permanent water bodies, temporary water bodies and herbaceous wetlands.

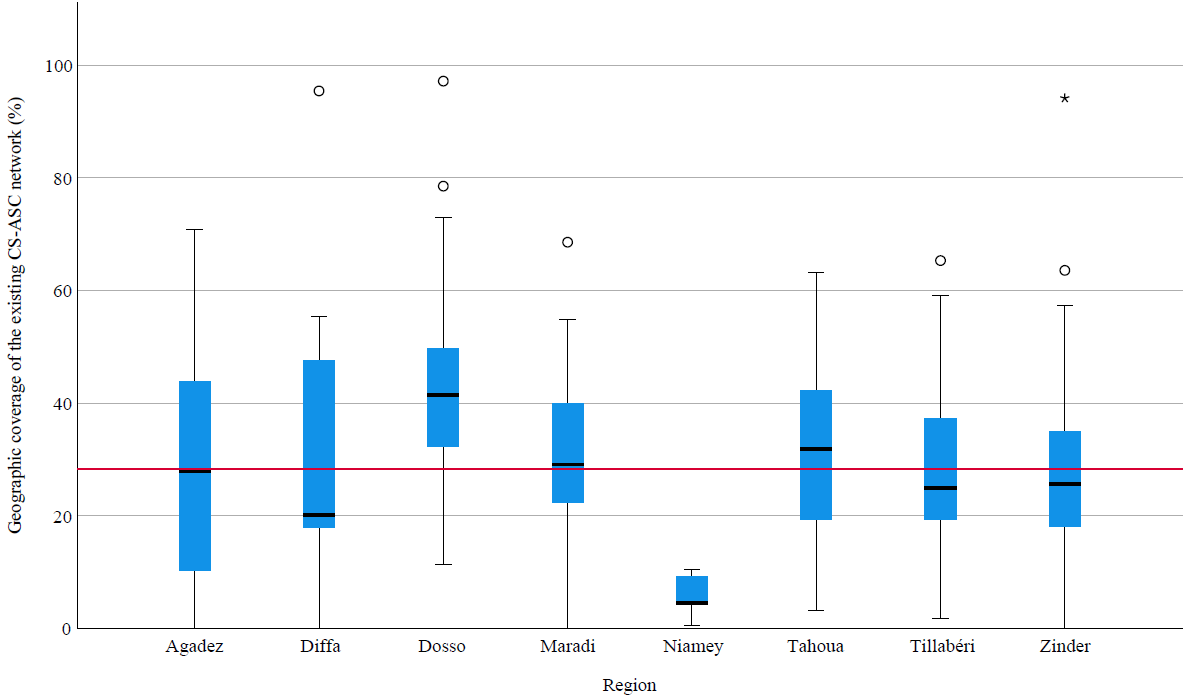

**Supplementary Figure 6. Median and interquartile range of geographic coverage at commune level (administrative level 3) of the residual population beyond the geographic coverage of the CSI network that were covered by the existing CS-ASC network, by region (administrative level 1) at 1km x 1km resolution.**
Median and interquartile range of geographic coverage at commune level (administrative level 3) of the residual population beyond the geographic coverage (60-minute catchment, walking scenario) of the CSI network that were covered by the existing CS-ASC network (60-minute catchment, walking scenario) by region (administrative level 1). Red line at national geographic coverage of 28.3%.

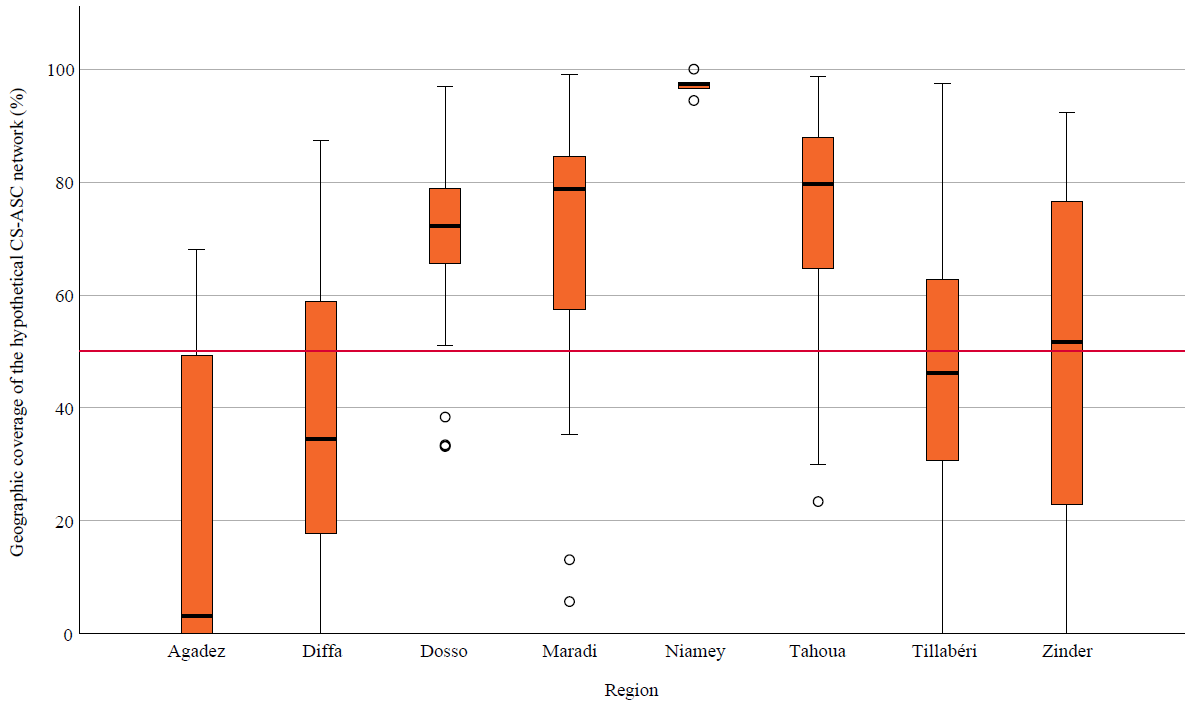

**Supplementary Figure 7. Median and interquartile range of geographic coverage at commune level (administrative level 3) of the residual population beyond the geographic coverage of the CSI network that were covered by the hypothetical CS-ASC network deployed to optimize geographic coverage of the residual population, by region (administrative level 1) at 1km x 1km resolution.**
Median and interquartile range of geographic coverage at commune level (administrative level 3) of the residual population beyond the geographic coverage (60-minute catchment, walking scenario) of the CSI network that were covered by the hypothetical CS-ASC network deployed to optimize geographic coverage of the residual population (60-minute catchment, walking scenario) by region (administrative level 1). Red line at national geographic coverage of 52.9%.

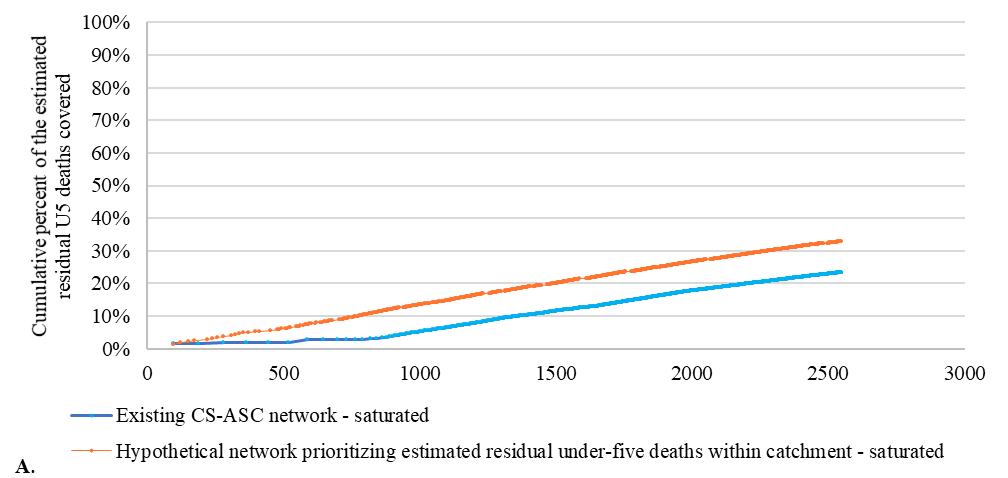

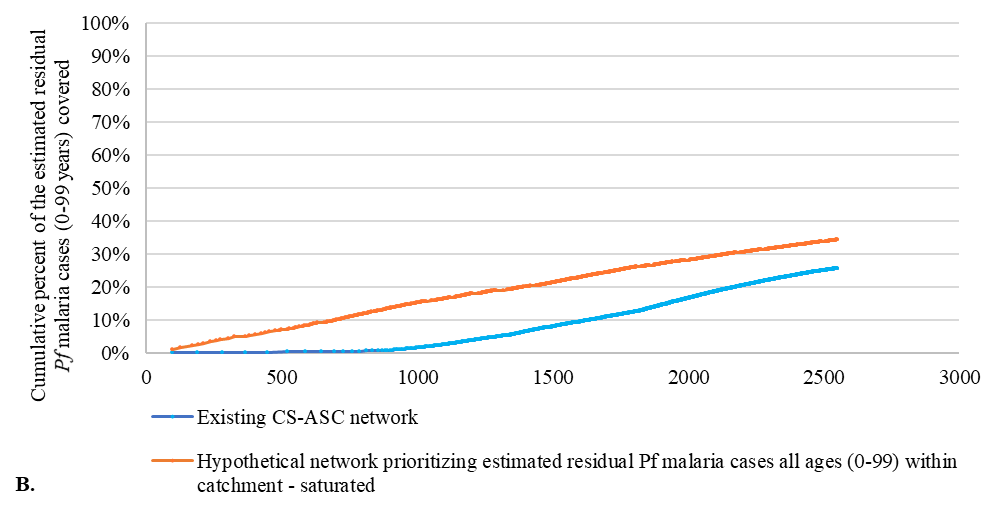

**Supplementary Figure 8. Targeting with saturation at 1km x 1km resolution.**
Targeting of the existing CS-ASC network compared to hypothetical networks deployed at “saturation” to optimize A) the estimated residual under-five deaths in 2013 and B) the estimated residual Pf malaria cases in 2013 beyond the geographic coverage of the existing CSI network (60-minute catchment, walking scenario). “Saturation” means sufficient CS-ASC were deployed to cover 100% of the estimated residual population within the catchment of each CS-ASC. CSI=Centre de santé intégré. CS=Case de santé. ASC=Agent de santé communautaire. Pf=Plasmodium falciparum.

Assumptions for (A): The hypothetical CS-ASC network started with 5796 candidate CS-ASC sites. Candidate sites were sorted from highest to lowest based on the estimated residual under-five deaths within a 60-minute catchment using the variable "amPopCatchmentTotal". The number of CS-ASC in the existing network was adjusted to ensure full coverage of the residual population in the theoretical catchment area of the existing CS-ASC network, up to 2550 CS-ASC. First, the estimated residual population in 2013 beyond the geographic coverage of the CSI network (60 minutes walking) was extracted for each CS-ASC catchment within the existing CS-ASC network. Next, the national normative ratio of 1 CS-ASC per 2500 population was applied to the estimated residual population within the theoretical 60-minute catchment of each CS-ASC site within the existing CS-ASC network to calculate the number of CS-ASC needed to cover 100% of the population within the existing CS-ASC network. The first 2550 CS-ASC in the existing CS-ASC network were retained for the comparison. This was repeated for the hypothetical CS-ASC network to calculate the number of CS-ASC needed to cover 100% of the population within the hypothetical CS-ASC network and identify the first 2550 CS-ASC in the hypothetical CS-ASC network.

Assumptions for (B): The hypothetical CS-ASC network started with 5796 candidate CS-ASC sites. Candidate sites were sorted from highest to lowest based on the estimated residual Pf malaria cases within a 60-minute catchment using the variable "amPopCatchmentTotal". The number of CS-ASC in the existing network was adjusted to ensure full coverage of the residual population in the theoretical catchment area of the existing CS-ASC network, up to 2550 CS-ASC. First, the estimated residual population in 2013 beyond the geographic coverage of the CSI network (60-minute walking) was extracted for each CS-ASC catchment within the existing CS-ASC network. Next, the national normative ratio of 1 CS-ASC per 2500 population was applied to the estimated residual population within the theoretical 60-minute catchment of each CS-ASC site within the existing CS-ASC network to calculate the number of CS-ASC needed to cover 100% of the population within the existing CS-ASC network. The first 2550 CS-ASC in the existing CS-ASC network were retained for the comparison. This was repeated for the hypothetical CS-ASC network to calculate the number of CS-ASC needed to cover 100% of the population within the hypothetical CS-ASC network and identify the first 2550 CS-ASC in the hypothetical CS-ASC network.

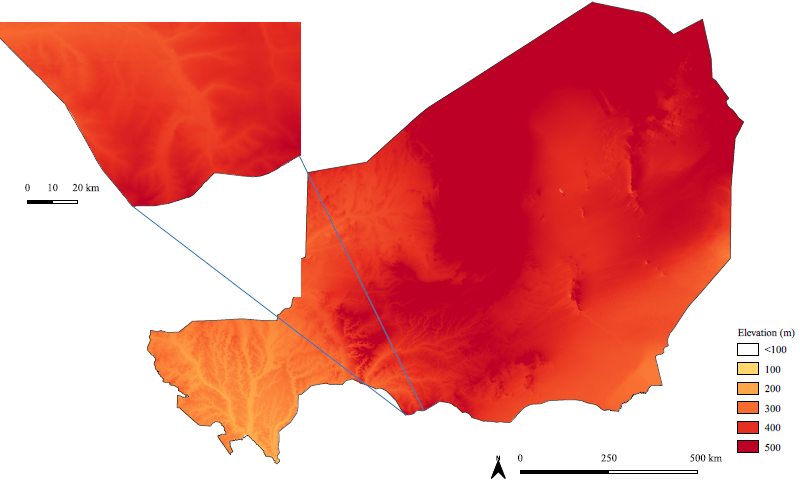

**Supplementary Figure 9. Digital elevation model at 100m x 100m resolution.**
NASA SRTMGL1 version 003 (approximately 30m x30m), resampled to 100m x 100m and 1km x 1km (later not shown). Accessed 4 October 2018. Inset near Madarounfa commune in the Maradi region.^1^

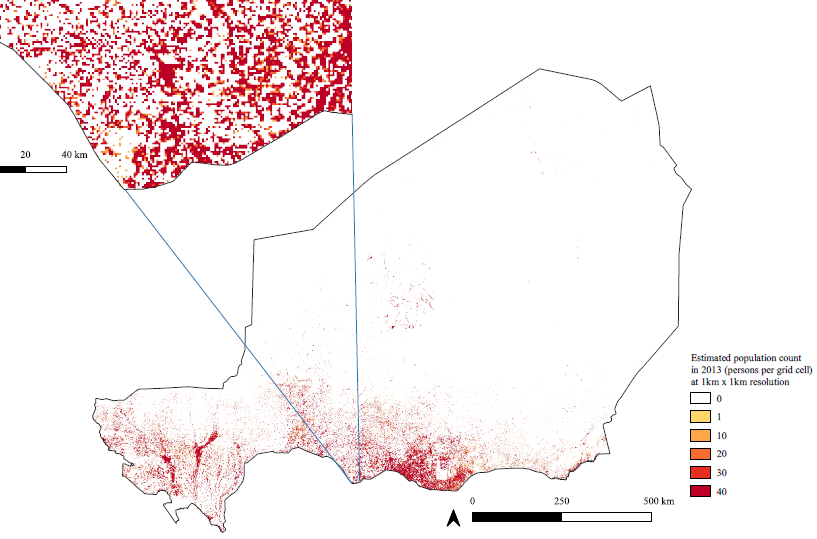

**Supplementary Figure 10. Estimated population count in 2013 (persons per grid cell) at 1km x 1km resolution.**
Population layers produced at 100m x 100m resolution and 1km x 1km resolution. 1km x 1km shown here for ease of visualization. HRSL 2015 at approximately 30m x 30m resampled to 100m x 100m and 1km x 1km resolutions and adjusted to Worldpop population totals for 2013 at administrative level 3 (commune). Inset near Madarounfa commune in the Maradi region. Source: Derived from Facebook Connectivity Lab and Center for International Earth Science Information Network - CIESIN - Columbia University. 2016. High Resolution Settlement Layer (HRSL). Source imagery for HRSL © 2016 [DigitalGlobe](http://explore.digitalglobe.com/Basemap-Vivid.html). Accessed 4 October 2018.^2^ WorldPop (www.worldpop.org - School of Geography and Environmental Science, University of Southampton; Department of Geography and Geosciences, University of Louisville; Département de Géographie, Université de Namur) and Center for International Earth Science Information Network (CIESIN), Columbia University (2018). Global High Resolution Population Denominators Project - Funded by The Bill and Melinda Gates Foundation (OPP1134076). <https://dx.doi.org/10.5258/SOTON/WP00645>. Accessed 4 October 2018.^3^

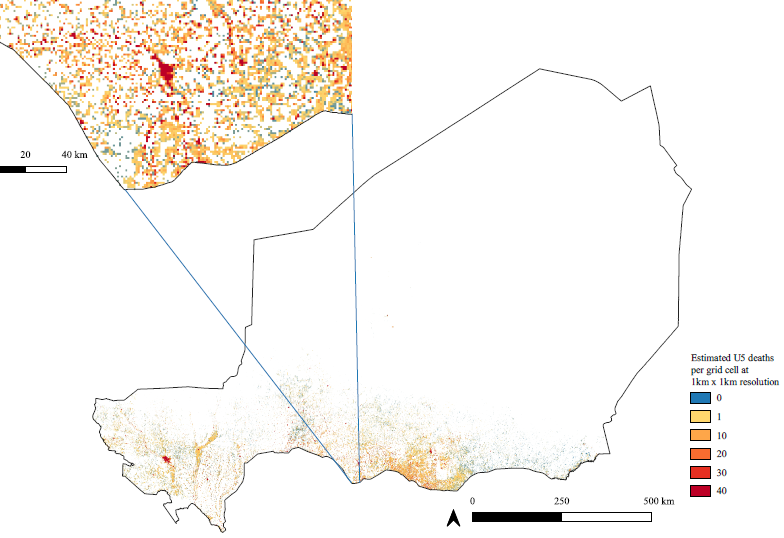

**Supplementary Figure 11. Mean U5 deaths in 2013 at 1km x 1km.**
Derived from the mean U5 mortality rate in 2013 (IHME) at 5km x 5km, resampled to 1kmx1km and multiplied by the 2013 under-five population layer (derived from HRSL and Worldpop population layers, described in Methods). U5=under-five. Source: Institute for Health Metrics and Evaluation (IHME). Low- and Middle-Income Country Neonatal, Infant, and Under-5 Mortality Geospatial Estimates 2000-2017. Seattle, United States: Institute for Health Metrics and Evaluation (IHME), 2019.^4^ Described in Burstein R, Henry JH, Collison ML, Marczak LB, Sligar A, Watson S, et al. Mapping 123 million neonatal, infant, and child deaths between 2000 and 2017. Nature. 16 October 2019.^5^

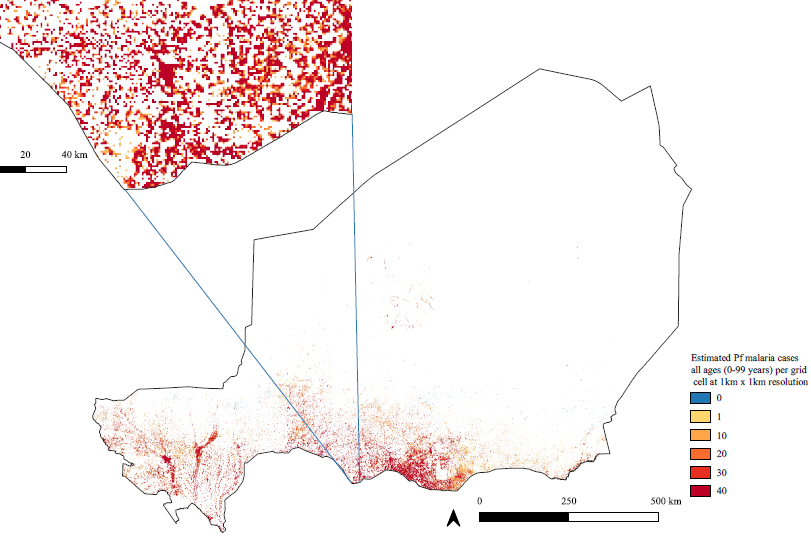

**Supplementary Figure 12. Estimated *Pf* malaria cases among all ages (0-99 years) per grid cell at 1km x 1km resolution.**
Annual mean incidence of Plasmodium falciparum (Pf) malaria among all ages (0-99 years) in 2013 globally at 2.5 arcminutes (approximately 5km x 5km) resolution from Weiss et al 2019, reprojected to 1km x 1km resolution and multiplied by the estimated population in 2013 (see Methods). Source: Weiss DJ, Lucas TCD, Nguyen M, et al. Mapping the global prevalence, incidence, and mortality of Plasmodium falciparum, 2000–17: a spatial and temporal modelling study. Lancet 2019; published online June 19. <http://dx.doi.org/10.1016/S0140-6736(19)31097-9>.^6^

 
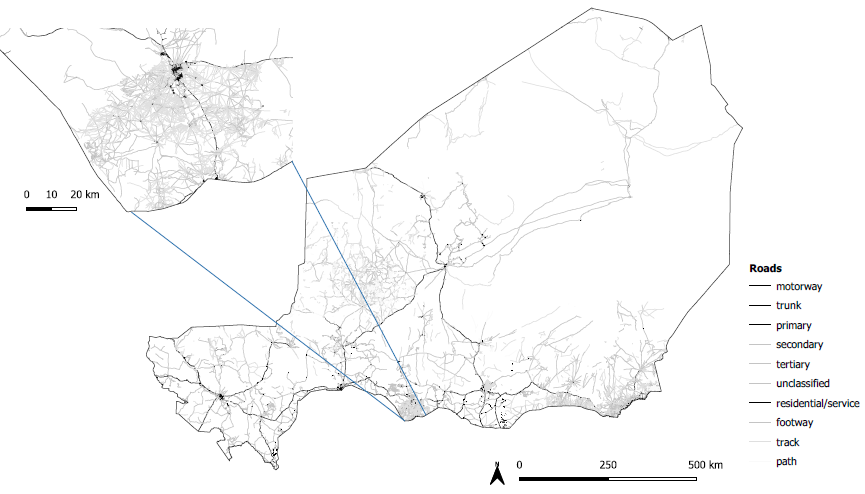

**Supplementary Figure 13. Road network**
Source: Humanitarian OpenStreetMap Team (HOT). Accessed 1 August 2018.^7^

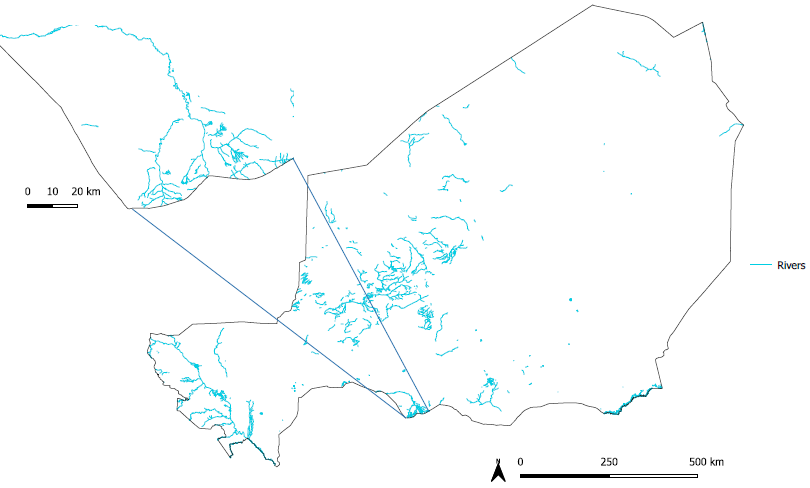

**Supplementary Figure 14. Rivers**
Source: Humanitarian OpenStreetMap Team (HOT). Accessed 15 January 2018. Note: other water bodies included in the land cover layer.^8^

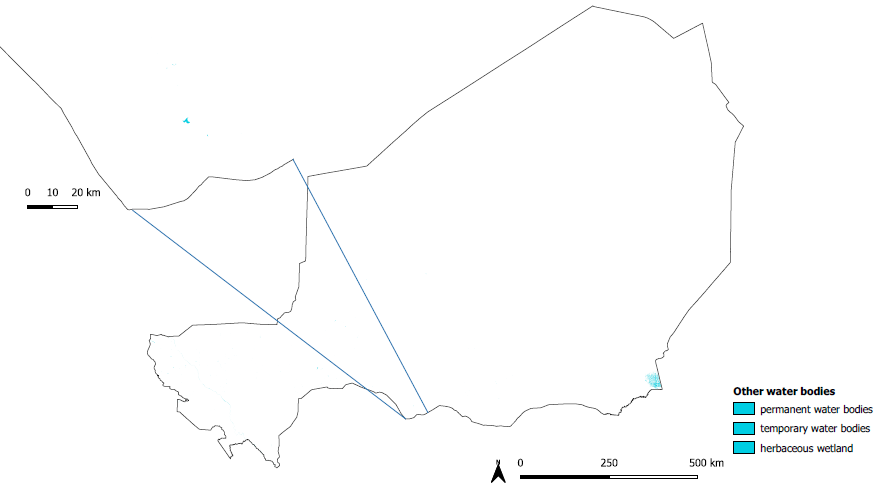

**Supplementary Figure 15. Other water bodies**
Includes the following land classes from the source below: permanent water bodies, temporary water bodies and herbaceous wetland. Source: Buchhorn, M., Smets, B., Bertels, L., Lesiv, M., Tsendbazar, N.-E., Masiliunas, D., Herold, M., Fritz, S. (2019). Copernicus Global Land Service: Land Cover 100m, Collection 2, epoch 2018 Africa Demo (Version V2.1.1). Zenodo. DOI:10.5281/zenodo.3518087. Copernicus Global Land Service, accessed on 27 March 2018 at <https://land.copernicus.eu/global/products/lc>.^9^

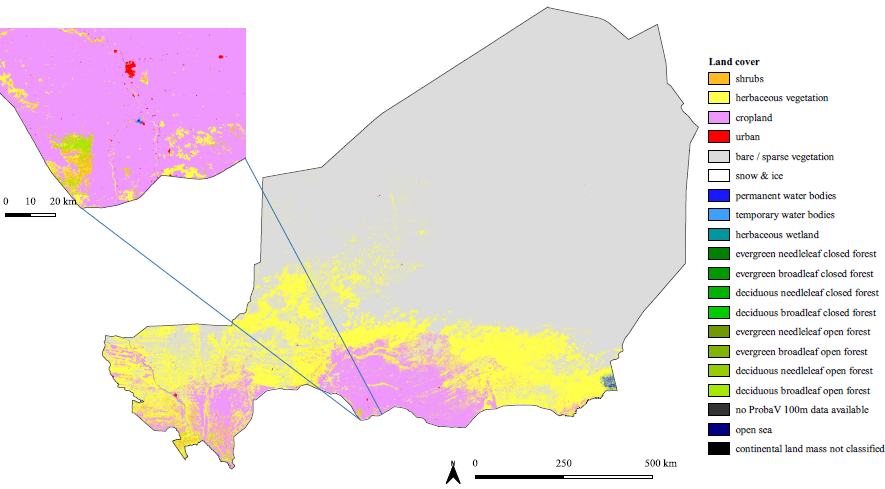

**Supplementary Figure 16. Land cover at 100m x 100m resolution.**
Land cover at 100m x 100m and 1km x 1km resolutions (latter not shown). Discreet land cover classes are based on the UN Land Cover Classification System (LCCS). Source: Buchhorn, M., Smets, B., Bertels, L., Lesiv, M., Tsendbazar, N.-E., Masiliunas, D., Herold, M., Fritz, S. (2019). Copernicus Global Land Service: Land Cover 100m, Collection 2, epoch 2018 Africa Demo (Version V2.1.1). Zenodo. DOI:10.5281/zenodo.3518087. Copernicus Global Land Service, accessed on 27 March 2018 at <https://land.copernicus.eu/global/products/lc>.^9^

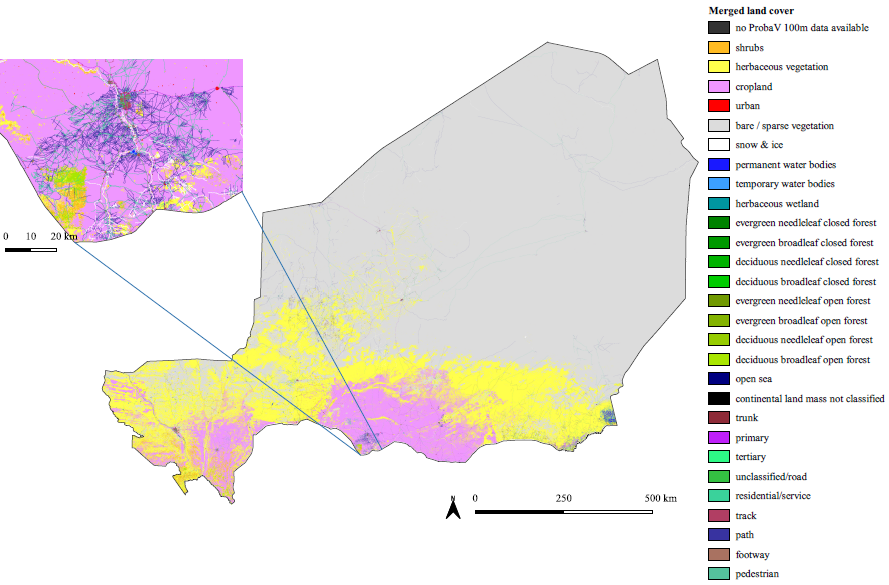

**Supplementary Figure 17. Merged land cover at 100m x 100m resolution.**
Merged land cover at 100m x 100m and 1km x 1km resolutions (latter not shown) derived using the “Merge land cover” tool in AccessMod v5^10^. Road classes “construction” and “bridleway” not shown due to space limitations.

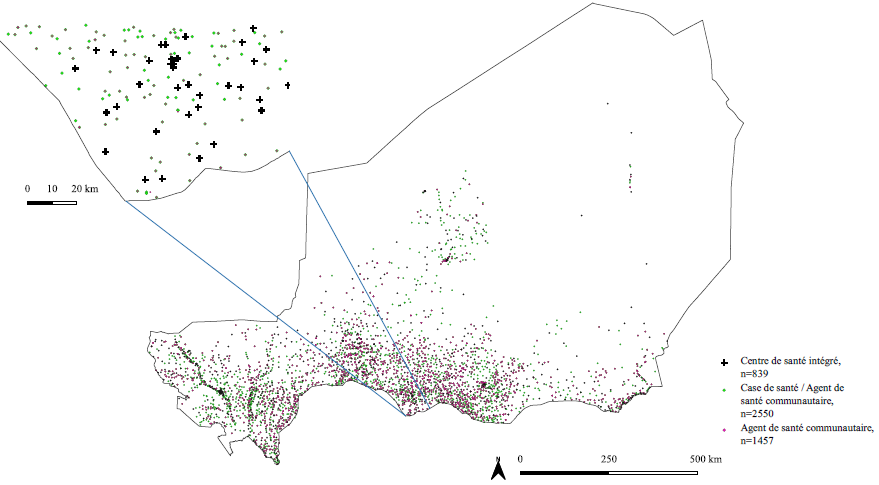

**Supplementary Figure 18. Health service delivery locations**
Source: UNICEF, Institut National de la Statistique, Ministère de la Santé Publique. 2013 Census of Agents de Santé Communautaire, Cases de Santé and Centres de Santé Intégré.^11^

**Data**

**Administrative boundaries**

We obtained vector shapefiles for administrative boundaries 0-3 developed by the Institut Géographique National Niger (IGNN) and OCHA in 2017 with updates from the REACH Initiative in 2018, accessed 14 February 2018, at https://data.humdata.org/dataset/niger-administrative-boundaries.^12^ We reprojected the shapefiles for the administrative boundaries 0-3 from the Coordinate Reference System (CRS) EPSG:4326, WGS 84 to the CRS EPSG:32632 - WGS 84 / UTM zone 32N using the GDAL “Warp” tool in QGIS 3.12.0-Bucareşti.^13^

**Health system pyramid and health service delivery networks mapped**

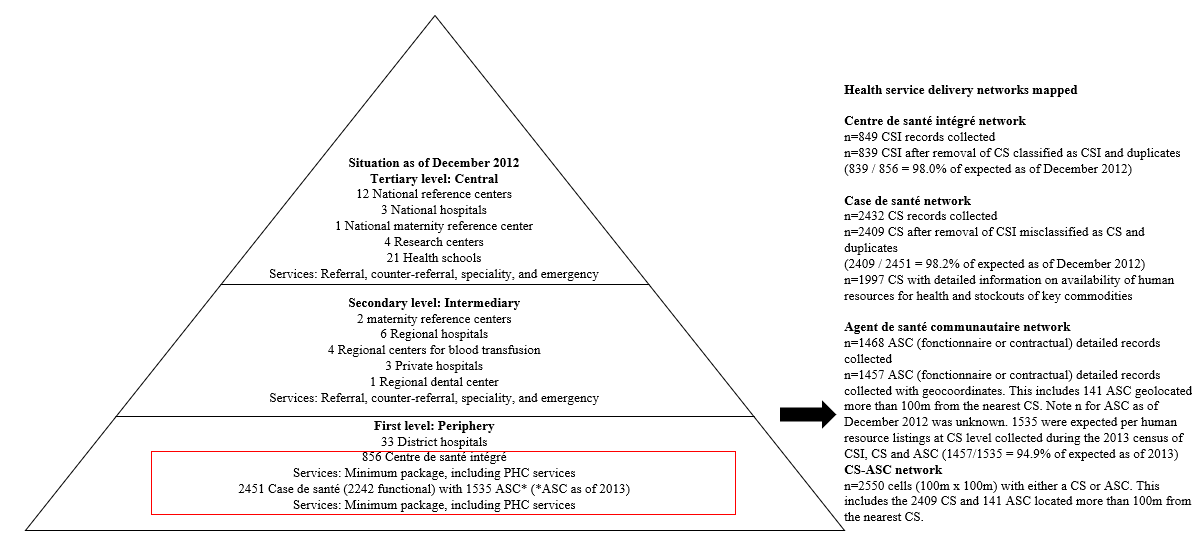

**Supplementary Figure 19. Health system pyramid and health service delivery networks mapped**
The health system of Niger included a public and private sector organized in a decentralized, pyramidal structure with three administrative levels: a central level composed of the cabinet of the Minister of Public Health, the Secretary General and General/National Directorates, responsible for strategy and managing national hospitals, and national maternities and referral centers; a regional level composed of Regional Directorates, responsible for managing regional hospitals, and regional maternities and referral centers; and a district level, composed of District Health Teams, responsible for managing district hospitals, a network of first-level health facilities called *centre de santé intégré* (CSI), a network of community health posts called *case de santé* (CS) – attached to the network of CSI – as well as a small network of private clinics and practices.^14^

Structures at the central and regional levels, as well as district hospitals provided referral, counter referral, specialist, and emergency services not available at the peripheral level through the *Centre de Santé Intégré* (CSI) and *Case de Santé* (CS) networks.^15^ As of December 2012, there were 856 CSI, offering a minimum package of services, focused on primary health care, referral from and counter-referral to the CS, and supervision of the CS. CSI were typically staffed by nurses and in certain large communes by a generalist doctor and midwives.^14^ According to national norms, CSI in rural areas (CSI Type I) serve a maximum population of 10000 and a maximum population of 5000 in rural areas with low population density, while CSI in urban areas or areas with high population density (CSI Type II) serve a maximum population of 15000.^15^ As of December 2012, there were 2451 CS.^14^ According to national norms, CS were attached to the CSI in the hierarchy of the health system, were intended to be situated beyond 5km from a CSI, and served a population of 2500 to 5000.^15^ CS provided a minimum package of activities, focused on primary health care: case management for common infectious diseases, including acute respiratory illness, diarrhea, and malaria, referral services for severe or complicated cases, reproductive health services (family planning, antenatal care, assisted delivery and referral for pregnancies at elevated risk or with complications) and health promotion.^15^ CS typically were staffed by a cadre of full-time *agent de santé communautaire* (ASC), community health workers, who were typically contracted, paid a monthly salary of roughly $100 USD, had completed at least secondary education, and received a six-month pre-deployment basic training on the minimum package and a six-day training on iCCM after deployment.^11,15—17^ ASC typically provided services from the CS (i.e. fixed site service delivery) and did not typically provide mobile services or household visits. In 2013, there were 1535 ASC (1154 male and 381 female).^11^ In addition 21.6% of CS had at least one nurse in 2013 (232 nurses were deployed at CS in 2013) and 42.0% of CS had at least one *relais communautiare* (RC) – a network of volunteer community health workers attached to the CS and providing health promotion and prevention interventions in the communities within the catchment area of the CS (2672 RC were supporting CS in 2013).^11^

**Centre de santé intégré network**

Through a data sharing agreement with UNICEF, we obtained a vector point shapefile dataset in the CRS EPSG:4326, WGS 84 with the global positioning system (GPS) coordinates and basic identification information for all CSI (n=849) in Niger collected through a national, georeferenced census of CSI, CS and ASC conducted in 2013 by the National Institute of Statistics of Niger (INS), Ministry of Public Health (MoPH) of Niger, and UNICEF.^11^ We found that 10 records were misclassified as CSI and these were removed, leaving 839 CSI. We triangulated the CSI dataset with the CS dataset (below) to ensure no duplication or misclassification of CSI as CS and *vice versa*. We reprojected the CSI shapefile to the CRS EPSG:32632 – WGS 84 / UTM zone 32N, using the GDAL “Warp” tool in QGIS 3.12.0-Bucareşti.^13^ For our analysis of geographic coverage, the maximum population capacity of a CSI was set at a population of 10000 for both CSI Type I CSI Type II to simplify the analysis and because we deemed a maximum population capacity of 15000 for CSI in urban areas (as noted above) unrealistic.

**Case de santé network**

We obtained, through the data sharing agreement with UNICEF noted above, a vector point shapefile dataset in the CRS EPSG:4326, WGS 84 with the GPS coordinates and basic identification information for 2409 CS in Niger collected through a national, georeferenced census of CSI, CS and ASC conducted in 2013 by the INS of Niger, MoPH of Niger, and UNICEF.^11^ Data was collected for 2432 structures (n=1703 structures with geocoordinates and complete interviews with the responsible health agent, n=294 structures with geocoordinates and partially complete interviews with the responsible health agent, n=3 structures with geocoordinates where the responsible agent declined to be interviewed, n=3 structures with geocoordinates but missing interviews with the responsible agent, and n=429 structures with geocoordinates that were closed at the time of the census or the responsible agent was absent. We excluded 23 CS records due to miscoding of CSI as CS, leaving 2409 CS records. In our analysis we included all CS records (n=2409) with geocoordinates, including those that were closed at the time of the census or the agent was absent – with the understanding that closures of CS and absences of responsible agents are typically temporary and vary from year to year^11,18^. We reprojected the CS shapefile to the CRS EPSG:32632 – WGS 84 / UTM zone 32N, using the GDAL “Warp” tool in QGIS 3.12.0-Bucareşti.^13^ For analyses at 1km resolution (geographic coverage, targeting and scale-up analysis) we adjusted GPS coordinates, where necessary, for barriers at 1km resolution – these changes were maintained for analyses at 100m resolution (geographic accessibility). Detailed data on the availability of human resources for health and stockouts was available for a subset (n=1997) of the 2409 CS and our analysis of geographic accessibility to CS with available professional/trained human resources for health (e.g. had a nurse – registered nurse, certified nurse, state registered nurse, senior nursing technician – or ASC) and CS without severe stockouts of key commodities for case management of malaria, pneumonia and diarrhea was based on this subset of the CS data. A “severe stockout” was defined as a stockout lasting seven days or longer. We considered key commodities for the case management of malaria (rapid diagnostic test, Artemether/lumefantrine 20/120 mg) pneumonia (cotrimoxazole in pill or syrup form), diarrhea (low osmolarity oral rehydration salt sachets and zinc sulfate 20 mg) and acute malnutrition (ready-to-use therapeutic food (RUTF)). For our analysis of geographic coverage, the maximum population capacity of a CS was set at a population of 2500.

**ASC network**

In 2013, there were 1535 ASC (1154 male and 381 female).^11^ We obtained, through the data sharing agreement with UNICEF noted above, a vector point shapefile dataset in the CRS EPSG:4326, WGS 84 with GPS coordinates of the work location of the ASC and detailed information for 1468 ASC (95.6% of the 1535 expected ASC) from 1421 CS, including socio-demographics, year of deployment, initial training and refresher training for specific interventions collected through a national, georeferenced census of CSI, CS and ASC conducted in 2013 by the INS of Niger, MoPH of Niger, and UNICEF.^11^ We found 11 ASC without GPS coordinates and excluded them from analysis, leaving 1457 ASC (94.9% of the 1535 expected ASC). We reprojected the ASC shapefile to the CRS EPSG:32632 – WGS 84 / UTM zone 32N, using the GDAL “Warp” tool in QGIS 3.12.0-Bucareşti.^13^ For analyses at 1km resolution (geographic coverage, targeting and scale-up analysis) we adjusted GPS coordinates, where necessary, for barriers at 1km resolution – these changes were maintained for analyses at 100m resolution (geographic accessibility).

We prepared separate vector point files using CRS EPSG:32632 – WGS 84 / UTM zone 32N for the ASC network, according to gender of the ASC, year of deployment and training on iCCM. We found that 1316 (90.4%) of ASC were located at the CS to which they were attached but 141 (9.6%) had unique GPS coordinates greater than 100m from the nearest CS. For our analysis of geographic coverage, the maximum population capacity of an ASC was set at a population of 2500. For ASC based at a CS, the maximum population capacity was maintained at 2500 (i.e. they were considered as contributors to the maximum population capacity of the CS).

**CS-ASC network**

We prepared a vector point shapefile with CRS EPSG:32632 – WGS 84 / UTM zone 32N that combined the CS (n=2409) and ASC with unique GPS coordinates (n=141) into a single CS-ASC network (n=2550). For our analysis of geographic coverage, the maximum population capacity of a CS-ASC was set at a population of 2500.

**Optimized CS-ASC network**

For our targeting analysis, we prepared three vector point shapefiles for hypothetical CS-ASC networks: 1) optimizing geographic coverage of the estimated residual population in 2013 beyond the geographic coverage of the CSI network (60-minutes walking considering maximum population capacity) 2) optimizing geographic coverage of the estimated residual under-five deaths in 2013 beyond the geographic coverage of the CSI network and 3) optimizing geographic coverage of the estimated residual *Pf* malaria cases among all ages (0-99) in 2013 beyond the geographic coverage of the CSI network to compare against the existing CS-ASC network, given the same number of CS-ASC as the existing CS-ASC network (n=2550), at 1km x 1km resolution. The optimized CS-ASC networks were prepared using the following steps:

1. Using the population beyond the geographic coverage of the CSI network -- i.e. the population beyond the 60-minute catchment of the CSI network, with maximum population capacity of 10000 population per CSI, we used the “Raster calculator” in QGIS 3.12.0-Bucareşti^13^ to create a dummy raster containing cells at 1km x 1km resolution with greater than or equal to 500 people. The cut-off of greater than or equal to 500 people was chosen as it reflects 25% of the maximum population capacity (2500 people) of a CS-ASC^16^ and we assumed deployment of CS-ASC to cells with less than 500 people would be contrary to country norms^16^.
2. We vectorized the raster from step 1 using the “Polygonize” tool in QGIS 3.12.0-Bucareşti,^13^ resulting in a point vector shapefile of 5796 potential CS-ASC sites.

See the section below on the Targeting analysis for further details on preparation of these datasets.

**Scaled-up *relais* *communautaire* network**

The MoPH in Niger plans to scale-up the network of CHWs called *relais communautaire* (RC) for two contexts: 1) rural contexts at a ratio of 2 RC per 1000 population in communities beyond 5km of the CS-ASC or CSI networks to provide preventive, promotional and curative (e.g. iCCM) interventions and 2) in urban/peri-urban contexts at a ratio of 1 RC per 1000 population in communities within 5km of the CS-ASC and CSI networks to provide preventive and promotional interventions. For our scale-up analysis, we focus on the former. We prepared a hypothetical “optimized” RC network (n=3295) to cover the population in cells with at least 500 people in 2013 beyond the geographic coverage of the existing CSI and CS-ASC networks at 1km x 1km resolution using the following steps:

1. Using the population beyond the geographic coverage of the existing CS-ASC network – i.e. the population beyond the 60-minute catchment of the CS-ASC network, with maximum population capacity of 2500 population per CS-ASC we used the “Raster calculator” in QGIS 3.12.0-Bucareşti^13^ to create a dummy TIFF raster containing cells at 1km x 1km resolution with greater than or equal to 500 people.
2. We vectorized the raster from step 1 using the “Polygonize” tool in QGIS 3.12.0-Bucareşti,^13^ resulting in a point vector shapefile of 3521 candidate RC sites, with 7042 RC at a ratio of 2 RC per site or 2 RC per 1000 people based on the national norm.^16^
3. In our scale-up analysis (described below) we filtered out candidate sites with a realized capacity (i.e., the population covered within the catchment) of less than 500 population, leaving 3296 candidate sites for the scale-up analysis.

**DEM**

We obtained 174 tiles of a gridded digital elevation model (DEM) – the NASA Shuttle Radar Topography Mission Global 1 arc second (SRTMGL1) dataset version 3.0, with a resolution of approximately 30 meters (m) x 30m (0.000277778 decimal degrees) for the area including Niger.^1^ The SRTMGL1 was retrieved 4 October 2017 from the online EarthExplorer, courtesy of the NASA EOSDIS Land Processes Distributed Active Archive Center (LP DAAC), USGS/Earth Resources Observation and Science (EROS) Center, Sioux Falls, South Dakota, https://earthexplorer.usgs.gov/. More information on the SRTMGL1 is available at https://lpdaac.usgs.gov/node/527. We used the “merge” function in QGIS 3.12-Bucareşti^13^ to mosaic the original tiles into one georeferenced Tagged Information File Format (GeoTIFF) raster. For our analysis at 100m x 100m resolution (geographic accessibility analysis) we prepared a DEM raster at 100m x 100m resolution using the GDAL “warp” tool in QGIS 3.12.0-Bucareşti^13^ to reproject the CRS of the original file from EPSG:4326, WGS 84 to the CRS EPSG:32632 - WGS 84 / UTM zone 32N, resample the resolution to 100m x 100m using bilinear as the resampling method and clip the file to the extent of the administrative level 3 (adm3) shapefile (see GeoTIFF file “r_NER_DEM_100m_final.tif” in Supplementary Appendix 3). For our analysis at 1km x 1km resolution (geographic coverage, targeting and scale-up analysis) we prepared a GeoTIFF DEM raster at 1km x 1km resolution using the GDAL “warp” tool in QGIS 3.12.0-Bucareşti^13^ and the process described above (see the GeoTIFF file “r_NER_DEM_1km_final.tif” in Supplementary Appendix 1c at [https://doi.org/10.6084/m9.figshare.13536779.v5](https://doi.org/10.6084/m9.figshare.13536779)).

**Land cover**

We obtained a GeoTIFF raster for land cover in Africa [c_gls_LC100-LCCS_201501010000_AFRI_PROBAV_1.0.1] at a resolution of approximately 100m x 100m from the Copernicus Global Land Service,^9^ accessed on 27 March 2018 at <https://land.copernicus.eu/global/products/lc>. The land cover dataset contains discreet land cover classes based on the UN Land Cover Classification System (LCCS). Further details on the Copernicus land cover data set are available at <https://land.copernicus.eu/global/products/lc>. For our analysis at 100m x 100m resolution (geographic accessibility analysis) we prepared a GeoTIFF land cover raster (see the GeoTIFF file “r_NER_land_100m_final.tif” in Supplementary Appendix 3) using the GDAL "warp" tool in QGIS 3.12.0-Bucareşti^13^ to reproject the CRS from EPSG:4326 - WGS84 to EPSG:32632 - WGS 84 / UTM zone 32N, resample the resolution to 100m x 100m using nearest neighbor as the sampling method and clip the file to the extent of the final DEM. For our analysis at 1km x1km resolution (geographic coverage, targeting and scale-up analysis) we prepared a GeoTIFF land cover raster at 1km x 1km resolution using the GDAL “warp” tool in QGIS 3.12.0-Bucareşti^13^ and the process described above (see the GeoTIFF file “r_NER_land_1km_final.tif” in Supplementary Appendix 1c at [https://doi.org/10.6084/m9.figshare.13536779.v5](https://doi.org/10.6084/m9.figshare.13536779)).

**Roads**

We obtained a vector line shapefile for the road network in Niger developed by the Humanitarian OpenStreetMap Team, accessed on 1 August 2018, at https://data.humdata.org/dataset/hotosm_niger_roads.^7^ To prepare the final roads file, we changed the column “Highway” to “label”; reclassified the road types using the standard OpenStreetMap categories described at <https://wiki.openstreetmap.org/wiki/Key:highway>; simplified the road typology by excluding road types with very few segments or of little importance/relevance to the study; added a “class” variable in order to enable linking with the travel time scenarios; and reprojected the CRS from EPSG:4326 - WGS84 to EPSG:32632 - WGS 84 / UTM zone 32N in alignment with the final DEM (see files t_NER_reclass_roads_OSM.xls, v_NER_roads_100m_final.shp and v_NER_roads_1km_final.shp in Supplementary Appendix 1c at [https://doi.org/10.6084/m9.figshare.13536779.v5](https://doi.org/10.6084/m9.figshare.13536779)). As described below in the section on the merged land cover raster, for our analysis at 100m x 100m resolution we uploaded the vector line shapefile for the road network into our Accessmod v5 project at 100m x 100m resolution and used the merge land cover tool in Accessmod v5 to rasterize the vector line shapefile for the road network as part of the merged land cover raster at 100m x 100m resolution. For our analysis at 1km x 1km resolution (geographic coverage, targeting and scale-up analysis) we repeated the above within our Accessmod v5^10^ project at 1km x 1km resolution.

**Rivers and Other Waterbodies**

Rivers and other waterbodies were considered barriers to movement, where they were not crossed by a road. We obtained vector line shapefiles for rivers from HOT Open Street Map (HOTOSM), accessed on 15 January 2018, at <https://data.humdata.org/dataset/hotosm_niger_waterways>.^8^ For our analysis at 100m x100m resolution (geographic accessibility), we reprojected the CRS from EPSG:4326 - WGS84 to CRS EPSG:32632 - WGS 84 / UTM zone 32N in alignment with the final DEM (see file “v_NER_rivers_final.shp” in Supplementary Appendix 1c at [https://doi.org/10.6084/m9.figshare.13536779.v5](https://doi.org/10.6084/m9.figshare.13536779)). As described below in the section on the merged land cover raster, for our analysis at 100m x 100m resolution we uploaded the vector line file for rivers into our Accessmod v5^10^ project at 100m x 100m resolution and used the merge land cover tool in Accessmod v5^10^ to rasterize the vector line shapefile for rivers as part of the merged land cover raster at 100m x 100m resolution. For our analysis at 1km x 1km resolution (geographic coverage, targeting and scale-up analysis) we repeated the above within our Accessmod v5^10^ project at 1km x 1km resolution. Data on other water bodies (permanent and temporary) were already included as part of the land cover raster described above.

**Merged land cover**

For our geographic accessibility analysis, we prepared a merged land cover raster at 100m x 100m resolution using the “Merge land cover” tool in AccessMod v5^10^ (see file “r_NER_land_merged_100m_final.tif” in Supplementary Appendix 1c at [https://doi.org/10.6084/m9.figshare.13536779.v5](https://doi.org/10.6084/m9.figshare.13536779)). The process is described in detail in Ray et al, 2008.^10^ In brief, the “Merge land cover” tool stacks, orders, and merges the road network, barriers (rivers and other waterbodies, the later from the land cover), and land cover files into a single raster dataset. For our analysis at 1km x 1km resolution (geographic coverage, targeting and scale-up analysis) we prepared a merged land cover raster at 1km x 1km resolution using the process described above within our Accessmod v5^10^ project at 1km x 1km resolution (see the file “r_NER_land_merged_100m_final.tif”).

**Travel scenario tables**

We developed travel scenario tables for the following scenarios walking in dry conditions and walking to the nearest road and then using motorized transportation in dry conditions (see files “t_NER_walk_dry.xls” and “t_NER_walk_veh_dry.xls” in Supplementary Appendix 1c at [https://doi.org/10.6084/m9.figshare.13536779.v5.v2](https://doi.org/10.6084/m9.figshare.13536779.v2)). We set traveling speeds by mode of transportation (walking or walking + motorized transportation) for each land cover class and road class. Travel speeds were adapted from previous studies.^18,19^

**Population**

**Data preparation of population raster layers for the year 2013**

We obtained a GeoTiff raster for the estimated population count for Niger in 2015 adjusted to UN population estimates at roughly 30m x 30m resolution, the High Resolution Settlement Layer (HRSL) from <https://data.humdata.org/dataset/highresolutionpopulationdensitymaps-ner>, courtesy of Facebook Connectivity Lab and Center for International Earth Science Information Network (CEISIN) at Columbia University, accessed 6 August 2020.^2^ The 2015 HRSL was developed with computer vision techniques and supervised machine learning applied to high resolution commercial satellite imagery from the DigitalGlobe, courtesy of Maxar^20^ to identify and classify human-built structures, combined with population estimates from the Gridded Population of the World v4^21^. Further details are provided elsewhere^22^. We also obtained a GeoTiff raster for the estimated population count for Niger in 2013, adjusted to UN population estimates, at roughly 100m x 100m resolution in Geographic Coordinate system WGS84 from Worldpop, accessed 3 March 2020.^3^ A random forest-based dasymetric redistribution approach was used to develop the Worldpop dataset and is described in detail elsewhere.^23^ We prepared a GeoTiff raster file for the estimated population count in 2013 at 100m x 100m resolution that adjusted the HRSL GeoTiff of the estimated population count in 2015 to the GeoTiff from Worldpop for the estimated population count in 2013 [ner_ppp_2013] at the lowest administrative level (adm3) but maintained the population settlement footprint of the 2015 HRSL. We kept the footprint of the 2015 HRSL because we deemed it more appropriate for our purposes (analysis of geographic accessibility to health services) than the footprint of the Worldpop raster based on visual inspection against satellite imagery for Niger and recent assessments of its accuracy^23^. The population footprint of the Worldpop raster is “unconstrained”, that is, smoothed across space,^24^ including cells where there are no settlements,^25^ whereas the HRSL is confined to cells with settlements.^2^ We note that since the time of our analysis, an additional dataset – the World Settlement Footprint 2015^26^– has been made publicly available and Worldpop has developed population count datasets constrained to population settlement footprints.^27^ Recent analyses suggest that modelling of population counts constrained to settlement footprints is improved through the use of multiple settlement footprints.^28,29^

We adjusted the counts of the 2015 HRSL dataset to the Worlpop counts for 2013 to align with our analysis for the year 2013. The raster layer for the population count in the year 2013 at 100m x 100m resolution was used in our analysis of geographic accessibility, geographic coverage and scale-up. We used the following steps to prepare the raster layer for the population count in 2013 at 100m x 100m resolution:

1. We reprojected the original HRSL GeoTiff raster file for the population count in 2015 at approximately 30 meter resolution [population_ner_2018-10-01] from the CRS EPSG:4326 - WGS84 to the CRS EPSG:32632 – WGS 84 / UTM zone 32N using the GDAL Warp tool in QGIS 3.12.0-Bucareşti^13^ and aggregated the reprojected raster to 100 meter resolution using the r.resamp.stats GRASS 7.8.2 plugin in QGIS 3.12.0-Bucareşti,^13^ with sum as the aggregation method and the final DEM at 100 meter resolution as the extent, resulting in the file [r_NER_FB15N_unadj_100m].
2. We used the “Zonal statistics” tool in QGIS 3.12.0-Bucareşti^13^ to calculate the count of the population from the original World pop population layer in 2013 [ner_ppp_2013] to a vector file for administrative level 3 in CRS WGS84 and used a spatial join to copy the population counts to the vector file for administrative level 3 in CRS EPSG:32632 – WGS 84 / UTM zone 32N [v_NER_adm3_final].
3. We used the “Zonal statistics” tool in QGIS 3.12.0-Bucareşti^13^ to calculate the count of the population from the raster of the 2015 population at 100 meters [r_NER_FB15N_unadj_100m] from step 1 to the vector file [v_NER_adm3_final].
4. We created a ratio called “WP13tFB15” in the adm3 vector file [v_NER_adm3_final] that divided the population count at administrative level 3 from the original World pop population layer in 2013 from step 2 [ner_ppp_2000] by the population count at administrative level 3 from the HRSL population layer in 2015 from step 1 [r_NER_FB15N_unadj_100m].
5. We rasterized this ratio at 100m resolution using the “Rasterize” tool in QGIS 3.12.0-Bucareşti^13^ [r_NER_ratWP13OtFB15N_100m] with the ratio from step 4 as the burn and the extent of the DEM at 100m resolution as the extent. Using raster calculator in QGIS 3.12.0-Bucareşti^13^, we multiplied the rasterized ratio [r_NER_ratWP13OtFB15N_100m] by the HRSL population in 2015 at 100m x 100m resolution [r_NER_FB15N_unadj_100m] to create a GeoTiff raster for the population in the year 2013 [r_NER_FB13_100m_unadj_barriers].
6. We uploaded the file [r_NER_FB13_100m_unadj_barriers] into Accessmod v5 and redistributed the population on cells with barriers to cells without barriers within the same administrative level 3 boundaries, resulting in the final raster file for the population in the year 2013 [raster_population_r_NER_FB13_100m_final].

We repeated the steps above at 1km x 1km resolution to produce the GeoTiff raster of the population in 2013 at 1km x 1km resolution [r_NER_FB13_1km_final] (see Supplementary Appendix 1c at [https://doi.org/10.6084/m9.figshare.13536779.v5](https://doi.org/10.6084/m9.figshare.13536779)).

**Data preparation of population raster layers for the years 2000-2012**
We prepared a GeoTiff raster layer for the population count in the year 2000 at 100m x100m resolution that matched the population count from the original Worldpop GeoTiff raster layer in 2000 [ ner_ppp_2000] at the lowest administrative level (adm3) but maintained the population settlement footprint of the 2015 HRSL. This assumes the actual population settlement footprint in 2000 would be similar to the HRSL, a limitation we acknowledge in the section on limitations. We used the raster layer for the population in the year 2000 to generate zonal statistics by administrative level for the estimated number and percent of the population in 2000 within 30 minutes and 60 minutes of the nearest ASC in 2000. We used the following steps to prepare the raster layer for the population count in 2000 at 100m x 100m resolution:

1. We reprojected the original Worldpop GeoTiff raster layer for the population in 2000 at approximately 90m x 90m resolution [ner_ppp_2000] from the CRS EPSG:4326 - WGS84 to the CRS EPSG:32632 – WGS 84 / UTM zone 32N using the GDAL Warp tool in QGIS 3.12.0-Bucareşti^13^ and aggregated the reprojected raster to 100m x 100m meter resolution using the r.resamp.stats GRASS 7.8.2 plugin in QGIS 3.12.0-Bucareşti,^13^ with sum as the aggregation method and the final DEM at 100m x 100m resolution as the extent, resulting in the file [r_NER_FB00_100m_unadj_barriers].
2. We used the “Zonal statistics” tool in QGIS 3.12.0-Bucareşti^13^ to calculate the count of the population from the original World pop population layer in 2000 [ner_ppp_2000] to a vector file for administrative level 3 in CRS WGS84 and used a spatial join to copy the population counts to the vector file for administrative level 3 in CRS EPSG:32632 – WGS 84 / UTM zone 32N [v_NER_adm3_final].
3. We created a ratio called “WP00tFB13” in the adm3 vector file [v_NER_adm3_final] that divided the count from the original Worldpop population layer for 2000 from step 2 [ner_ppp_2000] by the population count from the population layer for 2013 [raster_population_r_NER_FB13_100m_final], which as described above, maintains the population settlement footprint of the 2015 HRSL.
4. We rasterized this ratio at 100m resolution using the “Rasterize” tool in QGIS 3.12.0-Bucareşti^13^ [r_NER_ratWP00OtFB13F_100m_unadj_barriers] with the ratio from step 3 as the burn and the extent of the DEM at 100m x 100m resolution as the extent. Using raster calculator in QGIS 3.12.0-Bucareşti,^13^ we multiplied the rasterized ratio [r_NER_ratWP00OtFB13F_100m_unadj_barriers] by the 2013 population [raster_population_r_NER_FB13_100m_final] to create a raster for the population in the year 2000 [r_NER_FB00_100m_unadj_barriers]. This approach effectively maintained the spatial distribution of the population as in 2013 while adjusting the 2013 population count downward to match the population from Worldpop for the year 2000 at the lowest administrative level [v_NER_adm3_final].
5. We uploaded the file [r_NER_FB00_100m_unadj_barriers] into Accessmod v5 and redistributed the population on cells with barriers to cells without barriers within the same administrative level 3 boundaries, resulting in the final raster file for the population in the year 2000 [raster_population_r_NER_FB00_100m_final].

For the years 2001-2012, we repeated the steps taken above for the year 2000 using the appropriate input population layers from Worldpop to create the rasterized ratios for each year:

2001: input file from Worldpop [ner_ppp_2001]; rasterized ratio file [r_NER_ratWP01OtFB13F_100m_unadj_barriers]
2002: input file from Worldpop [ner_ppp_2002]; rasterized ratio file [r_NER_ratWP02OtFB13F_100m_unadj_barriers]
2003: input file from Worldpop [ner_ppp_2003]; rasterized ratio file [r_NER_ratWP03OtFB13F_100m_unadj_barriers]
2004: input file from Worldpop [ner_ppp_2004]; rasterized ratio file [r_NER_ratWP04OtFB13F_100m_unadj_barriers]
2005: input file from Worldpop [ner_ppp_2005]; rasterized ratio file [r_NER_ratWP05OtFB13F_100m_unadj_barriers]
2006: input file from Worldpop [ner_ppp_2006]; rasterized ratio file [r_NER_ratWP06OtFB13F_100m_unadj_barriers]
2007: input file from Worldpop [ner_ppp_2007]; rasterized ratio file [r_NER_ratWP07OtFB13F_100m_unadj_barriers]
2008: input file from Worldpop [ner_ppp_2008]; rasterized ratio file [r_NER_ratWP08OtFB13F_100m_unadj_barriers]
2009: input file from Worldpop [ner_ppp_2009]; rasterized ratio file [r_NER_ratWP09OtFB13F_100m_unadj_barriers]
2010: input file from Worldpop [ner_ppp_2010]; rasterized ratio file [r_NER_ratWP10OtFB13F_100m_unadj_barriers]
2011: input file from Worldpop [ner_ppp_2011]; rasterized ratio file [r_NER_ratWP11OtFB13F_100m_unadj_barriers]
2012: input file from Worldpop [ner_ppp_2012]; rasterized ratio file [r_NER_ratWP12OtFB13F_100m_unadj_barriers]

We repeated step 3 above (redistribution of the population on cells with barriers to cells without barriers in Accessmod v5) for the 2001-2012 datasets, resulting in the following final population layers for the years 2001-2012 at 100m x 100m resolution to be used in our analysis of the trends in geographic accessibility between 2000-2013 (see Supplementary Appendix 1c at [https://doi.org/10.6084/m9.figshare.13536779.v5.v2](https://doi.org/10.6084/m9.figshare.13536779.v2)):

2001: [ raster_population_r_NER_FB01_100m_final]
2002: [raster_population_r_NER_FB02_100m_final]
2003: [raster_population_r_NER_FB03_100m_final]
2004: [raster_population_r_NER_FB04_100m_final]
2005: [raster_population_r_NER_FB05_100m_final]
2006: [raster_population_r_NER_FB06_100m_final]
2007: [raster_population_r_NER_FB07_100m_final]
2008: [raster_population_r_NER_FB08_100m_final]
2009: [raster_population_r_NER_FB09_100m_final]
2010: [raster_population_r_NER_FB10_100m_final]
2011: [raster_population_r_NER_FB11_100m_final]
2012: [raster_population_r_NER_FB12_100m_final]

**Data preparation of under-five population raster layer for the year 2013**
We prepared a GeoTiff raster layer for the count of the under-five population in 2013 at 1km x 1km resolution to be used in our targeting analysis for under-five deaths. We used the following steps:

1. We reprojected the original 2015 HRSL GeoTiff raster layer for the count of the under-five population in 2016 [ NER_children_under_five] from the CRS EPSG:4326 - WGS84 to the CRS EPSG:32632 – WGS 84 / UTM zone 32N using the GDAL Warp tool in QGIS 3.12.0-Bucareşti^13^ and aggregated the reprojected raster to 1km x 1km resolution using the r.resamp.stats GRASS 7.8.2 plugin in QGIS 3.12.0-Bucareşti,^13^ with sum as the aggregation method and the final DEM at 1km x1km resolution as the extent, resulting in the file [r_NER_U5FB15_unadj_1km].
2. We used zonal stats to calculate the sum of the count of the under-five population from the original 2015 HRSL GeoTiff raster by administrative level 3 and created a ratio called “FBU5OtN” in the adm3 vector file [v_NER_adm3_final] that divided the original HRSL under-five population count in 2015 [NER_children_under_five] by the sum of the new under-five population count from step 1 above [r_NER_U5FB15_unadj_1km].
3. We rasterized this ratio at 1km x 1km resolution using the “Rasterize” tool in QGIS 3.12.0-Bucareşti^13^ [r_NER_ratU5FB15tFB15_1km] with the ratio from step 2 as the burn and the extent of the DEM at 1km x 1km meter resolution as the extent. Using raster calculator in QGIS 3.12.0-Bucareşti,^13^ we multiplied the rasterized ratio [r_NER_ratU5FB15tFB15_1km] by the 2013 population count [raster_population_r_NER_FB13_100m_final] to create a raster for the count of the under-five population in the year 2013 [r_NER_U5FB13_1km_unadj_barriers]. This approach effectively maintained the spatial distribution of the population layer for 2013, which was based on the population settlement footprint of the 2015 HRSL, while using the ratio of the count of the under-five population in 2015 to the count of the total population in 2015 as a means to adjust the count of the total population in 2013 downward to derive the count of the under-five population in 2013.
4. We uploaded the file [r_NER_U5FB13_1km_unadj_barriers] into Accessmod v5 and redistributed the population on cells with barriers to cells without barriers within the same administrative level 3 boundaries, resulting in the final raster file for the population in the year 2013 at 1km resolution [raster_population_r_NER_U5FB13_1km_final] (see Supplementary Appendix 1c at [https://doi.org/10.6084/m9.figshare.13536779.v5](https://doi.org/10.6084/m9.figshare.13536779)).

**Estimated under-five deaths**

We used the following steps to prepare the raster layer for the estimated count of under-five (0-5 years old) deaths in Niger in 2013 at 1km x 1km resolution to be used in our targeting analysis:

1. We obtained a GeoTiff raster file for modelled pixel-level estimates of the mean probability of under-five (0-5 years old) mortality at 2.5 arcminutes (approximately 5km x 5km) resolution developed by the Institute for Health Metrics and Evaluation (IHME),^4,5^ accessed on 3 March 2020, at <http://ghdx.healthdata.org/lbd-data> [IHME_LMICS_U5M_2000_2017_Q_UNDER5_MEAN].
2. We used the “Raster calculator” tool in QGIS 3.12.0-Bucareşti^13^ to prepare a GeoTiff raster for the estimated count of under-five deaths in 2013 at 1km x 1km resolution [r_NER_U5d13_1km] by multiplying the raster for the mean probability of under-five mortality in 2013 (band 14) at 5km x 5km resolution [IHME_LMICS_U5M_2000_2017_Q_UNDER5_MEAN] by the estimated count of the under-five population in 2013 [raster_population_r_NER_U5FB13_1km_final], with the CRS EPSG:32632 – WGS 84 / UTM zone 32N and the final DEM at 1km x 1km as the extent. Given the 5km x 5km resolution of the raster for the mean probability of under-five mortality, each 1km x1km cell belonged to a block of 25 (5 x 5) 1km x 1km cells with the same mean probability of under-five mortality. In the calculation above, the corresponding value for the mean probability of under-five mortality of the 5km x5km area to which a given 1km x 1km cell belonged was multiplied by the estimated count of the under-five population for the given 1km x 1km cell.
3. We used the “Raster calculator” tool in QGIS 3.12.0-Bucareşti^13^ to prepare a GeoTiff raster for the estimated count of residual under-five deaths in 2013 beyond the geographic coverage (1hr catchment, considering capacity) of the existing CSI network at 1km x 1km resolution [ r_NER_residU5d13_gcCSI_60min_1km] by multiplying the estimated count of the under-five deaths in 2013 from step 2 above [r_NER_U5d13_1km] by a dummy mask for the area with non-zero residual population beyond the geographic coverage of the existing CSI network in 2013 [r_NER_raster_popuation__residual_r_NER_gcCSI_60min_1km_prioritize_FB13TT_g0] (see Supplementary Appendix 1c at [https://doi.org/10.6084/m9.figshare.13536779.v5.v2](https://doi.org/10.6084/m9.figshare.13536779.v2)).

Note that we did not need to adjust for the estimated under-five deaths on barriers because this step was conducted when preparing the raster for the estimated population under-five in 2013.

**Estimated *Plasmodium falciparum* malaria cases**

We used the following steps to prepare a GeoTiff raster layer for the estimated count of *Plasmodium falciparum* malaria cases among all ages (0-99 years) in Niger in 2013 at 1km x 1km resolution to be used in our targeting analysis:

1. We obtained a GeoTiff raster file for modelled pixel-level estimates of the annual mean incidence of *Plasmodium falciparum* (*Pf*) malaria among all ages (0-99 years) in 2013 globally at 2.5 arcminutes (approximately 5km x 5km) resolution developed by the Malaria Atlas Project,^6^ accessed on 29 July 2020, at <https://malariaatlas.org/malaria-burden-data-download/> [2019_Global_Pf_Incidence_2013].
2. We used the “Raster calculator” tool in QGIS 3.12.0 to prepare a GeoTiff raster for the count of *Pf* malaria among all ages (0-99 years) in 2013 at 1km x 1km resolution [r_NER_cases13_1km] by multiplying the raster for the mean incidence of *Pf* malaria among all ages in 2013 at 5km x 5km resolution [2019_Global_Pf_Incidence_2013] by the count of the population in Niger in 2013 [raster_population_r_NER_FBpop2013_1km_final ] with the CRS EPSG:32632 – WGS 84 / UTM zone 32N and the final DEM at 1km x 1km as the extent. Given the 5km x 5km resolution of the raster for the mean annual incidence of *Pf* malaria in 2013, each 1km x1km cell belonged to a block of 25 (5 x 5) 1km x 1km cells with the same mean annual incidence. In the calculation above, the corresponding value for the mean annual incidence of the 5km x5km area to which a given 1km x 1km cell belonged was multiplied by the count of the population in 2013 for the given 1km x 1km cell.
3. We used the “Raster calculator” tool in QGIS 3.12.0-Bucareşti^13^ to prepare a GeoTiff raster for the estimated count of residual *Pf* malaria cases in 2013 beyond the geographic coverage (1hr catchment, considering capacity) of the existing CSI network at 1km x 1km resolution [r_NER_residcases13_gcCSI_60min_1km] by multiplying the estimated count of *Pf* malaria cases in 2013 from step 2 above [r_NER_cases13_1km ] by a dummy mask for the area with non-zero residual population beyond the geographic coverage of the existing CSI network in 2013 [r_NER_raster_popuation__residual_r_NER_gcCSI_60min_1km_prioritize_FB13TT_g0] (see Supplementary Appendix 1c at [https://doi.org/10.6084/m9.figshare.13536779.v5](https://doi.org/10.6084/m9.figshare.13536779)).

Note that we did not need to adjust for the estimated *Pf* malaria cases on barriers because this step was conducted when preparing the raster for the estimated population in 2013.

**Analysis**

**Geographic accessibility**

Research questions

1. What was geographic accessibility to the CSI network in 2013?
   a. What percentage of the population was within 30 min and 60 min of a CSI in 2013, assuming a walking scenario in dry conditions? How did this vary across geographies?
   b. What percentage of the population was within 30 min and 60 min of a CSI in 2013, assuming a scenario of walking to the nearest road and then using motorized transportation in dry conditions? How did this vary across geographies?
2. What was geographic accessibility to the CS network in 2013?
   a. What percentage of the population was within 30 min and 60 min of a CS in 2013, assuming a walking scenario in dry conditions? How did this vary across geographies?
   b. What percentage of the population was within 30 min and 60 min of a CS in 2013, assuming a scenario of walking to the nearest road and then using motorized transportation in dry conditions? How did this vary across geographies?
3. What was geographic accessibility to the ASC network?
   a. What percentage of the population was within 30 min and 60 min of an ASC in 2013, assuming a walking scenario in dry conditions? How did this vary across geographies?
   b. What percentage of the population was within 30 min and 60 min of an ASC in 2013, assuming a scenario of walking to the nearest road and then using motorized transportation in dry conditions? How did this vary across geographies?
   c. What was the contribution of ASC to additional geographic accessibility beyond the network of CSI and CS (without ASC) in 2013? Assuming a walking scenario in dry conditions? Assuming a scenario of walking to the nearest road and then using motorized transportation in dry conditions? How did this vary across geographies?
   d. How did geographic accessibility to an ASC evolve over time between 2000-2013? Assuming a walking scenario in dry conditions? Assuming a scenario of walking to the nearest road and then taking motorized transportation in dry conditions?
   e. How did geographic accessibility to an ASC in 2013 differ by gender of the ASC?
   f. What percentage of the population in 2013 was within 30 min and 60 min of an ASC trained on iCCM? Assuming a walking scenario in dry conditions? Assuming a scenario of walking to the nearest road and then using motorized transportation in dry conditions? How did this vary across geographies?
4. What was geographic accessibility to the CS-ASC network in 2013?
   a. What percentage of the population was within 30 min and 60 min of a CS-ASC in 2013, assuming a walking scenario in dry conditions? How did this vary across geographies? How did this vary by availability of trained human resources (nurse, ASC) and essential commodities?
   b. What percentage of the population was within 30 min and 60 min of a CS-ASC in 2013, assuming a scenario of walking to the nearest road and then using motorized transportation in dry conditions? How did this vary across geographies? How did this vary by availability of trained human resources (nurse, ASC) and essential commodities?

**Methods for Geographic Accessibility question 1**

We define accessibility coverage as the estimated percentage of people within a given travel time to the nearest health service delivery location of a given health service delivery network, accounting for travel speeds of different modes of transportation over different land cover classes and slope, with the direction of travel toward the health service delivery location.^10^ We estimated accessibility coverage at 100m x 100m resolution for the CSI, CS and ASC networks in 2013 – and for the ASC network by gender, year of deployment (2000-2013), training, and availability of essential commodities – using 30-minute and 60-minute cutoffs for administrative levels 0-3 and the two travel scenarios. We used 30-minute and 60-minute cutoffs as previous analyses have shown careseeking decays as a function of travel time after these cutoffs^30^ and they are clinically relevant (e.g. for prompt treatment of severe illness).^31^ The analysis was constrained to national borders but allowed for travel across subnational administrative boundaries. We used the “geographic accessibility” module within AccessMod 5 (v5.6.48)^10^ to calculate travel time layers and the “zonal statistics” module to calculate the zonal statistics for each travel time layer by administrative level.

Analysis

1. We conducted a geographic accessibility analysis of the existing CSI network in 2013 based on a travel scenario of walking in dry conditions scenario at 100m x 100m resolution using Accessmod v5.
   a. We used the following data inputs:
      i. Population: raster_population_r_NER_FB13_100m_final
      ii. Land cover merged: raster_land_cover_merged_r_NER_land_merged
      iii. Scenario table: table_scenario_walk_dry
      iv. Select existing health facilities layer (vector): v_NER_CSI
      v. ID field: id
      vi. Facility name field: nom_centre
      vii. Select zones layer (vector): adm3
           1. Select zones unique ID (integer): objectid
           2. Select zone name (text): nom_com
   b. We used the following analysis settings:
      i. Type of analysis: anisotropic
      ii. Direction of travel: towards facilities
      iii. Maximum travel time (minutes): 60
      iv. Options
          1. Compute population catchment area layer: Yes
          2. Remove the covered population at each iteration: Yes
          3. Compute a layer of population cells on barriers: Yes
          4. Generate zonal statistics: Yes (adm 3)
          5. Optimize dynamically computation according to the scenario: Yes
          6. Add short tag: raster_travel_time_r_NER_ga_CSI_wd_100m
2. We repeated steps 1 using a travel scenario for walking to the nearest road, then using motorized transportation in dry conditions. [raster_travel_time_r_NER_ga_CSI_walkvehd_100m] (see Supplementary Appendix 1b at https://doi.org/10.5281/zenodo.4482969).
3. We used the “Zonal statistics” tool within Accessmod v5 to calculate the percent of the population within 30 minutes and 60 minutes travel time in 2013 for the walking in dry conditions scenario and the walking + motorized transportation in dry conditions scenario (see Table 1 and Supplementary Appendix 2).

**Methods for Geographic Accessibility research question 2**

We repeated the analysis described in Methods for Geographic Accessibility question 1, replacing the existing health facilities layer with the layer [v_NER_CS_100m_final]. This resulted in the raster travel time layers [raster_travel_time_r_NER_ga_CS_wd_100m] and [raster_travel_time_r_NER_ga_CS_walkvehd_100m] (see Supplementary Appendix 1b at https://doi.org/10.5281/zenodo.4482969). See Table 1 for zonal statistics from these travel time rasters and detailed results for administrative layers 0-3 in Supplementary Appendix 2.

**Methods for Geographic Accessibility question 3**

We repeated the analysis described in Methods for Geographic Accessibility question 1, as follows:

2000: Input [v_NER_ASC_detailed_ASC_le2000]; output travel time rasters [raster_travel_time_r_NER_ga_ASC_detailed_le2000_wd_100m] and [raster_travel_time_r_NER_ga_ASC_detailed_le2000_walkvehd_100m]

2001: Input [v_NER_ASC_detailed_ASC_le2001]; output travel time rasters [raster_travel_time_r_NER_ga_ASC_detailed_le2001_wd_100m] and [raster_travel_time_r_NER_ga_ASC_detailed_le2001_walkvehd_100m]

2002: Input [v_NER_ASC_detailed_ASC_le2002]; output travel time rasters [raster_travel_time_r_NER_ga_ASC_detailed_le2002_wd_100m] and [raster_travel_time_r_NER_ga_ASC_detailed_le2002_walkvehd_100m]

2003: Input [v_NER_ASC_detailed_ASC_le2003]; output travel time rasters [raster_travel_time_r_NER_ga_ASC_detailed_le2003_wd_100m] and [raster_travel_time_r_NER_ga_ASC_detailed_le2003_walkvehd_100m]

2004: Input [v_NER_ASC_detailed_ASC_le2004]; output travel time rasters [raster_travel_time_r_NER_ga_ASC_detailed_le2004_wd_100m] and [raster_travel_time_r_NER_ga_ASC_detailed_le2004_walkvehd_100m]

2005: Input [v_NER_ASC_detailed_ASC_le2005]; output travel time rasters [raster_travel_time_r_NER_ga_ASC_detailed_le2005_wd_100m] and [raster_travel_time_r_NER_ga_ASC_detailed_le2005_walkvehd_100m]

2006: Input [v_NER_ASC_detailed_ASC_le2006]; output travel time rasters [raster_travel_time_r_NER_ga_ASC_detailed_le2006_wd_100m] and [raster_travel_time_r_NER_ga_ASC_detailed_le2006_walkvehd_100m]

2007: Input [v_NER_ASC_detailed_ASC_le2007]; output travel time rasters [raster_travel_time_r_NER_ga_ASC_detailed_le2007_wd_100m] and [raster_travel_time_r_NER_ga_ASC_detailed_le2007_walkvehd_100m]

2008: Input [v_NER_ASC_detailed_ASC_le2008]; output travel time rasters [raster_travel_time_r_NER_ga_ASC_detailed_le2008_wd_100m] and [raster_travel_time_r_NER_ga_ASC_detailed_le2008_walkvehd_100m]

2009: Input [v_NER_ASC_detailed_ASC_le2009]; output travel time rasters [raster_travel_time_r_NER_ga_ASC_detailed_le2009_wd_100m] and [raster_travel_time_r_NER_ga_ASC_detailed_le2009_walkvehd_100m]

2010: Input [v_NER_ASC_detailed_ASC_le2010]; output travel time rasters [raster_travel_time_r_NER_ga_ASC_detailed_le2010_wd_100m] and [raster_travel_time_r_NER_ga_ASC_detailed_le2010_walkvehd_100m]

2011: Input [v_NER_ASC_detailed_ASC_le2011]; output travel time rasters [raster_travel_time_r_NER_ga_ASC_detailed_le2011_wd_100m] and [raster_travel_time_r_NER_ga_ASC_detailed_le2011_walkvehd_100m]

2012: Input [v_NER_ASC_detailed_ASC_le2012]; output travel time rasters [raster_travel_time_r_NER_ga_ASC_detailed_le2012_wd_100m] and [raster_travel_time_r_NER_ga_ASC_detailed_le2012_walkvehd_100m]

2013: Input [v_NER_ASC_detailed_ASC_le2013]; output travel time rasters [raster_travel_time_r_NER_ga_ASC_detailed_le2013_wd_100m] and [raster_travel_time_r_NER_ga_ASC_detailed_le2013_walkvehd_100m]

Female: Input [v_NER_ASC_detailed_ASC_female_100m]; output travel time rasters [raster_travel_time_r_NER_ga_ASC_detailed_female_wd_100m] and [raster_travel_time_r_NER_ga_ASC_detailed_female_walkvehd_100m]

Male: Input [v_NER_ASC_detailed_ASC_male_100m]; output travel time rasters [raster_travel_time_r_NER_ga_ASC_detailed_male_wd_100m] and [raster_travel_time_r_NER_ga_ASC_detailed_male_walkvehd_100m]

iCCM: Input [v_NER_ASC_detailed_iCCM_le2013_100m]; output travel time rasters [raster_travel_time_r_NER_ga_ASC_detailed_iCCM_le2013_wd_100m] and [raster_travel_time_r_NER_ga_ASC_detailed_iCCM_le2013_walkvehd_100m]

See Supplementary Appendix 1b at https://doi.org/10.5281/zenodo.4482969). See Table 1 for zonal statistics from these travel time rasters and detailed results for administrative layers 0-3 in Supplementary Appendix 2.

We calculated the additional contribution of the ASC network to geographic accessibility beyond the existing network of CSI and CS (without ASC) in 2013 at 60 minutes walking in dry conditions using the following steps:

1. We used the “Raster calculator” tool in QGIS 3.12.0-Bucareşti^13^ to prepare a raster [r_NER_ga_dummy_CS_without_ASC_wd_100m ] for the cells within 60 minutes walking in dry conditions of a CS in 2013, using the travel time raster [raster_travel_time_r_NER_ga_CS_wd_100m], but beyond 60 minutes walking in dry conditions of an ASC in 2013, using the travel time raster [raster_travel_time_r_NER_ga_ASC_detailed_le2013_wd_100m] at 100m resolution in CRS EPSG:32632 – WGS 84 / UTM zone 32N with the final DEM at 100m as the extent.
2. We used the “Raster calculator” tool in QGIS 3.12.0-Bucareşti^13^ to prepare a raster [r_NER_ga_dummy_additional_contribition_ASC_le2013_wd_100m] for the cells beyond 60 minutes of a CS (without an ASC), using travel time raster [r_NER_ga_dummy_CS_without_ASC_wd_100m], and beyond 60 minutes of a CSI, using travel time raster [raster_travel_time_r_NER_ga_CSI_wd_100m] but within 60 minutes of an ASC, using travel time raster [raster_travel_time_r_NER_ga_ASC_detailed_le2013_wd_100m] walking in dry conditions 2013 at 100m resolution in CRS EPSG:32632 – WGS 84 / UTM zone 32N with the final DEM at 100m as the extent.
3. We used the “Raster calculator” tool in QGIS 3.12.0-Bucareşti^13^ to multiply the dummy raster for the additional contribution of ASC in 2013 to geographic accessibility to basic health services beyond the existing CS (without ASC) and CSI networks [r_NER_ga_dummy_additional_contribition_ASC_le2013_wd_100m] by the travel time raster for geographic accessibility to an ASC in 2013 [raster_travel_time_r_NER_ga_ASC_detailed_le2013_wd_100m]. This resulted in a travel time raster for the areas with additional geographic accessibility beyond 60 minutes walking in dry conditions of a CSI or CS (without an ASC) due to the contribution of ASC in 2013 at 100m resolution in CRS EPSG:32632 – WGS 84 / UTM zone 32N with the final DEM at 100m as the extent [r_NER_ga_additional_contribition_ASC_le2013_wd_100m] (see Supplementary Appendix 1b at https://doi.org/10.5281/zenodo.4482969). See Table 1 for zonal statistics from these travel time rasters and detailed results for administrative layers 0-3 in Supplementary Appendix 2.

We repeated steps 1-4 above for the travel scenario walking to the nearest road and then taking motorized transportation, resulting in the a travel time raster for the areas with additional geographic accessibility beyond 60 minutes walking + motorized transportation in dry conditions of a CSI or CS (without an ASC) due to the contribution of ASC in 2013 at 100m resolution in CRS EPSG:32632 – WGS 84 / UTM zone 32N with the final DEM at 100m as the extent [r_NER_ga_additional_contribition_ASC_le2013_walkvehd_100m] (see Supplementary Appendix 1b at https://doi.org/10.5281/zenodo.4482969). See Table 1 for zonal statistics from these travel time rasters and detailed results for administrative layers 0-3 in Supplementary Appendix 2.

We repeated steps 1-4 above for ASC trained on iCCM, resulting in the a travel time raster for the areas with additional geographic accessibility to iCCM services beyond 60 minutes walking in dry conditions of a CSI or CS (without an ASC) due to the contribution of ASC trained on iCCM in 2013 at 100m resolution in CRS EPSG:32632 – WGS 84 / UTM zone 32N with the final DEM at 100m as the extent [r_NER_ga_additional_contribition_ASC_le2013_iCCM_wd_100m] (see Supplementary Appendix 1b at https://doi.org/10.5281/zenodo.4482969). See Table 1 for zonal statistics from these travel time rasters and detailed results for administrative layers 0-3 in Supplementary Appendix 2.

Finally we repeated steps 1-4 above for ASC trained on iCCM, using the walking + motorized transportation travel scenario, resulting in the a travel time raster for the areas with additional geographic accessibility to iCCM services beyond 60 minutes walking + motorized transportation in dry conditions of a CSI or CS (without an ASC) due to the contribution of ASC trained on iCCM in 2013 at 100m resolution in CRS EPSG:32632 – WGS 84 / UTM zone 32N with the final DEM at 100m as the extent [r_NER_ga_additional_contribition_ASC_le2013_iCCM_walkvehd_100m] (see Supplementary Appendix 1b at https://doi.org/10.5281/zenodo.4482969). See Table 1 for zonal statistics from these travel time rasters and detailed results for administrative layers 0-3 in Supplementary Appendix 2.

We used the “Zonal statistics” tool in Accessmod v5 to calculate the percent of the population beyond 60 min of a CSI and CS (without an ASC) that were within 30 minutes and 60 minutes of an ASC in 2013, using walking and walking + motorized transportation travel scenarios. See Table 1 for zonal statistics from these travel time rasters and detailed results for administrative layers 0-3 in Supplementary Appendix 2.

**Methods for Geographic Accessibility question 4**

We repeated steps 1-3 from research question 1, using the following facility inputs:

1. CS-ASC network
2. CS-ASC network without a severe stockout of any iCCM commodity (severe stockout=stockout of any iCCM commodity lasting longer than seven days; iCCM commodities = RDT and AL for malaria, low osmolarity ORS and zinc sulfate for diarrhea, cotrimoxazole (pill or syrup) for pneumonia. A stockout of any of these commodities lasting longer than 7 days resulted in the CS being considered as a CS with a severe stockout of any iCCM commodity.)
3. CS-ASC network with trained human resources (nurse and/or ASC)
4. CS-ASC network with trained human resources (nurse and/or ASC) and no stockout of any iCCM commodity (severe stockout=stockout of any iCCM commodity lasting longer than seven days; iCCM commodities = RDT and AL for malaria, low osmolarity ORS and zinc sulfate for diarrhea, cotrimoxazole (pill or syrup) for pneumonia. A stockout of any of these commodities lasting longer than 7 days resulted in the CS being considered as a CS with a severe stockout of any iCCM commodity.)
5. CS-ASC network with trained human resources (nurse and/or ASC) and no severe stockout of RDT or AL (severe stockout=stockout of any iCCM commodity lasting longer than seven days; iCCM commodities = RDT and AL for malaria)
6. CS-ASC network with trained human resources (nurse and/or ASC) and no severe stockout of ORS or zinc (severe stockout=stockout of any iCCM commodity lasting longer than seven days; ORS = low osmolarity oral rehydration solution)
7. CS-ASC network with trained human resources (nurse and/or ASC) and no severe stockout of cotromoxazole (pill or syrup) (severe stockout=stockout of any iCCM commodity lasting longer than seven days; cotrimoxazole was the first-line antibiotic for pneumonia)
8. CS-ASC network with trained human resources (nurse and/or ASC) and no severe stockout of RUTF (severe stockout=stockout of any iCCM commodity lasting longer than seven days; RUTF=ready-to-eat therapeutic food)

See Supplementary Appendix 1b at https://doi.org/10.5281/zenodo.4482969). See Table 1 for zonal statistics from these travel time rasters and detailed results for administrative layers 0-3 in Supplementary Appendix 2.

**Geographic coverage**

We defined geographic coverage as the theoretical catchment area of a health service delivery location, within a maximum travel time, accounting for the mode of transportation and the maximum population capacity of the type of health service delivery location.^10^ We used the "geographic coverage" module of AccessMod 5 (v5.6.48)^10^ to estimate geographic coverage for the CSI and CS-ASC networks in 2013 at 1km x 1km resolution for the two travel scenarios. The maximum travel time was set at 60 minutes. The maximum population capacity was set at 10000 for CSI and 2500 for CS-ASC based on the norms of the MOPH of Niger.^15^ The maximum extent of a catchment was therefore delimited by the maximum travel time of 60 minutes except in cases where the estimated population in the catchment exceeded the maximum population capacity of the health service delivery location – in which case the extent of the catchment was smaller than the maximum travel time and was defined by the area containing the estimated population, up to the maximum population capacity.

Research questions

1. What percentage of the estimated population was covered by the CSI network in 2013?
2. What percentage of the estimated residual population beyond the geographic coverage of the existing CSI network was covered by the CS-ASC network in 2013?
3. What percentage of the estimated population was covered by the combination of the CSI and CS-ASC networks in 2013?

**Methods for Geographic Coverage research question 1**

We conducted a geographic coverage analysis of the estimated population covered by the existing CSI network in 2013, with each CSI catchment defined by a maximum travel time of 60 min (walking or walking + motorized vehicle) and maximum population capacity of 10000.

Data preparation

1. Preparation of the processing order variable “rFB13TT” within the point vector file for the existing CSI network: We conducted a geographic coverage analysis for the CSI network in 2013 without considering maximum population capacity. This provided the shapefile defining the 60 min catchment (not considering maximum population capacity) for each CSI [r_NER_gcCSI _60min_1km_no_capacity].
   a. We used the following data inputs:
      i. Population: raster_population_r_NER_FBpop2013_1km_final
      ii. Land cover merged: raster_land_cover_merged_r_NER_land_merged
      iii. Scenario table: table_scenario_walk_dry
      iv. Select existing health facilities layer (vector): v_NER_CSI_adj_barriers_1km
      v. ID field: id
      vi. Facility name field: nom_centre
      vii. Select zones layer (vector): adm3
           1. Select zones unique ID (integer): objectid
           2. Select zone name (text): nom_com
   b. We used the following analysis settings:
      i. Type of analysis: anisotropic
      ii. Direction of travel: towards facilities
      iii. Facilities processing order according to: A field in the facility layer “capacity”
      iv. Processing order: Descending
      v. Maximum travel time (minutes): 60
      vi. Options
          1. Compute population catchment area layer: Yes
          2. Remove the covered population at each iteration: Yes
          3. Compute a layer of population cells on barriers: Yes
          4. Generate zonal statistics: Yes (adm 3)
          5. Run the analysis without considering capacities: Yes
          6. Add column with original population sum under each facility’s travel time: Yes
          7. Optimize dynamically computation according to the scenario: Yes
          8. Add short tag: r_NER_gcCSI _60min_1km_no_capacity
   c. We calculated the population in 2013 for each catchment area from Data Processing step 1 (variable called “FB13TT”, using zonal statistics based on the population file “r_NER_FB_pop_2013_1km_final” and the shapefile generated from Data Processing step 1.
   d. We used a spatial join to link the shapefile from step 1a (with the variable “FB13TT”) to the input vector file v_NER_CSI_adj_barriers_1km”, using the unique ID for each CSI to facilitate the join.

Analysis

1. Geographic coverage analysis of the existing CSI network in 2013: We completed a geographic coverage analysis for the CSI network in 2013 considering maximum population capacity using the variable “capacity” and processing order based on the population of each catchment using the variable “FB13TT” derived from Data Processing step 2. This provided the final outputs for the CSI geographic coverage analysis.
a. We used the following data inputs:
   i. Population: raster_population_r_NER_FBpop2013_1km_final
   ii. Land cover merged: raster_land_cover_merged_r_NER_land_merged
   iii. Scenario table: table_scenario_walk_dry
   iv. Select existing health facilities layer (vector): v_NER_CSI_adj_barriers_1km
   v. ID field: id
   vi. Facility name field: nom_centre
   vii. Capacity: capacity
   viii. Select zones layer (vector): adm3
         1. Select zones unique ID (integer): objectid
         2. Select zone name (text): nom_com
b. We used the following analysis settings:
   ix. Type of analysis: anisotropic
   x. Direction of travel: towards facilities
   xi. Facilities processing order according to: A field in the facility layer “FB13TT”
   xii. Processing order: Descending
   xiii. Maximum travel time (minutes): 60
   xiv. Options
        1. Compute population catchment area layer: Yes
        2. Remove the covered population at each iteration: Yes
        3. Compute a layer of population cells on barriers: Yes
        4. Generate zonal statistics: Yes (adm 3)
        5. Run the analysis without considering capacities: No
        6. Add column with original population sum under each facility’s travel time: Yes
        7. Optimize dynamically computation according to the scenario: Yes
        8. Add short tag: r_NER_gcCSI _60min_1km_prioritize_FB13TT

Variable “amPopCoveredPercent” in the tab “Pop_CSI” of Supplementary Appendix 3 provides the cumulative geographic coverage of the population covered by the CSI network. See Supplementary Appendix 1b at https://doi.org/10.5281/zenodo.4482969) for the vector shapefile (polygons) of the modelled catchment area of each CSI.

**Methods for Geographic Coverage research question 2**

We conducted a geographic coverage analysis of the estimated residual population beyond the geographic coverage of the existing CSI network in 2013 that were covered by the existing CS-ASC network in 2013, with each CS-ASC catchment defined by a maximum travel time of 60 min (walking or walking + motorized vehicle) and maximum population capacity of 2500.

Data preparation

1. Preparation of the processing order variable “rFB13TT” within the point vector file for the existing CS-ASC network in 2013: We conducted a geographic coverage analysis for the CS-ASC network in 2013 without considering maximum population capacity. This provided the shapefile defining the 60 min catchment (not considering maximum population capacity) for each CS-ASC [r_NER_gcCS-ASC_60min_1km_no_capacity].
   a. We used the following data inputs:
      i. Population: raster_population_residual_r_NER_gcCSI_60min_1km_prioritize_FB13TT
      ii. Land cover merged: raster_land_cover_merged_r_NER_land_merged
      iii. Scenario table: table_scenario_walk_dry
      iv. Select existing health facilities layer (vector): v_NER_cells_at_100m_with_CS_or_ASC_adj_barriers_1km
      v. ID field: id
      vi. Facility name field: cat
      vii. Select zones layer (vector): adm3
           1. Select zones unique ID (integer): objectid
           2. Select zone name (text): nom_com
   b. We used the following analysis settings:
      i. Type of analysis: anisotropic
      ii. Direction of travel: towards facilities
      iii. Facilities processing order according to: A field in the facility layer “capacity”
      iv. Processing order: Descending
      v. Maximum travel time (minutes): 60
      vi. Options
          1. Compute population catchment area layer: Yes
          2. Remove the covered population at each iteration: Yes
          3. Compute a layer of population cells on barriers: Yes
          4. Generate zonal statistics: Yes (adm 3)
          5. Run the analysis without considering capacities: Yes
          6. Add column with original population sum under each facility’s travel time: Yes
          7. Optimize dynamically computation according to the scenario: Yes
          8. Add short tag: r_NER_gcCS-ASC_60min_1km_no_capacity
   c. We used zonal statistics to calculate the population in 2013 for each catchment area in Data Preparation step 1 in a variable called “rFB13TT”.
   d. We used a spatial join to link the shapefile from Data Preparation step 1 (with the variable “rFB13TT”) to the input vector file v_NER_CSI_adj_barriers_1km”, using the unique ID for each CS-ASC to facilitate the join.

Data analysis

1. We conducted a geographic coverage analysis for the CS-ASC network in 2013 considering maximum population capacity using the variable “capacity” (set at 2500 population per MOH norms) and processing order based on the population of each catchment using the variable “rFB13TT”. This provided the final outputs for the CS-ASC geographic coverage analysis.
   a. We used the following data inputs:
      i. Population: raster_population_residual_r_NER_gcCSI_60min_1km_prioritize_FB13TT
      ii. Land cover merged: raster_land_cover_merged_r_NER_land_merged
      iii. Scenario table: table_scenario_walk_dry
      iv. Select existing health facilities layer (vector): v_NER_cells_at_100m_with_CS_or_ASC_adj_barriers_1km
      v. ID field: id
      vi. Facility name field: cat
      vii. Capacity: capacity
      viii. Select zones layer (vector): adm3
            1. Select zones unique ID (integer): objectid
            2. Select zone name (text): nom_com
   b. We used the following analysis settings:
      i. Type of analysis: anisotropic
      ii. Direction of travel: towards facilities
      iii. Facilities processing order according to: A field in the facility layer “rFB13TT”
      iv. Processing order: Descending
      v. Maximum travel time (minutes): 60
      vi. Options
          1. Compute population catchment area layer: Yes
          2. Remove the covered population at each iteration: Yes
          3. Compute a layer of population cells on barriers: Yes
          4. Generate zonal statistics: Yes (adm 3)
          5. Run the analysis without considering capacities: No
          6. Add column with original population sum under each facility’s travel time: Yes
          7. Optimize dynamically computation according to the scenario: Yes
          8. Add short tag: r_NER_gcCS-ASC _60min_1km_prioritize_rFB13TT

Variable “amPopCoveredPercent” in the tab “Pop_CS-ASC” of Supplementary Appendix 3 provides the cumulative geographic coverage of the population covered by the CS-ASC network. See Supplementary Appendix 1b at https://doi.org/10.5281/zenodo.4482969 for the vector shapefile (polygons) of the modelled catchment area of each CS-ASC.

**Methods for Geographic Coverage research question 3**

The zonal statistics from Geographic Coverage research question 2 defacto provide the geographic coverage of the combined CSI + CS-ASC network.

**Scaleup**

Research questions

1. How many community health workers are needed (and where) to optimally cover the population beyond the 1-hour catchment of the existing network of CS + ASC and CSI?

The MoPH in Niger has planned to scale-up RC in communities beyond 5km of CS or CSI to provide a standard package of preventive, promotive and curative services, including iCCM. We conducted a geographic coverage analysis to determine how many RC would be needed (and where) to optimally cover the estimated residual population beyond the geographic coverage of the existing CSI and CS-ASC networks in 2013, within a maximum travel time of 60 min walking from/to the RC and maximum population capacity of 1000 for each RC. This analysis aimed to provide information (or at least a methodology) that could be used to inform a rational scale-up of the RC that would maximize geographic coverage of the residual population beyond the geographic coverage of the CS-ASC and CSI networks in 2013.

**Methods for Scaleup research question 1**

Data preparation

1. Identification of potential RC sites for scaleup:
   a. Given the norm for the RC-to-population ratio is 1 per 1000, we used a 500 people as a minimum cutoff to identify cells for potential RC sites because it would be inefficient and impractical to place RC in all communities beyond the geographic coverage of the existing CS-ASC and CSI networks, regardless of population size. We used the “Raster calculator” tool in QGIS 3.12.0 to prepare a GeoTiff raster that identified cells from the residual population raster of the geographic coverage analysis of the existing CS-ASC network in 2013 [raster_population_residual_r_NER_gcCS_ASC_60min_1km_prioritize_rFB13TT] with greater than or equal to 500 people. Note that the cells identified here were also beyond the geographic coverage of the CSI network, since the geographic coverage analysis for the existing CS-ASC network used the residual population from the geographic coverage analysis of the existing CSI network as the input population dataset. This resulted in 3521 cells identified as potential RC sites for scaleup.
   b. We used the “Polygonize” tool in QGIS 3.12.0-Bucareşti^13^ to convert the Geotiff raster from step 1 to a vector shapefile of 3521 potential RC sites for scaleup [v_NER_scaleup_RC_rFB13TTge500_1km].
2. Preparation of the processing order variable “rFB13TT” within the point vector file for the potential RC network:
a. We conducted a geographic coverage analysis of the estimated residual population beyond the geographic coverage of the existing CS-ASC network in 2013 (and defacto beyond the geographic coverage of the combined CSI + CS-ASC network) that were covered by a hypothetical network of RC in 2013, with each RC catchment defined by a maximum travel time of 60 min (walking or walking + motorized vehicle) without considering maximum population capacity [r_NER_gcscaleup_potential_sites_ge500_60min_wd_1km_no_capacity.zip]. This provided the shapefile defining the 60 min catchment (not considering maximum population capacity) for each potential RC site:
   i. We used the following data inputs:
      1. Population: raster_population_residual_r_NER_gcCS_ASC_60min_1km_prioritize_rFB13TT
      2. Land cover merged: raster_land_cover_merged_r_NER_land_merged
      3. Scenario table: table_scenario_walk_dry
      4. Select existing health facilities layer (vector): v_NER_scaleup_RC_rFB13TTge500_1km
      5. ID field: id
      6. Facility name field: cat
      7. Select zones layer (vector): adm3
         a. Select zones unique ID (integer): objectid
         b. Select zone name (text): nom_com
   ii. We used the following analysis settings:
       1. Type of analysis: anisotropic
       2. Direction of travel: towards facilities
       3. Facilities processing order according to: A field in the facility layer “capacity”
       4. Processing order: Descending
       5. Maximum travel time (minutes): 60
       6. Options
          a. Compute population catchment area layer: Yes
          b. Remove the covered population at each iteration: Yes
          c. Compute a layer of population cells on barriers: Yes
          d. Generate zonal statistics: Yes (adm 3)
          e. Run the analysis without considering capacities: Yes
          f. Add column with original population sum under each facility’s travel time: Yes
          g. Optimize dynamically computation according to the scenario: Yes
          h. Add short tag: r_NER_gcscaleup_RC_rFB13TTge500_60min_wd_1km_no_capacity
b. We used zonal statistics to calculate the population in 2013 for each catchment area in Data Preparation step 2 in a variable called “rFB13TT”.
c. We used a spatial join to link the shapefile from Data Preparation step 2 (with the variable “rFB13TT”) to the input vector file [v_NER_scaleup_potential_sites_ge500_1km] using the unique ID for each hypothetical RC site to facilitate the join.
3. Identification of the number of RC per hypothetical RC site and total capacity per hypothetical RC site:
   a. We created a new whole number integer variable “RC” in the vector shapefile from step 2 that divided the residual population “rFB13TT” in each potential RC site by 1000 (the MOPH norm) resulting in the number of RC per potential RC site, with a cutoff of 500 people used to round to the nearest 1 RC. Use of a whole number reflects the reality that policy makers cannot deploy fractions of RC, with the implication that some communities may be slightly “over-supplied” or “under-supplied” (e.g. a community of 500 people would receive 1 RC and, with a maximum population capacity of 1000 people, there would be a surplus capacity of 500 people which could be used to serve nearby satellite communities; a community of 1200 people would receive 1 RC and, with a maximum population capacity of 1000, there would be a capacity deficit of 200 people which would need to be addressed by nearby RC with surplus capacity or additional RC.)
   b. Calculation of the total capacity of each potential RC site: We created a new whole number integer variable “capacity” in the vector shapefile from step 2 that multiplied the number of RC in the variable “RC” by the maximum population capacity per RC of 1000. This provided the total capacity of each of the 3521 potential RC sites.
   c. We converted the vector shapefile from step 3 to a vector point file of the 3521 potential RC sites for use in our analysis of geographic coverage of the hypothetical “optimized” RC network [v_NER_scaleup_RC_rFB13TTge500_1km].

Analysis

1. We conducted a geographic coverage analysis of the estimated residual population beyond the geographic coverage of the existing CS-ASC network in 2013 (and defacto beyond the geographic coverage of the combined CSI + CS-ASC network) that were covered by a hypothetical network of RC in 2013, with each RC site catchment defined by a maximum travel time of 60 min (walking or walking + motorized vehicle) and total maximum population capacity dependent on the number of RC x 1000 people, and processing order prioritizing (highest to lowest) the residual population “rFB13TT”.
   a. We used the following data inputs:
i. Population: raster_population_residual_r_NER_gcCS_ASC_60min_1km_prioritize_FB13TT
ii. Land cover merged: raster_land_cover_merged_r_NER_land_merged
iii. Scenario table: table_scenario_walk_dry
iv. Select existing health facilities layer (vector): v_NER_scaleup_RC_rFB13TTge500_1km
v. ID field: id
vi. Facility name field: cat
vii. Capacity: capacity
viii. Select zones layer (vector): adm3
1. Select zones unique ID (integer): objectid
2. Select zone name (text): nom_com
   b. We used the following analysis settings:
i. Type of analysis: anisotropic
ii. Direction of travel: towards facilities
iii. Facilities processing order according to: A field in the facility layer “rFB13TT”
iv. Processing order: Descending
v. Maximum travel time (minutes): 60
vi. Options
1. Compute population catchment area layer: Yes
2. Remove the covered population at each iteration: Yes
3. Compute a layer of population cells on barriers: Yes
4. Generate zonal statistics: Yes (adm 3)
5. Run the analysis without considering capacities: No
6. Add column with original population sum under each facility’s travel time: Yes
7. Optimize dynamically computation according to the scenario: Yes
8. Add short tag: r_NER_gcscaleup_RC_rFB13TTge500_60min_wd_1km_prioritize_rFB13TT

See Supplementary Appendix 4 for outputs of the scale-up analysis and Supplementary Appendix 1b at https://doi.org/10.5281/zenodo.4482969 for the vector shapefiles (polygons) of the modelled catchment areas of each RC in the scaled-up RC network.

**Targeting**

We assessed how well targeted the existing network of CS-ASC in 2013 was in terms of targeting a) the estimated residual population b) the estimated residual under-five deaths and c) the estimated residual *Pf* malaria cases beyond the catchment of the CSI network in 2013 compared to three hypothetical CS-ASC networks:

a. Hypothetical CS-ASC network that optimized geographic coverage of the estimated residual population beyond the catchment of the existing CSI network in 2013 by ordering the deployment (processing order) based on the estimated residual population in 2013 within the catchment area of a given CS-ASC, prioritizing catchments with higher estimated residual population over those with lower estimated residual population
b. Hypothetical CS-ASC network that optimized geographic coverage of the estimated residual under-five deaths beyond the catchment of the existing CSI network in 2013 by ordering the deployment (processing order) based on the estimated residual under-five deaths in 2013 within the catchment area of a given CS-ASC, prioritizing catchments with higher estimated residual under-five deaths over those with lower estimated residual under-five deaths
c. Hypothetical CS-ASC network that optimized geographic coverage of the estimated residual *Pf* malaria cases among all ages (0-99 years) beyond the catchment of the existing CSI network in 2013 by ordering the deployment (processing order) based on the estimated residual *Pf* malaria cases among all ages (0-99 years) in 2013 within the catchment area of a given CS-ASC, prioritizing catchments with higher estimated residual *Pf* malaria cases over those with lower estimated residual *Pf* malaria cases.

Research questions

1. How well targeted was the existing network of CS-ASC in 2013 in terms of geographic coverage of the estimated residual population beyond the catchment of the existing CSI network in 2013 compared to a hypothetical network of CS-ASC deployed to optimize geographic coverage of the residual estimated population?
2. How well targeted was the existing network of CS-ASC in 2013 in terms of geographic coverage of the estimated residual under-five deaths beyond the catchment of the existing CSI network compared to a hypothetical network of CS-ASC deployed to optimize geographic coverage of the estimated residual under-five deaths?
3. How well targeted was the existing network of CS-ASC in 2013 in terms of geographic coverage of the estimated residual *Pf* malaria cases among all ages (0-99 years) beyond the catchment of the existing CSI network compared to a hypothetical network of CS-ASC deployed to optimize geographic coverage of the estimated residual *Pf* malaria cases?

**Methods for Targeting research question 1**

Data preparation

1. Preparation of the GeoTiff for the estimated count of the residual population beyond the geographic coverage of the existing CSI network in 2013 [raster_population_residual_ r_NER_gcCS-ASC _60min_1km_prioritize_rFB13TT]:
   a. See Methods for Geographic Coverage research question 2, Data analysis, step 1
2. Preparation of the processing order variable “rFB13TT” within the point vector file for the existing CS-ASC network:
   a. See Methods for Geographic Coverage research question 2, Data preparation, step 1.
3. Preparation of the processing order variable “rFB13TT” within the point vector file for the hypothetical CS-ASC network: We conducted a geographic coverage analysis for the network of 5796 potential CS-ASC sites in 2013 without considering maximum population capacity. This provided the shapefile defining the 60 min catchment (not considering maximum population capacity) for each hypothetical CS-ASC site. File “r_NER_gcCS-ASC_60min_1km_no_capacity”
   a. We used the following data inputs:
      i. Population: raster_population_residual_r_NER_gcCSI_60min_1km_prioritize_FB13TT
      ii. Land cover merged: raster_land_cover_merged_r_NER_land_merged
      iii. Scenario table: table_scenario_walk_dry
      iv. Select existing health facilities layer (vector): v_NER_targeting_potentialCS-ASC_sites_FB13TTge500_1km
      v. ID field: id
      vi. Facility name field: cat
      vii. Select zones layer (vector): adm3
           1. Select zones unique ID (integer): objectid
           2. Select zone name (text): nom_com
   b. We used the following analysis settings:
      i. Type of analysis: anisotropic
      ii. Direction of travel: towards facilities
      iii. Facilities processing order according to: A field in the facility layer “capacity”
      iv. Processing order: Descending
      v. Maximum travel time (minutes): 60
      vi. Options
          1. Compute population catchment area layer: Yes
          2. Remove the covered population at each iteration: Yes
          3. Compute a layer of population cells on barriers: Yes
          4. Generate zonal statistics: Yes (adm 3)
          5. Run the analysis without considering capacities: Yes
          6. Add column with original population sum under each facility’s travel time: Yes
          7. Optimize dynamically computation according to the scenario: Yes
          8. Add short tag: r_NER_potentialCS_ASC_FB13TTge500_60min_1km_no_capacity
   c. We used zonal statistics to calculate the residual population in 2013 for each catchment area, using the residual population from the geographic coverage analysis of the CSI network [raster_population_residual_r_NER_gcCSI_60min_1km_prioritize_FB13TT] and the shapefile for the geographic coverage analysis of the hypothetical CS-ASC network in Data Preparation step 3a above [shape_catchment_r_NER_gcCS_ASC_60min_1km_no_capacity] in a variable called “rFB13TT”.
   d. We used a spatial join to join the variable “rFB13TT” from the shapefile in Data Preparation step 2 to the input vector file [v_NER_Targeting_Hypothetical_CS_ASC_sites_FB13TTge500_1km] using the unique ID for each potential CS-ASC to facilitate the join.

Data analysis

1. Geographic coverage analysis of the estimated residual population by the existing network of CS-ASC, prioritizing estimated residual population: See Methods for Geographic Coverage research question 2, Data analysis, step 1.
2. Geographic coverage analysis of the estimated residual population by the hypothetical network of CS-ASC, prioritizing estimated residual population: We conducted a geographic coverage analysis for the network of the 5796 potential CS-ASC sites in 2013 considering maximum population capacity (set at 2500 population per MOPH norms) and a descending processing order (highest to lowest) based on the residual estimated population beyond the CSI network in 2013 within each catchment, using the variable “rFB13TT”. This prioritized the deployment of CS-ASC according to the size (highest to lowest) of the estimated residual population in their catchment. This provided the final outputs for the geographic coverage analysis for the hypothetical network of CS-ASC sites that prioritized geographic coverage of the estimated residual population.
   a. We used the following data inputs:
      i. Population: raster_population_residual_r_NER_gcCSI_60min_1km_prioritize_rFB13TT
      ii. Land cover merged: raster_land_cover_merged_r_NER_land_merged
      iii. Scenario table: table_scenario_walk_dry
      iv. Select existing health facilities layer (vector): v_NER_Targeting_Hypothetical_CS_ASC_sites_FB13TTge500_1km
      v. ID field: id
      vi. Facility name field: cat
      vii. Capacity: capacity
      viii. Select zones layer (vector): adm3
            1. Select zones unique ID (integer): objectid
            2. Select zone name (text): nom_com
   b. We used the following analysis settings:
      i. Type of analysis: anisotropic
      ii. Direction of travel: towards facilities
      iii. Facilities processing order according to: A field in the facility layer “rFB13TT”
      iv. Processing order: Descending
      v. Maximum travel time (minutes): 60
      vi. Options
          1. Compute population catchment area layer: Yes
          2. Remove the covered population at each iteration: Yes
          3. Compute a layer of population cells on barriers: Yes
          4. Generate zonal statistics: Yes (adm 3)
          5. Run the analysis without considering capacities: No
          6. Add column with original population sum under each facility’s travel time: Yes
          7. Optimize dynamically computation according to the scenario: Yes
          8. Add short tag: r_NER_Targeting_Hypothetical_CS_ASC_FB13TTge500_60min_1km_prioritize_rFB13TT

For outputs, see Supplementary Appendix 5, tabs “Pop_Existing” and “Pop_Hypothetical”, in which the variable “amPopCoveredPercent” indicates the cumulative geographic coverage of the residual population. Supplementary Appendix 1b at https://doi.org/10.5281/zenodo.4482969 contains the vector shapefile (polygon) indicating the modelled catchment area of each health service delivery point.

3. Comparison of geographic coverage of the existing CS-ASC network and the hypothetical CS-ASC network: See tabs “Pop_Existing”, “Pop_Hypothetical” and “Comparison_Population” in Appendix 6. We compared the percentage of the estimated residual population beyond the geographic coverage of the existing CSI network in 2013 that was covered by the existing network of CS-ASC (from Geographic Coverage research Question 2) with the percentage of the estimated residual population beyond the geographic coverage of the existing CSI network in 2013 that was covered by the hypothetical network of CS-ASC that prioritized the estimated residual population in the processing order (from Targeting research question 1, Data Analysis step 1 above) given the same number of potential CS-ASC sites as in the existing network of CS-ASC (i.e. 2550) as well as for the total number of potential CS-ASC sites (i.e. 5796).
   a. In tab “Pop_Existing” of Supplementary Appendix 5, we ensured the results were sorted using variable “amPopOrigTravelTimeMax_rFB13TT” from highest to lowest (this was defacto the case, having set the processing order as descending based on variable “rFB13TT”). Maintaining this order, the last value for the variable “amPopCoveredPercent_rFB13TT” provided the percentage of the population beyond the 1hr catchment of the existing CSI network in 2013 that was covered by the existing network of CS-ASC.
   b. In tab “Pop_Hypothetical” of Supplementary Appendix 5, we ensured the results were sorted using variable “amPopOrigTravelTimeMax_rFB13TT” from highest to lowest (this was the default, having set the processing order as descending based on “rFB13TT”). Maintaining this order, the value for the variable “amPopCoveredPercent_rFB13TT” for the 2550^th^ potential CS-ASC provided the percentage of the population beyond the 1hr catchment of the existing CSI network in 2013 that was covered by the hypothetical network of CS-ASC that prioritized the residual population in the processing order, using the same number of sites as the existing CS-ASC network. The value for the variable “amPopCoveredPercent_rFB13TT” for the 5796^th^ potential CS-ASC provided the percentage of the population beyond the 1hr catchment of the existing CSI network in 2013 that was covered by the hypothetical network of CS-ASC, prioritizing the residual population in the processing order.

**Methods for Targeting research question 2**

Data preparation

1. Preparation of the GeoTiff for the estimated count of residual under-five deaths beyond the geographic coverage of the existing CSI network
   a. See section I. Data inputs, Estimated under-five mortality for details.
2. Preparation of the processing order variable “rFB13TT” within the point vector file for the existing CS-ASC network:
   a. See section Methods for Targeting research question 1, Data preparation, step 3.
3. Preparation of the processing order variable “rU5d13” within the point vector file for the hypothetical network of CS-ASC:
   a. Based on the geographic coverage analysis of the hypothetical CS-ASC network, not considering maximum population capacity (Methods for Targeting research question 2, Data preparation, step 1) we used zonal statistics to calculate the estimated count of the residual under-five deaths beyond the geographic coverage of the existing CSI network in 2013 for the catchment area of each hypothetical CS-ASC site, using the GeoTiff of the estimated residual under-five deaths in Data preparation step 1 [r_NER_residU5d13_gcCSI_60min_1km] and the shapefile for the geographic coverage analysis of the hypothetical CS-ASC network in Data Preparation step 2 above [shape_catchment_r_NER_gcCS_ASC_60min_1km_no capacity] in a variable called “rU5d13”.
   b. We used a spatial join to join the variable “rU5d13” in the shapefile from Data Preparation step 3a to the input point vector file [v_NER_Targeting_Hypothetical_CS_ASC_sites_FB13TTge500_1km] using the unique ID for each hypothetical CS-ASC to facilitate the join.

Analysis

1. Geographic coverage analysis of the estimated residual under-five deaths by the existing network of CS-ASC, prioritizing estimated residual population in the processing order: We conducted a geographic coverage analysis for the estimated residual under-five deaths beyond the geographic coverage of the existing CSI network, using the existing network of CS-ASC sties in 2013, with the processing order based on the estimated residual population “rFB13TT” within each catchment area and setting the maximum population capacity at 100000 to effectively not consider maximum population capacity as a constraint to the CS-ASC catchment areas. The analysis removed the under-five deaths within each catchment area at each iteration (calculation of each catchment area) to avoid double counting under-five deaths where the 60 min catchment areas overlap. This provided the final outputs for the analysis of geographic coverage of the estimated residual under-five deaths by the existing CS-ASC network.
   a. We used the following data inputs:
      i. Population: r_NER_residU5d13_gcCSI_60min_1km
      ii. Land cover merged: raster_land_cover_merged_r_NER_land_merged
      iii. Scenario table: table_scenario_walk_dry
      iv. Select existing health facilities layer (vector): v_NER_cells_at_100m_with_CS_or_ASC_adj_barriers_1km
      v. ID field: id
      vi. Facility name field: cat
      vii. Capacity: capacity
      viii. Select zones layer (vector): adm3
            1. Select zones unique ID (integer): objectid
            2. Select zone name (text): nom_com
   b. We used the following analysis settings:
      ix. Type of analysis: anisotropic
      x. Direction of travel: towards facilities
      xi. Facilities processing order according to: A field in the facility layer “rFB13TT”
      xii. Processing order: Descending
      xiii. Maximum travel time (minutes): 60
      xiv. Options
           1. Compute population catchment area layer: Yes
           2. Remove the covered population at each iteration: Yes
           3. Compute a layer of population cells on barriers: Yes
           4. Generate zonal statistics: Yes (adm 3)
           5. Run the analysis without considering capacities: No
           6. Add column with original population sum under each facility’s travel time: Yes
           7. Optimize dynamically computation according to the scenario: Yes
           8. Add short tag: r_NER_gcCS_ASC_rU5d13_60min_1km_prioritize_rFB13TT
2. Geographic coverage analysis of the estimated residual under-five deaths by the hypothetical network of CS-ASC, prioritizing estimated residual under-five deaths in the processing order: We conducted a geographic coverage analysis for the estimated residual under-five deaths beyond the geographic coverage of the existing CSI network, using the hypothetical network of CS-ASC sites in 2013, with the processing order based on the estimated residual count of under-five deaths within each catchment “rU5d13” and setting the maximum population capacity at 100000 to effectively not consider maximum population capacity as a constraint to the CS-ASC catchment areas. The analysis removed the under-five deaths within each catchment area at each iteration (calculation of each catchment area) to avoid double counting under-five deaths where the 60 min catchment areas overlap. This provided the final outputs for the geographic coverage analysis for the optimized CS-ASC network, prioritizing deployment based on the estimated count of under-five deaths.
   a. We used the following data inputs:
i. Population: r_NER_residU5d13_gcCSI_60min_1km
ii. Land cover merged: raster_land_cover_merged_r_NER_land_merged
iii. Scenario table: table_scenario_walk_dry
iv. Select existing health facilities layer (vector): v_NER_Targeting_Hypothetical_CS_ASC_sites_FB13TTge500_1km
v. ID field: id
vi. Facility name field: cat
vii. Capacity: capacity
viii. Select zones layer (vector): adm3
1. Select zones unique ID (integer): objectid
2. Select zone name (text): nom_com
   b. We used the following analysis settings:
ix. Type of analysis: anisotropic
x. Direction of travel: towards facilities
xi. Facilities processing order according to: A field in the facility layer “rU5d13”
xii. Processing order: Descending
xiii. Maximum travel time (minutes): 60
xiv. Options
1. Compute population catchment area layer: Yes
2. Remove the covered population at each iteration: Yes
3. Compute a layer of population cells on barriers: Yes
4. Generate zonal statistics: Yes (adm 3)
5. Run the analysis without considering capacities: No
6. Add column with original population sum under each facility’s travel time: Yes
7. Optimize dynamically computation according to the scenario: Yes
8. Add short tag: r_NER_gcTargeting_Hypothetical_CS_ASC_rU5d13_60min_1km_prioritize_ru5d13

For outputs, see Supplementary Appendix 5, tabs “U5d_Existing” and “U5d_Hypothetical”, in which the variable “amPopCoveredPercent” indicates the cumulative geographic coverage of the residual U5 deaths. Supplementary Appendix 1b at https://doi.org/10.5281/zenodo.4482969 contains the vector shapefile (polygon) indicating the modelled catchment area of each health service delivery point.

3. Comparison of geographic coverage of the existing CS-ASC network and the hypothetical CS-ASC network: See tabs “U5d_Existing”, “U5d_Hypothetical” and “Comparison_U5deaths” in Supplementary Appendix 5. We compared the percentage of the estimated residual U5 deaths beyond the geographic coverage of the existing CSI network in 2013 that was covered by the existing network of CS-ASC, prioritizing the estimated residual population in the processing order, with the percentage of the estimated residual U5 deaths beyond the geographic coverage of the existing CSI network in 2013 that was covered by the hypothetical network of CS-ASC, prioritizing the estimated residual U5 deaths in the processing order given the same number of potential CS-ASC sites as in the existing network of CS-ASC (i.e. 2550) as well as for the total number of potential CS-ASC sites (i.e. 5796).
   a. In tab “U5d_Existing” of Supplementary Appendix 5, the last value for the variable “amPercentCovered” provided the cumulative percent of estimated residual U5 deaths beyond the geographic coverage of the existing CSI network that were covered by the existing CS-ASC network in 2013.
   b. In the tab “U5d_Hypothetical” of Supplementary Appendix 5, the last value for the variable “amPercentCovered” provided the cumulative percent of the estimated residual U5 deaths beyond the geographic coverage of the existing CSI network that were covered by the hypothetical CS-ASC network in 2013, where the estimated residual under-five deaths, prioritizing the residual U5 deaths in the processing order.
   c. In tab “Comparison_U5deaths”, we compared the results from 3a and to the results from 3b for the first 2550 hypothetical CS-ASC sites in 2013 potential CS-ASC sites to ensure comparability with the existing CS-ASC network in 2013. We then compared the results from 3a with the results from the full network of 5796 hypothetical CS-ASC sites in 2013.

Uncertainty analysis

We assessed the potential effect of uncertainty of the estimates for under-five deaths on targeting as follows. We used the “Zonal statistics” tool in QGIS 3.12.0-Bucareşti^13^ to extract the estimated mean and 95% confidence intervals for the number of under-five deaths for each catchment area defined by the geographic coverage analysis for the hypothetical network from step 2 of targeting research question 2. We sorted the catchments by the estimated mean number of under-five deaths from largest to smallest, as this reflected the prioritization order of the geographic coverage analysis used for the targeting analysis (step 2 of targeting research question 2). Because policy makers and planners typically support scale-up of facilities and CHWs in groups or “blocks” we identified four potential “blocks” of CS-ASC for consideration. Block 1 included the 500 CS-ASC with the highest estimated mean number of under-five deaths, (median of means across catchments = 688, lower 95% confidence interval minimum = 97, and upper 95% confidence interval maximum = 7211). Block 2 included 1925 CS-ASC with the next highest estimated mean number of under-five deaths (median of means across catchments = 121, lower 95% confidence interval minimum = 30, and upper 95% confidence interval maximum = 329). Block 3 included 2992 CS-ASC with next highest estimated mean number of under-five deaths (median of means across catchments = 46, lower 95% confidence interval minimum = 20, and upper 95% confidence interval maximum = 95). Block 4 included 379 catchments with the next highest mean number of under-five deaths (median of means across catchments = 12, lower 95% confidence interval minimum = 0, and maximum 95% confidence interval maximum = 24). Based on the 95% confidence interval minimums and maximums, the estimated number of under-five deaths was significantly higher in Block 1 than Block 3 or Block 4, and the estimated number of under-five deaths was significantly higher in Block 2 than Block 4, suggesting that policy makers and planners could confidently prioritize Block 1 over Block 3 and Block 4, and prioritize Block 2 over Block 4 (see Supplementary Appendix 5 – Targeting uncertainty, tabs “Summary_uncertainty_rU5d13” and “Blocks_uncertainty_rU5d13”.

**Methods for Targeting research question 3**

Data preparation

1. Preparation of the GeoTiff for the estimated count of residual *Pf* malaria cases among all ages (0-99 years): See section I. Data inputs, Estimated *Plasmodium falciparum* malaria cases
2. Preparation of the processing order variable “rCases13” within the point vector file for the hypothetical network of CS-ASC:
   a. Based on the geographic coverage analysis of the hypothetical CS-ASC network, not considering capacity (Methods for Targeting research question 2, Data preparation, step 1) we used zonal statistics to calculate the estimated count of the residual *Pf* malaria cases beyond the geographic coverage of the existing CSI network in 2013 for the catchment area of each hypothetical CS-ASC site, using the GeoTiff of the estimated residual *Pf* malaria cases in Data preparation step 1 [ r_NER_residCases13_gcCSI_60min_1km] and the shapefile for the geographic coverage analysis of the hypothetical CS-ASC network from Targeting research question 1, Data Preparation, step 2 [r_NER_Targeting_Hypothetical_CS_ASC_FB13TTge500_60min_1km_no_capacity] in a variable called “rCases13”.
   b. We used a spatial join to join the variable “rCases13” in the shapefile from Data Preparation step 3a to the input point vector file [v_NER_Targeting_Hypothetical_CS_ASC_sites_FB13TTge500_1km] using the unique ID for each hypothetical CS-ASC to facilitate the join.

Analysis

1. Geographic coverage analysis of the estimated residual *Pf* malaria cases among all ages (0-99 years) by the existing network of CS-ASC, prioritizing estimated residual population in the processing order: We conducted a geographic coverage analysis for the estimated residual *Pf* malaria cases among all ages (0-99 years) beyond the geographic coverage of the existing CSI network, using the existing network of CS-ASC sties in 2013, with the processing order based on the estimated residual population “rFB13TT” within each catchment area and setting the maximum population capacity at 100000 to effectively not consider maximum population capacity as a constraint to the CS-ASC catchment areas. The analysis removed the estimated *Pf* malaria cases within each catchment area at each iteration (calculation of each catchment area) to avoid double counting estimated *Pf* malaria cases where the 60 min catchment areas overlap. This provided the final outputs for the analysis of geographic coverage of the estimated residual under-five deaths by the existing CS-ASC network.
   a. We used the following data inputs:
i. Population: r_NER_residCases13_gcCSI_60min_1km
ii. Land cover merged: raster_land_cover_merged_r_NER_land_merged
iii. Scenario table: table_scenario_walk_dry
iv. Select existing health facilities layer (vector): v_NER_cells_at_100m_with_CS_or_ASC_adj_barriers_1km
v. ID field: id
vi. Facility name field: cat
vii. Capacity: capacity
viii. Select zones layer (vector): adm3
      1. Select zones unique ID (integer): objectid
      2. Select zone name (text): nom_com
      b. We used the following analysis settings:
i. Type of analysis: anisotropic
ii. Direction of travel: towards facilities
iii. Facilities processing order according to: A field in the facility layer “rFB13TT”
iv. Processing order: Descending
v. Maximum travel time (minutes): 60
vi. Options
1. Compute population catchment area layer: Yes
2. Remove the covered population at each iteration: Yes
3. Compute a layer of population cells on barriers: Yes
4. Generate zonal statistics: Yes (adm 3)
5. Run the analysis without considering capacities: No
6. Add column with original population sum under each facility’s travel time: Yes
7. Optimize dynamically computation according to the scenario: Yes
8. Add short tag: r_NER_gcCS_ASC_rCases13_60min_1km_prioritize_rFB13TT
2. Geographic coverage analysis of the estimated residual *Pf* malaria cases among all ages (0-99 years) by the hypothetical network of CS-ASC, prioritizing estimated residual *Pf* cases in the processing order: We conducted a geographic coverage analysis for the estimated residual *Pf* malaria cases among all ages (0-99 years) beyond the geographic coverage of the existing CSI network, using the existing network of CS-ASC sties in 2013, with the processing order based on the estimated residual *Pf* malaria cases in 2013 “rCases13” within each catchment area and setting the maximum population capacity at 100000 to effectively not consider maximum population capacity as a constraint to the CS-ASC catchment areas. The analysis removed the estimated *Pf* malaria cases within each catchment area at each iteration (calculation of each catchment area) to avoid double counting estimated *Pf* malaria cases where the 60 min catchment areas overlap. This provided the final outputs for the analysis of geographic coverage of the estimated residual under-five deaths by the existing CS-ASC network.
   a. We used the following data inputs:
i. Population: r_NER_residCases13_gcCSI_60min_1km
ii. Land cover merged: raster_land_cover_merged_r_NER_land_merged
iii. Scenario table: table_scenario_walk_dry
iv. Select existing health facilities layer (vector): v_NER_cells_at_100m_with_CS_or_ASC_adj_barriers_1km
v. ID field: id
vi. Facility name field: cat
vii. Capacity: capacity
viii. Select zones layer (vector): adm3
1. Select zones unique ID (integer): objectid
2. Select zone name (text): nom_com
   b. We used the following analysis settings:
i. Type of analysis: anisotropic
ii. Direction of travel: towards facilities
iii. Facilities processing order according to: A field in the facility layer “rCases13”
iv. Processing order: Descending
v. Maximum travel time (minutes): 60
vi. Options
1. Compute population catchment area layer: Yes
2. Remove the covered population at each iteration: Yes
3. Compute a layer of population cells on barriers: Yes
4. Generate zonal statistics: Yes (adm 3)
5. Run the analysis without considering capacities: No
6. Add column with original population sum under each facility’s travel time: Yes
7. Optimize dynamically computation according to the scenario: Yes
8. Add short tag: r_NER_gcCS_ASC_rCases13_60min_1km_prioritize_rCases13

For outputs, see Supplementary Appendix 5, tabs “Malaria_Existing” and “Marlaria_Hypothetical”, in which the variable “amPopCoveredPercent” indicates the cumulative geographic coverage of the residual population. Supplementary Appendix 1b at https://doi.org/10.5281/zenodo.4482969 contains the vector shapefile (polygon) indicating the modelled catchment area of each health service delivery point.

3. Comparison of geographic coverage of the existing CS-ASC network and the hypothetical CS-ASC network: We compared the percentage of the estimated residual *Pf* malaria cases among all ages (0-99 years) beyond the geographic coverage of the existing CSI network in 2013 that was covered by the existing network of CS-ASC, prioritizing the estimated residual population in the processing order, with the percentage of the estimated residual *Pf* malaria cases among all ages (0-99 years) beyond the geographic coverage of the existing CSI network in 2013 that was covered by the hypothetical network of CS-ASC, prioritizing the estimated residual *Pf* malaria cases among all ages (0-99 years) in the processing order given the same number of potential CS-ASC sites as in the existing network of CS-ASC (i.e. 2550) as well as for the total number of potential CS-ASC sites (i.e. 5796).
a. In tab “Malaria_Existing” of Supplementary Appendix 5, the last value for the variable “amPercentCovered” provided the cumulative percent of estimated residual *Pf* malaria cases beyond the geographic coverage of the existing CSI network that were covered by the existing CS-ASC network in 2013.
b. In tab “Malaria_Hypothetical” of Supplementary Appendix 5, the last value for the variable “amPercentCovered” provided the cumulative percent of the estimated residual *Pf* malaria cases beyond the geographic coverage of the existing CSI network that were covered by the hypothetical CS-ASC network in 2013, prioritizing the estimated residual *Pf* malaria cases in the processing order.
c. In tab “Comparison_Malaria” of Supplementary Appendix 5, we compared the results from 3a and to the results from 3b for the first 2550 hypothetical CS-ASC sites in 2013 potential CS-ASC sites to ensure comparability with the existing CS-ASC network in 2013. We then compared the results from 3a with the results from the full network of 5796 hypothetical CS-ASC sites in 2013.

Uncertainty analysis

We assessed the potential effect of uncertainty of the estimates for under-five deaths on targeting as follows. We used the “Zonal statistics” tool in QGIS 3.12.0-Bucareşti^13^ to extract the estimated mean and 95% confidence intervals for the number of *Pf* malaria cases for all ages (0-99 years) for each catchment area defined by the geographic coverage analysis for the hypothetical network from step 2 of targeting research question 3. We sorted the catchments by the estimated mean number of *Pf* malaria cases for all ages (0-99 years) from largest to smallest, as this reflected the prioritization order of the geographic coverage analysis used for the targeting analysis (step 2 of targeting research question 3). Because policy makers and planners typically support scale-up of facilities and CHWs in groups or “blocks” we identified four potential “blocks” of CS-ASC for consideration. Block 1 included the 500 CS-ASC with the highest estimated mean number of *Pf* malaria cases, (median of means across catchments = 12866, lower 95% confidence interval minimum = 4892, and upper 95% confidence interval maximum = 146491). Block 2 included 389 CS-ASC with the next highest estimated mean number of *Pf* malaria cases (median of means across catchments = 3832, lower 95% confidence interval minimum = 3090, and upper 95% confidence interval maximum = 6984). Block 3 included 1134 CS-ASC with next highest estimated mean number of *Pf* malaria cases (median of means across catchments = 2107, lower 95% confidence interval minimum = 1608, and upper 95% confidence interval maximum = 4230). Block 4 included 3773 catchments with the next highest mean number of *Pf* malaria cases (median of means across catchments = 762, lower 95% confidence interval minimum = 0, and maximum 95% confidence interval maximum = 2214). Based on the 95% confidence interval minimums and maximums, the estimated number of *Pf* malaria cases was significantly higher in Block 1 than Block 3 or Block 4, and the estimated number of *Pf* malaria cases was significantly higher in Block 2 than Block 4, suggesting that policy makers and planners could confidently prioritize Block 1 over Block 3 and Block 4, and prioritize Block 2 over Block 4. (see Supplementary Appendix 6 – Targeting uncertainty, tabs “Summary_uncertainty_rCases13” and “Blocks_uncertainty_rCases13”.
